# Anaesthetic Higher Specialty Training Recruitment in the United Kingdom During the COVID-19 Pandemic: A National Survey

**DOI:** 10.1101/2021.07.03.21259616

**Authors:** Fionnuala Durrant, Stuart Edwardson, Sally El-Ghazali, Christopher Holt, Roopa McCrossan, Ileena Pramanik, Jeevakan Subramaniam, Danny J. N. Wong

## Abstract

The most recent ST3 Anaesthetic recruitment for posts commencing in August 2021 saw larger numbers of applicants (n = 1,056) compared to previous years, with approximately 700 applicants failing to secure an ST3 post. We surveyed 536 anaesthetic junior doctors who applied for ST3 posts during this application round with the aim of investigating their experience of the recruitment process this year (response rate 536/1,056 = 51%). Approximately 61% were not offered ST3 posts (n = 326), a similar proportion to that previously reported. We asked all respondents what their potential career plans were for the next 12 to 24 months. The majority expressed intentions to take up either CT3 top-up posts or non-training fellow posts from August 2021 (79%). Other options considered by respondents included: pursuing work abroad (17%), embarking on a career break (16%), taking up an ST3 post in intensive care medicine instead of anaesthetics (15%) and permanently leaving the medical profession (9%). A number of respondents expressed a desire to pursue training in a different medical specialty (9%). Some respondents expressed an intention to pursue further education or research (10%). A large proportion of respondents (42%) expressed a lack of confidence in being able to achieve the necessary training requirements to later apply for ST4 in August 2023. The majority of respondents reported not feeling confident in achieving GMC Specialty Registration in Anaesthesia in the future without a training number (75%), and that their wider life plans have been disrupted due to the impending time out of training (78%). We received a total of 384 free-text responses to a question asking about general concerns regarding the ST3 applications process. Sentiment analysis of these free-text responses indicated that respondents felt generally negatively about the ST3 recruitment process. Some themes that were elicited from the responses included: respondents feeling the recruitment process lacked fairness, respondents suffering burnout and negative impacts on their wellbeing, difficulties in making plans for their personal lives, and feeling undervalued and abandoned despite having made personal sacrifices to support the health service during the COVID-19 pandemic. These results suggest that junior anaesthetic doctors in the UK currently have a negative perception towards postgraduate training structures, which has been exacerbated by the COVID-19 pandemic, changes to the postgraduate training curriculum and difficulties in securing higher training posts.

## Introduction

Prior to the COVID-19 pandemic, a major change was planned to the UK anaesthetics training curriculum. This was initially due to be enacted in 2020, but was deferred to 2021 due to disruptions to the health service brought about by the pandemic. Previously, after basic anaesthetic training, a trainee in the UK would be eligible to apply for intermediate and higher training posts at the ST3 (Specialty Training Year 3) level, through a competitive application process including interview. However, with the 2021 anaesthetics curriculum, the transition between basic and intermediate training has been replaced by new milestones, renamed as Level 1 and Level 2 training, respectively [1].

This change in curriculum means that entry into intermediate and higher training at ST3 level will eventually cease and will instead be replaced by entry into Level 2 training at ST4 level. Therefore, if a trainee is unsuccessful at ST3 in the final recruitment round, they will have to apply in 2023 for ST4 entry. If an individual wishes to continue with anaesthetic training, they will have to find an non-training post in which they can complete all required educational and professional development competencies to be eligible to apply for ST4 entry in accordance with the new curriculum.

The curriculum transition, together with a number of other factors, may have resulted in increased competition during the most recent recruitment round for ST3 posts commencing in August 2021, with a larger number of applicants this year. According to figures released by the Royal College of Anaesthetists, 1056 applicants competed for 359 posts at ST3 level this year, while 865 were granted an interview [2].

This is a difficult time for many trainees who have worked hard over the past 12 months of the pandemic supporting their colleagues in Intensive Care Units (ICUs) up and down the country. From the start of the pandemic, the UK required rapid expansion of ICU capacity and anaesthesia trainees were among the first cohort of trainees redeployed to care for the increased numbers of patients requiring mechanical ventilation. It is perhaps unsurprising that many who took part in the applications process have publicly expressed disappointment with this year’s ST3 applications process [3,4].

Many anaesthetic trainees unsuccessful with applications this year have expressed confusion as to how bridge the gap between the termination of their current training appointments and rejoining training under the new curriculum. Therefore, we sought to find out what their plans were for the next 12 to 24 months, with the intention of informing relevant stakeholders at the Royal College, Association of Anaesthetists and employing organisations, so that they may best support trainees with their training and welfare needs.

## Methods

We conducted a survey in collaboration with the Association of Anaesthetists, collecting data from 20 May 2021 to 30 June 2021 via a Google Forms (Mountain View, California, USA) survey tool. The survey was constructed using a modified Delphi method: questions were drafted using themes derived from the UK national F2 Career Destinations Reports [5]. A pilot survey was conducted with a group of 8 trainees within the South London School of Anaesthesia (round 1). The feedback from this first pilot round was used to refine the survey questions which were then subject to internal peer-review at the Association of Anaesthetists Trainee Committee (round 2) before dissemination nationally.

The final survey was approved by the Association of Anaesthetists internal governance applications process, and is available from: http://bit.ly/ST3Anaesthetics2021. Full survey questions are available as Supplementary Material.

The survey was publicised via the Association of Anaesthetists email mailing list, and via the Association of Anaesthetists’ and the authors’ social media accounts on Twitter, WhatsApp and Facebook. To ensure as many trainees as possible were aware of the survey, the Heads of Schools of Anaesthesia for each UK postgraduate training region was contacted via email, and asked to distribute the survey within their regions. Periodic reminders were sent out at 1 month and 1 week prior to survey closure via the aforementioned email and social media routes.

## Analysis

We undertook an exploratory analysis of the response data. Possible duplicate submissions were identified and removed before analysis. Descriptive statistics of the response rate, respondent demographics, ST3 application outcomes and destinations, and answers to questions requiring Likert scale responses are reported. Means and standard deviations, or medians and interquartile ranges are reported for continuous variables, as appropriate to their distributions. Frequencies (numbers) and percentages are reported for categorical variables. Illustrative quotes are reported within the results text for the subset of questions requiring free-text responses. A sentiment analysis using a method described by Rinker [6] was performed to analyse responses to a free-text question asked at the end of the survey. Sentiment analysis is a computer natural language processing approach to investigating the attitudes and opinions expressed in texts: responses were algorithmically scored for text sentiment polarity (positive, negative, or neutral) at the sentence level against a dictionary of polarised words, with positive and negative words are scored with a +1 and -1, respectively. In this analysis, a score greater than zero indicated a positive sentiment, less than zero a negative sentiment, and zero a neutral sentiment, for the whole free-text response overall. This was conducted to rapidly screen the large number of free-text generated in the survey in lieu of a full qualitative thematic analysis. We then went through the free-text responses by hand to extract themes from the text. A free-text response software tool was constructed that facilitated the filtering of free-text responses by respondent characteristics, and which allowed the text to be read more easily by the authors via their web browsers (https://dannyjnwong.shinyapps.io/ST3RecruitmentSurvey/). Analysis was undertaken in R version 4.1.0 (2021-05-18) (R Foundation for Statistical Computing, Vienna, Austria). Code for analysis will be available from https://github.com/dannyjnwong/ST3Anaesthetics2021.

## Results

### Response rate

Of the total 765 responses submitted, 132 (17.2%) were not eligible to apply for an ST3 post due to commence in August 2021, and were excluded from analysis (Figure 1). The total number of responses included in analysis was therefore 633. The number of respondents who applied for an ST3 post was 536. Of the respondents who applied for an ST3 post, 210/536 (39.2%) were successfully appointed to an ST3 post, and 326/536 (60.8%) were unsuccessful with their application.

**Figure 1:**
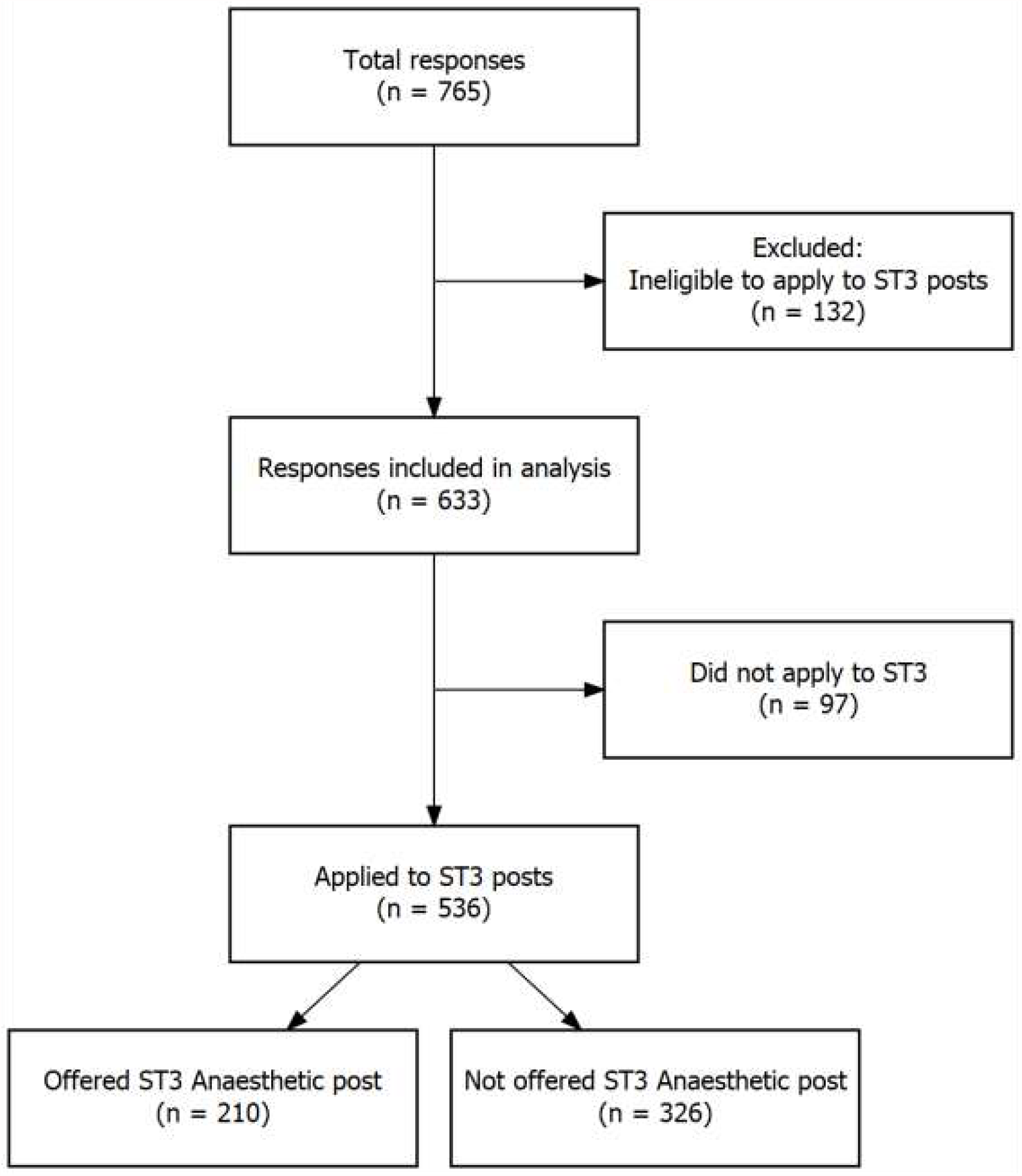
Flowchart of responses included in analysis.

Our survey therefore represents data from 50.8% of the 1,056 candidates who applied, or alternatively, 46.8% of the 697 candidates who were unsuccessful in obtaining an ST3 post in this application round.

## Demographics

The baseline demographics of respondents are reported in Table 1. The majority of respondents were over the age of 30 (330/633 = 52.1%) and reported their sex as Male (375/633 = 59.2%). We received responses from all postgraduate training regions across the UK. The median (IQR) number of years of clinical experience in anaesthesia completed by August 2021 reported by respondents was 3 (2 to 4), however a quarter of respondents reported between 4 and 14 years of experience (Figure 2).

**Table 1:** Respondent demographics

|  | *Overall* |
| --- | --- |
| **n** | 633 |
| **Age (%)** |  |
| 25-30 | 303 (47.9) |
| 31-35 | 262 (41.4) |
| 36-40 | 62 ( 9.8) |
| 41-45 | 6 ( 0.9) |
| **Sex (%)** |  |
| Female | 255 (40.3) |
| Male | 375 (59.2) |
| Prefer not to say | 3 ( 0.5) |
| **Current Region (%)** |  |
| Health Education and Improvement Wales | 33 ( 5.2) |
| Health Education East Midlands | 35 ( 5.5) |
| Health Education East of England | 46 ( 7.3) |
| Health Education Kent, Surrey & Sussex | 57 ( 9.0) |
| Health Education London | 119 (18.8) |
| Health Education North East | 36 ( 5.7) |
| Health Education North West (Mersey & North Western) | 78 (12.3) |
| Health Education South West (Peninsula & Severn) | 47 ( 7.4) |
| Health Education Thames Valley | 20 ( 3.2) |
| Health Education Wessex | 16 ( 2.5) |
| Health Education West Midlands | 40 ( 6.3) |
| Health Education Yorkshire and the Humber | 49 ( 7.7) |
| NHS Education for Scotland | 46 ( 7.3) |
| Northern Ireland Medical and Dental Training Agency | 3 ( 0.5) |
| Outside UK | 8 ( 1.3) |
| **Current Role (%)** |  |
| Acute Care Common Stem (ACCS) - Anaesthetics Trainee | 122 (19.3) |
| Anaesthetic Core Trainee | 179 (28.3) |
| Anaesthetic specialty registrar, applying to continue training in a different region | 1 ( 0.2) |
| Intensive Care Medicine Trainee | 24 ( 3.8) |
| LAT ST3 | 10 ( 1.6) |
| MTI Fellow | 3 ( 0.5) |
| Non-training Fellow, SAS or other Trust-Grade post | 277 (43.8) |
| Not currently in contracted employment, or currently a locum | 17 ( 2.7) |
| **Years of Clinical Experience in Anaesthesia Completed by August 2021 (median [IQR])** | 3 [2, 4] |
| **Current Contracted Working Pattern (%)** |  |
| Full-time | 559 (88.3) |
| LTFT | 52 ( 8.2) |
| Not currently contracted, or working as a locum | 22 ( 3.5) |

**Figure 2:**
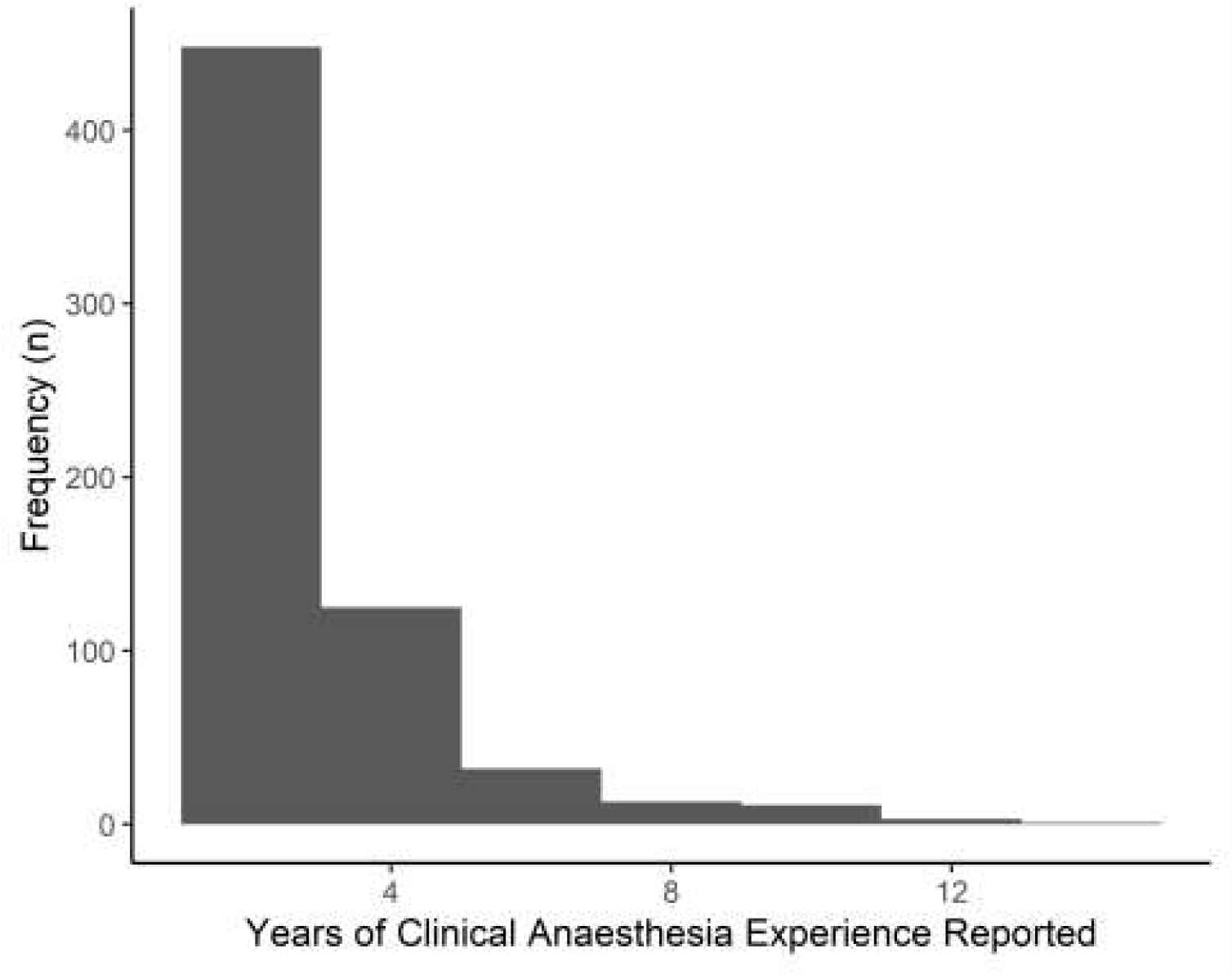
Distribution of years of clinical anaesthesia experience reported by respondents.

## ST3 Applications Outcomes

Of the 536 survey respondents that applied to ST3 anaesthetic posts, 210 respondents (39.2%) were successfully offered posts, while 326 respondents (60.8%) were not.

14 respondents were offered an ST3 post but turned down their offer, citing reasons including:

> **Issues with location of the offer (7 responses)**.
>
> **Accepting an offer in another specialty instead (4 responses)**.
>
> **Electing to pursue training in another country (1 response)**.

One notable quote from this group of respondents:

> **“After careful consideration, I have chosen to finish my training outside of the UK. I feel that a return to the NHS would entail significantly worse hours, pay, commutes, and working conditions - together with significantly longer training times. I would see my new daughter at least 2500 hours less over the next six years, and earn ∼£300K less over the same period. In addition, HEE’s conduct during the recruitment process - which was unfair, inconsistent, free from accountability, and treated trainees disgracefully - has reinforced to me that national training programmes and the bodies which oversea [sic] them are fundamentally toxic, and unfit for purpose.”**

## Reasons for not applying in this ST3 application round

97 respondents reported not applying during this ST3 recruitment round. The top 5 reasons cited were (respondents could offer more than one reason):

> **Awareness of increased competition ratios and/or reduced CV opportunities during COVID (50 responses [51.5%])**
>
> **Wanting to take a break from training (43 responses [44.3%])**
>
> **Wanting to pursue other interests or opportunities, e.g. research or management, etc. (24 responses [24.7%])**
>
> **Lacking core experience due to the COVID-19 pandemic (25 responses [25.8%])**
>
> **Planning to pursue training abroad (2 responses [2.1%])**

Some notable quotes from this group of trainees:

> **“Applications opened almost immediately after primary viva and was too exhausted to launch straight in to all of the preparation required.”**
>
> **“Didn’t realise there would be a freeze on recruitment and felt I needed more Obstetric and Paediatric experience before becoming a registrar.”**
>
> **“Am very unlikely to succeed in the modern tick-box environment of our specialty and our profession, so will likely be leaving medicine.”**
>
> **“Missed application window. Stressed/overwhelmed with COVID and exams.”**
>
> **“Love the job but find the environment, constant rotation, lack of autonomy in training very difficult and need a break to decide about whether to pursue a different career or keep going. I don’t think systems in place adequately support trainees to balance work stress against health.”**

38/97 of the respondents (39.2%) who did not apply in this ST3 application round were not aware that there was a planned freeze in ST3 recruitment from August 2022. While 83/97 of the respondents (85.6%) did not feel the effects of the curriculum changes on ST3 recruitment were well-explained to them prior to the ST3 application deadline (Table 2). Similarly, 40/97 of the respondents (41.2%) felt that more information regarding changes to the curriculum and recruitment may have changed their decision about not applying during this round.

**Table 2:** Responses to questions around curriculum changes.

| Question | No | Not sure | Yes |
| --- | --- | --- | --- |
| Did you feel the effects of curriculum changes on recruitment were well explained to you prior to the ST3 application deadline? | 85.5% | 3.1% | 11.3% |
| Would more information regarding changes to the curriculum and recruitment have changed your decision not to apply? | 39.2% | 19.6% | 41.2% |

## Future plans

A total of 437 respondents either did not apply for ST3 despite being eligible, or applied for ST3 and were not offered a post, or applied for ST3 and were offered a post which they turned down. These respondents represented 69.0% of the total responses (n = 633) included in analysis, and signify the cohort of anaesthetists for whom planning for the next 2 years during the transition to the new 2021 Anaesthetic Curriculum is associated with uncertainty.

These respondents were asked about their plans for the next 12 to 24 months from August 2021 as they did not have an ST3 Anaesthetic post secured. Respondents were allowed to specify more than one possible plan.

The majority of respondents expressed a plan to take up either CT3 top-up posts or non-training fellow posts from August 2021 (347/437 = 79.4%). These were followed in descending orders of frequency by pursuing work abroad (n = 76, 17.4%), embarking on a career break (n = 68, 15.6%), taking up an ST3 Intensive Care Medicine post in August 2021 (n = 64, 14.6%) and permanently leaving the medical profession (n = 41, 9.4%).

A number of respondents further expressed a desire to pursue training in a different medical specialty (n = 39, 8.9%), with possible destination specialties including intensive care medicine, general practice and emergency medicine cited. Some respondents expressed an intention to pursue a further education or research (n = 42, 9.6%). A small number of respondents whose existing roles were as Specialty/SAS Doctors expressed a plan to continue in that role (n = 6), and a similar number expressed a plan to pursue Specialty Registration via the Certificate of Eligibility for Specialist Registration (CESR) route (n = 8).

A large proportion of respondents expressed a lack of confidence in being able to achieve the necessary training requirements to later apply for ST4 in August 2023, and ultimately achieving GMC Specialty Registration in Anaesthesia without a training number (Figure 3). A substantial number of respondents expressed that their wider life plans have suffered disruption due to the impending time out of training (341/437 = 78.0%).

**Figure 3:**
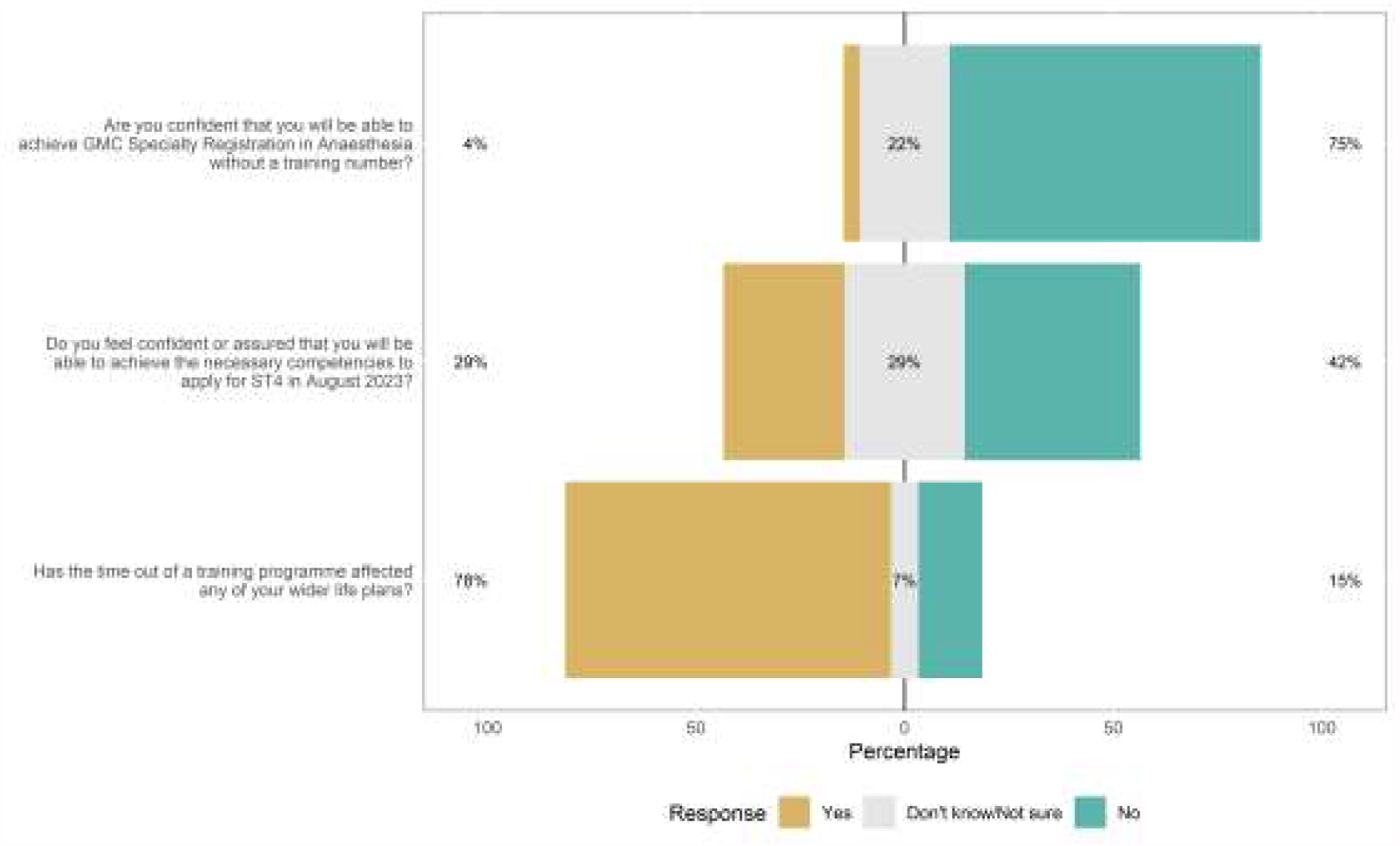
Responses to questions about future plans (n = 437).

## Sentiments of free-text responses

We allowed respondents the opportunity to describe their thoughts about the ST3 recruitment process in their own words via a free-text box at the end of the survey. They were asked, “What general concerns would you like to share with us regarding this year’s ST3 applications? If there are specific personal circumstances you are happy to share with the relevant stakeholders, please feel free to contribute them here. We may publish some of these as anonymised quotes to convey the depth of feeling around this issue.”

We received a total of 384 free-text responses to this question (see Supplementary Material and online [https://dannyjnwong.shinyapps.io/ST3RecruitmentSurvey/] for quotes).

The responses with the largest magnitude (most negative or most positive) sentiment scores as calculated by Rinker’s method were identified and highlighted in Table 3 (most negative sentiment scores) and Table 4 (most positive sentiment scores). The sentiment of the free-text responses was overall negative, with a mean (SD) score across all available responses of -0.06 (0.20), and a median (IQR) of -0.05 (−0.17 to 0.05). This suggested that responses to this question conveyed a generally negative sentiment. A range of topics were elicited in the free-text responses by the authors on hand-searching following sentiment analysis screening (Table 5).

**Table 3:** Top 5 free-text quotes that produced the most negative sentiment scores.

| *Sentiment score* | *Free-text quote* |
| --- | --- |
| -1.01 | Devastated, exhausted, disillusioned. |
| -0.72 | The whole process from online application to job allocation was extremely flawed and inherently unfair. All the processes put in place to try and communicate or appeal were difficult to use and over all the communication from ANRO was at best unfeeling and at worse callous with complete disregard of the magnitude of the importance of moving onto the next stage has on every part of a doctors life. Extremely disappointing. |
| -0.67 | Lack of transparency over recruitment process, discarding current anaesthetic trainees despite their sacrifices during the pandemic, reinforces feeling undervalued, overworked, and mistreated. These changes only add to the poor working environment that trainees have had to put up with in order to pursue a career they are passionate about - the lack of autonomy in placement/progress with no good alternative pathway to specialism is a real concern to me |
| -0.60 | I specifically withdrew from the process due to ill health so may not be wholly representative but that pressure of the high stakes nature of the process did not assist with my wellbeing. |
| -0.54 | Dreadful process which is damaging to the mental health of doctors. Made worse by the RCoA's response which belittled the feedback doctors gave them and denied the difficulties we have faced, especially in light of the hard work we have all put in in the pandemic. A real slap in the face. |

**Table 4:** Top 5 free-text quotes that produced the most positive sentiment scores.

| *Sentiment score* | *Free-text quote* |
| --- | --- |
| 0.72 | Provide more training slot [sic]. |
| 0.72 | Admission heavily favour [sic] those who have taken extra fellow years or pre anaesthetic medicine etc, putting the dedicated hard working core trainee at a disadvantage |
| 0.69 | Denary [sic] has no formal CT3 equivalency plans, left to us to arrange multiple fellow jobs to achieve right competence. |
| 0.54 | I think it's extraordinary that under the current system various parties involved in training will with one hand promote wellbeing initiatives, and with the other be complicit in trainees abandoning their friends/family/social support structures when national recruitment schemes require them to move hundreds of miles for jobs to continue training. It's not complicated - recognising the importance of being able to put down roots, build healthy relationships/friendships outside work, and develop a healthy work/life balance by having more control over where you live is key to promoting happiness. Mindfulness initiatives, whilst still valuable, are clearly not going to make up for abandoning these things to take a training post in a location that you have no links to and are moving to solely because of work. |
| 0.47 | I would like there to be clear guidance as to what we need to do for a CT3 equivalent year, along with access to the lifelong learning platform (or something like it) to evidence our training. |

**Table 5:** Topics elicited from free-text responses (total free-text reponses received, n = 384).

| *Responses mentioning the following themes* | *Number of responses* | *% of responses* |
| --- | --- | --- |
| Curriculum changes | 68 | 17.7 |
| Wellbeing, burnout and mental health | 56 | 14.6 |
| Feeling supported versus abandonment | 52 | 13.5 |
| Fairness | 51 | 13.3 |
| Communication | 22 | 5.7 |

## Discussion

A large proportion of applicants have been unsuccessful in the most recent round of ST3 Anaesthetic applications. This follows a particularly difficult year for the NHS where doctors-in-training from all specialties, especially anaesthetics, have faced significant disruption to their training in both reductions in elective lists and redeployment or supporting roles in intensive care. It is postulated that multiple factors have contributed to the increased ST3 competition ratios this year. These may include relaxation of previous requirements to secure the full Primary FRCA to apply to intermediate and higher training, a reduction in trainees seeking temporary work abroad and anticipation of a freeze in recruitment. Therefore, trainees may have seen this round as the final opportunity to enter training for 2 years with only a limited round of recruitment in February 2022. The results of the survey highlighted important themes — some specific to this year’s ST3 application round, but also some wider general training themes that should be further discussed in order to guide future recruitment.

Our survey reports data from approximately half of the 1056 candidates who applied, and a third of the 697 candidates who were unsuccessful in obtaining an ST3 post in this application round. Around a third of respondents included in this analysis have accepted an ST3 post, while around two thirds of respondents did not. The Royal College of Anaesthetists quoted a 34% applicant success rate for this application round. This parity suggests acceptable external validity of our study, and further suggests that we avoided sampling bias based on whether respondents were successful with their ST3 application. We gained responses from across all UK health boards, with the majority of respondents being males aged between 25 and 35. Only 5 respondents currently work outside the UK. Notably, while increased competition was cited as a reason for many going without job offers, over 30 of our respondents cited anticipated increased competition as a reason for avoiding application this year.

It was necessary to alter the recruitment format this year in order to respect social distancing and a potential reduction in available consultant interviewers. This followed a previously disrupted recruitment round in 2020 which saw applicants appointed on unverified self-assessment scores. Many respondents commented on the combination of what they thought were vague instructions for the self-scoring assessment and a virtual interview. One wrote that “…30 minutes by 2 consultants about non clinical experience — hardly a way to interview for future consultants.” This sentiment was repeated multiple times in free-text responses, and suggests a feeling among respondents that the recruitment process was “unfair, inconsistent, and free from accountability.” Whilst trainees accepted reasons why the recruitment format needed to alter this year, many respondents cited the experience as negatively affecting their faith in the recruitment process, and the bodies that govern it.

A major theme of the free-text responses was the negative impact of this recruitment process on trainee wellbeing. Approximately 4 out of 5 respondents told us that their life plans had been negatively affected by this year’s recruitment issues. Respondents described it as “stressful” especially due to its “impact on major life decisions/family planning/dependencies.” Many described worsening “feelings of burnout.” There is an existing large body of evidence that anaesthetic trainees are especially susceptible to work-related stress, burnout, and depression [7,8]. Indeed, one respondent commented on it being “extraordinary that under the current system various parties involved in training will with one hand promote wellbeing initiatives, and with the other be complicit in trainees abandoning their friends/family/social support structures when national recruitment schemes require them to move hundreds of miles for jobs to continue training.” The already recognised stressful characteristics of anaesthetic training are amongst the many things which have intensified over the last 12 months, especially in relation to ST3 recruitment. Anaesthetic trainees have been instrumental in the COVID-19 pandemic response, due to the skills they possess which made them suitable for redeployment onto Intensive Care Units with ventilated COVID-19 patients that experienced high mortality rates. The compounding negative impact this recruitment has added to the effects of a difficult 12 months are summarised by one respondent, “After diligently propping up surge ICUs, and caring for critically-ill COVID patients, the NHS has treated anaesthetists-in-training as single-use, disposable, items — burning them out on surge rosters when needed, and then casually discarding them, and leaving them without jobs or careers, when not.” Similar findings were reflected in the recently published report by the House of Commons’ Health and Social Care Committee on the “emergency” level of burnout reached in health and social care workers [9]. The resulting feelings of burnout and clinical fatigue are linked with future poor patient care, worsened clinical outcomes, and increased risk of medical error [10].

Prior to the pandemic, there were already significant waiting lists for elective operating. This has been further exacerbated by the necessary extended cancellation of many elective procedures over the last year [11,12]. In March 2021, only 64.4% of patients in England awaiting surgical treatment were seen within 18 weeks [13]. The government’s 92% target has not been met since February 2016 [14]. Notably, over 430,000 patients in England have been waiting over a year for surgical treatment. The Royal College of Surgeons of England published a report on the New Deal for Surgery which estimates an annual average 11% increase in the number of NHS elective procedures performed every year in order to meet the 18 week waiting time target by 2023/24. The report states that this increase will require 4,000 extra surgical consultants [15]. Outside this potential increase in surgical workload, the Royal College of Anaesthetists already estimates that an extra 600-700 anaesthetic trainees will be required over the next seven years in order to meet expected baseline future workforce demands [16]. The House of Commons’ Health and Social Care Committee have stated that it is “clear that workforce planning has been led by the funding envelope available to health and social care rather than by demand and the capacity required to service that demand.” Their recent report on burnout in the NHS points to future uncertainty in workforce planning and lack of a formal solution as a major contributor to significant stress within the workforce [9].

However, in the face of these requirements, the future ripple effect of this year’s recruitment has produced some concerning indicators. One in 10 of respondents expressed desire to pursue training in a different medical specialty as a direct result of their treatment this year. Approximately 1 in 5 said they would seek work abroad, and 1 in 10 indicated that they were planning to permanently leave medicine altogether.

A total of 437 respondents did not take up ST3 posts for various reasons, including: not applying for ST3 despite being eligible, applying for ST3 but were not offered a post, or applying for ST3 but turning down a post offered. Of those, the survey revealed that the majority of respondents expressed a plan to take up either CT3 top-up posts or non-training fellow posts from August 2021 (approximately 80%) as first priority plan for future job. However, these individuals raised concerns regarding the information they have received about these posts. Some respondents expressed desire for “clear guidance” as to requirements. Whilst there is recognition that relevant bodies had produced guidance, some respondents noted that this was not communicated effectively and lacked the necessary clarity. There were also concerns amongst those undertaking CT3 posts that “…training opportunities to achieve these competencies will not be easy to come by….” Working alongside trainees in programme, there are no assurances provided that the training needs of those in CT3 top-up posts will be equitable within the department. There is no formal recourse for these trainees to highlight training issues, as governing bodies ultimately have no authority over trusts and departments to ensure they deliver what they have recommended in their guidance. This was reflected in one respondent’s experience as a non-training fellow, “expecting anaesthetic departments to treat fellows out of training equally to trainees and facilitate the top-up competencies, despite no funding, is idealistic.” Some respondents felt that if the CT3 top up posts were centrally co-ordinated, this would be more equitable and avoid discrepancies between different areas of the UK and Schools of Anaesthesia.

Only a small number of survey respondents stated they would pursue CESR (Certificate of Eligibility of Specialist Registration) accreditation. This may be a suitable option for “…experienced anaesthetists from abroad (who) have lots of experience to offer our NHS and they should have the opportunity to become UK consultants….” Some respondents did express they wanted more information about the CESR pathway but highlighted concerns about the process as they felt it was “clunky and tiresome.” These responses reflect a lack of familiarity with the CESR process amongst UK trainees. Indeed, this process itself is currently increasingly complex with the need to bridge between the 2010 and 2021 curricula [17,18]. Independent collection and presentation of evidence for CESR assessment can easily result in collation of between 800-1,000 pieces of evidence collected and submitted by the individual without an infrastructure of assistance and supervision supplied when enrolled in UK specialty training. CESR also inherently asks for significant flexibility and clinical investment from the health trust that the individual is working within. This is something which is likely to have significant regional variation.

In the free-text responses, 44 respondents make reference to “poor communication” of the changes to recruitment and the selection process. Trainees described the process as “opaque” and “confusing” during a time of such significant change to vital processes. One trainee called for “improved lines of communication” and far more transparent and open publication of the decision-making processes that lead to these life-changing alterations.

There was an overall theme of feeling “disillusioned,” “frustrated,” “disheartened,” and “let down” amongst the free text responses. One trainee notes that “coupled with an exhausting and difficult 12+ months of covid, I feel undervalued and appreciated” by their regulatory bodies. Trainees made significant personal and professional sacrifices to work exhausting emergency surge rotas while not obtaining the necessary professional requirements to be competitive at ST3 application. Another trainee expands on this by saying “the amount of sacrifice this year to not be given a training job feels beyond insulting.” The responses to this questionnaire give a clear indication that there is a sense of disconnect now felt between trainees and their regulatory bodies.

Many trainees also feel “vulnerable” for not having support to find verified CT3 top up training, and still feel unsure as to how they can be guaranteed that these training experiences will count upon re-application to training at ST4 level. This survey demonstrates a consistent sentiment that clear guidance, support, and assurance from regulatory bodies to show a transparency of assessment and validation will be key to tempering this vulnerability, and start to bridge the disconnect which has emerged.

The strengths and limitations of this survey must be considered when interpreting its findings. The data represent a candid account from junior doctors affected by this year’s ST3 recruitment process and the survey was rapidly disseminated with minimal delay after the recruitment process ended, thus minimising recall bias. We obtained responses from across the UK, thus allowing for generalisability of our findings between regions. However, due to the need for rapid analysis we relied on algorithmically-assisted sentiment analysis to screen free-text responses for themes and topics, and while the method of sentiment analysis adopted provided a means of scoring responses according to a dictionary of words with ascribed polarities, this approach did result in text passages that still appeared generally negative to the human eye despite being scored positively by the algorithm.

This survey has clearly highlighted perceived issues in the ST3 recruitment process 2021 that needs to be further addressed in order to guide future workforce planning and the delivery of training. Ideally, the highlighted discontent and frustration felt by some trainees will help relevant stakeholders better understand what would be of benefit going forward. Regardless of the decision an individual trainee has taken in light of the recruitment process, it is imperative that they are fully supported locally, regionally and nationally. Trainees have raised a lack of clear access to information regarding competencies required for the standalone posts and how these should be signed off. One solution could include junior anaesthetic doctors being provided a supervisor affiliated with the RCoA in temporary non-training posts to ensure adherance to published College guidance regardig such posts. From a CESR point of view, trainees have expressed that the GMC and RCoA should produce clearer guidance regarding the paperwork needed to achieve this, as well as highlighting the advantages and disadvantages of this pathway. The key to achieving the above is clear communication and easy access to the relevant information. Navigating the complexity of workforce planning must involve a collaborative approach between relevant stakeholders, membership organisations and trainees in order to secure the future of the specialty.

## Supporting information

Full survey questions

## Data Availability

Data and code available on GitHub.

https://github.com/dannyjnwong/ST3Anaesthetics2021

## Data Availability

Data and code available on GitHub.

https://github.com/dannyjnwong/ST3Anaesthetics2021

## Data Availability

Data and code available on GitHub.

https://github.com/dannyjnwong/ST3Anaesthetics2021

## Acknowledgements

We thank the Association of Anaesthetists for their assistance in dissemination of the survey to their members.

## References

1. Royal College of Anaesthetists. 2021 Curriculum for a CCT in Anaesthetics. 2021. https://www.rcoa.ac.uk/training-careers/training-anaesthesia/2021-anaesthetics-curriculum

2. Royal College of Anaesthetists. Anaesthesia National Recruitment 2021 applicants CT1 and ST3. 2021. https://www.rcoa.ac.uk/news/anaesthesia-national-recruitment-2021-applicants-ct1-st3

3. Lintern S. ‘A slap in the face’: Hundreds of frontline Covid doctors told they won’t have jobs from August. The Independent, 2021. https://www.independent.co.uk/news/health/covid-nhs-junior-doctors-jobs-b1843757.html

4. Munro C. Nearly 700 anaesthetists have training interrupted after cuts. BMJ 2021: n1213. https://www.bmj.com/lookup/doi/10.1136/bmj.n1213

5. UK Foundation Programme. UK National F2 Career Destinations Reports. 2021. https://foundationprogramme.nhs.uk/resources/reports/

6. Rinker TR. Sentimentr: Calculate Text Polarity Sentiment. 2019. https://github.com/trinker/sentimentr

7. Looseley A, Wainwright E, Cook TM et al. Stress, burnout, depression and work satisfaction among UK anaesthetic trainees; a quantitative analysis of the Satisfaction and Wellbeing in Anaesthetic Training study. Anaesthesia 2019; 74: 1231–9. doi: 10.1111/anae.14681

8. Larsson J, Rosenqvist U, Holmström I. Being a young and inexperienced trainee anesthetist: A phenomenological study on tough working conditions. Acta Anaesthesiologica Scandinavica 2006; 50: 653–8. doi: 10.1111/j.1399-6576.2006.01035.x

9. Health and Social Care Committee. Workforce Burnout and Resilience in the NHS and Social Care - Second Report of Session 2021. House of Commons, 2021. https://publications.parliament.uk/pa/cm5802/cmselect/cmhealth/22/2202.htm

10. West CP, Tan AD, Habermann TM, Sloan JA, Shanafelt TD. Association of resident fatigue and distress with perceived medical errors. JAMA 2009; 302: 1294–300. doi: 10.1001/jama.2009.1389

11. Fowler A, Abbott TEF, Pearse RM. Can we safely continue to offer surgical treatments during the COVID-19 pandemicã BMJ Qual Saf 2021; 30: 268–70. doi: 10.1136/bmjqs-2020-012544

12. Fowler AJ, Dobbs TD, Wan YI et al. Resource requirements for reintroducing elective surgery during the COVID-19 pandemic: Modelling study. British Journal of Surgery 2021; 108: 97–103. doi: 10.1093/bjs/znaa012

13. Moon D. Statistical Press Notice: NHS Referral to Treatment (RTT) Waiting Times Data. NHS England & NHS Improvement, 2021. https://www.england.nhs.uk/statistics/wp-content/uploads/sites/2/2021/05/Mar21-RTT-SPN-publication-version-1.pdf

14. Charlesworth A, Watt T, Gardner T. Returning NHS waiting times to 18 weeks for routine treatment. The Health Foundation, 2020. https://www.health.org.uk/publications/long-reads/returning-nhs-waiting-times-to-18-weeks

15. Royal College of Surgeons (England). A New Deal for Surgery. 2021. https://www.rcseng.ac.uk/about-the-rcs/government-relations-and-consultation/position-statements-and-reports/action-plan-for-england/

16. Royal College of Anaesthetists. Medical Workforce Census Report 2020. 2020. https://www.rcoa.ac.uk/sites/default/files/documents/2020-11/Medical-Workforce-Census-Report-2020.pdf

17. Royal College of Anaesthetists. CESR and Equivalence. 2021. https://www.rcoa.ac.uk/training-careers/working-anaesthesia/cesr-equivalence

18. Fletcher S. Certificate of Eligibility for Specialist Registration. The SAS Handbook 2020. Fourth. Association of Anaesthetists, 2020. 40–2. https://anaesthetists.org/Portals/0/PDFs/Guidelines%20PDFs/The_SAS_Handbook_2020.pdfãver=2020-01-07-101915-670

