## Supplementary Material for "Anaesthetic Higher Specialty Training Recruitment in the United Kingdom During the COVID-19 Pandemic: A National Survey"

Here we present all the free-text responses submitted to the survey with some anonymised demographic characteristics of the respondent provided for context.

Supplementary Table 1: Free-text responses and sentiment scores, arranged by sentiment score, from most negative to most positive.

| Age | Sex | Current Role | What general concerns would you like to share with us? | Sentiment score |
| --- | --- | --- | --- | --- |
| 31-35 | Male | Non-training Fellow, SAS or other Trust-Grade post | Devastated, exhausted, disillusioned. | -1.01 |
| 31-35 | Female | Anaesthetic Core Trainee | The whole process from online application to job allocation was extremely flawed and inherently unfair. All the processes put in place to try and communicate or appeal were difficult to use and over all the communication from ANRO was at best unfeeling and at worse callous with complete disregard of the magnitude of the importance of moving onto the next stage has on every part of a doctors life. Extremely disappointing. | -0.718 |
| 25-30 | Female | Anaesthetic Core Trainee | Lack of transparency over recruitment process, discarding current anaesthetic trainees despite their sacrifices during the pandemic, reinforces feeling undervalued, overworked, and mistreated. These changes only add to the poor working environment that trainees have had to put up with in order to pursue a career they are passionate about - the lack of autonomy in placement/progress with no good alternative pathway to specialism is a real concern to me | -0.672 |
| 31-35 | Male | Non-training Fellow, SAS or other Trust-Grade post | I specifically withdrew from the process due to ill health so may not be wholly representative but that pressure of the high stakes nature of the process did not assist with my wellbeing. | -0.596 |
| 25-30 | Female | Non-training Fellow, SAS or other Trust-Grade post | Dreadful process which is damaging to the mental health of doctors. Made worse by the RCoA’s response which belittled the feedback doctors gave them and denied the difficulties we have faced, especially in light of the hard work we have all put in in the pandemic. A real slap in the face. | -0.541 |
| 31-35 | Female | Not currently in contracted employment, or currently a locum | I feel the limited interview provided poor discrimination between candidates. The weighting of interview vs self score placed too much emphasis on ‘performance’ on the day rather than years of work. Candidates were forced to take jobs out of region, making the next years of their lives more challenging with a long commute, due to limited alternative options. | -0.536 |
| 31-35 | Female | Acute Care Common Stem (ACCS) - Anaesthetics Trainee | Lack of transparency about how competitive it would be this year/lack of workforce planning and inability to follow guidelines for awarding portfolio points | -0.521 |
| 25-30 | Male | Anaesthetic Core Trainee | Upsetting and disappointing that we have been messed about with higher training, especially after the hard work put into COVID. | -0.514 |
| 31-35 | Male | Non-training Fellow, SAS or other Trust-Grade post | The colleges lack of insight is ruining my life. | -0.5 |
| 25-30 | Female | Anaesthetic Core Trainee | Unable to apply for mortgage as no work contract available greater than 1 year | -0.458 |
| 25-30 | Female | Acute Care Common Stem (ACCS) - Anaesthetics Trainee | Unfair competition ratios | -0.433 |
| 25-30 | Female | Anaesthetic Core Trainee | No regard for trainees with the lack of recruitment in 2022. Leaving massive competition ratios for the next 3-4 years undoubtedly. In post covid core training. Really outrageous. | -0.43 |
| 25-30 | Male | Acute Care Common Stem (ACCS) - Anaesthetics Trainee | With the additional stress of the curriculum change, if my wife (also an ACCS anaesthetics CT3) and I don’t get a training post starting February 2022 we will have to significantly delay/alter many life plans and goals which adds greatly and unnecessarily to our stress levels. I wish the college would delay the curriculum change and it only compounds issues with anaesthetic recruitment. | -0.429 |
| 25-30 | Female | Non-training Fellow, SAS or other Trust-Grade post | Scored down on self assessment (given 1 mark for a masters degree instead of 4) despite presenting the same evidence as precious rounds of applications. Unable to appeal as did not meet 5 mark threshold but points would have made the difference to me getting an interview which I didnt get. Tried to complain but told my complaint fell outside of the complaint policy. So missed out on the chance for an interview with no route to appeal which seems unfair. | -0.411 |
| 25-30 | Male | Acute Care Common Stem (ACCS) - Anaesthetics Trainee | A constant shifting of the goal posts in very uncertain times. | -0.407 |
| 31-35 | Male | Not currently in contracted employment, or currently a locum | Having taken time out and there being no recruitment next year, I will end up having had â€œtoo much experienceâ€ to apply to ST4 in 2023 | -0.4 |
| 31-35 | Male | Intensive Care Medicine Trainee | Poor instructions during the self assessment section (not clear that cv was needed to be uploaded or that points would be awarded for how easy it was to review the evidence and that these points would be more than most of the other sections) | -0.398 |
| 31-35 | Male | Anaesthetic Core Trainee | Lack of NTNs relative to applicants, poor comms from ANRO | -0.395 |
| 41-45 | Male | Anaesthetic Core Trainee | Very disruptive to life plans Inconsistent scoring of portfolio…at least not in accordance of what was laid out by the college | -0.39 |
| 41-45 | Male | Non-training Fellow, SAS or other Trust-Grade post | Online interview was very difficult to participate in - no time to answer fully as so many questions and delay in connection which was stressful. I will have to move my family to another region as did not get a post in my current one. It just seems that all the issues this year could have been avoided - I think it’s ridiculous to change the curriculum during such a time, therefore massively increasing competition and leaving hundreds of trainees without training posts unnecessarily. | -0.39 |
| 36-40 | Male | Non-training Fellow, SAS or other Trust-Grade post | Interviewed for 30 minutes by 2 consultants about non clinical experience - hardly a way to interview for future consultants. So few jobs, and yet there are CT3 posts, these could have been ST3’s. Feel betrayed and unsupported after being redeployed to cover the COVID pandemic. | -0.375 |
| 25-30 | Male | Anaesthetic Core Trainee | The job i accepted wasn’t the ideal location for me but felt like I couldn’t not accept a training post given the competition and the time I’d need to take out of training to reapply | -0.368 |
| 31-35 | Female | Anaesthetic Core Trainee | Incredibly disheartened that I qualified with enough points for a job but just not enough jobs available | -0.361 |
| 36-40 | Male | Non-training Fellow, SAS or other Trust-Grade post | Opening the process up to anyone with the MCQ felt unfair as I worked really hard to pass the viva and osce, and they are much harder than the MCQ alone. | -0.356 |
| 25-30 | Female | Non-training Fellow, SAS or other Trust-Grade post | It has added more stress and anxiety and I have noticed a deterioration in my mental health. I am worried that recruitment in to ST4 will remain just as competitive and that my progression through training will be delayed by many years. At times this has made me think about leaving anaesthetics and leaving a career in medicine altogether. | -0.355 |
| 25-30 | Male | Anaesthetic Core Trainee | I feel the response from the college to be very vague and slow out of the gate. Many issues were raised by trainees upon announcing the issue, but seem not to have been worked out yet. We also get very bland responses about our mandated year out as ‘opportunities to do interesting things,’ when in fact what we need is a proper apology for the kick in the teeth to this enforced delay in training and recognition that the college has let us down with this. | -0.344 |
| 31-35 | Female | Intensive Care Medicine Trainee | I feel very disappointed and used. I was redeployed to help on ICU and this is how I am thanked - by not getting a ST3 post and therefore being unemployed. I feel exhausted, demoralised and I am now considering a career in Big Pharma. | -0.341 |
| 25-30 | Male | Acute Care Common Stem (ACCS) - Anaesthetics Trainee | Lack of clarity for those who obtain an ICM training number without having completed a CT3 equivalent year in anaesthetics and then want to dual train is very frustrating and makes us unable to plan a career. | -0.337 |
| 25-30 | Female | Anaesthetic Core Trainee | So disappointed and let down by HEE and RCOA. | -0.333 |
| 31-35 | Female | Non-training Fellow, SAS or other Trust-Grade post | The portfolio points scoring wasn’t transparent and completely unfair - they didn’t stick to the published mark schemes (e.g. I was marked down for not providing a CV when the mark scheme clearly said not too - and my comments on this were ignored on appeal). The questions asked at interview were the exact same during the entire interview period which grossly disadvantaged those interviewed in the first week. Very difficult interview where the interviewers seemingly had a checklist of questions to get through - zero back & forth communication so no opportunity to build a rapport or display who we really are as people. | -0.324 |
| 25-30 | Female | Anaesthetic Core Trainee | Given a lot of hospitals can’t fully staff their current anaesthetics on call rotas it seems crazy that people were declined jobs. | -0.32 |
| 25-30 | Male | Non-training Fellow, SAS or other Trust-Grade post | Fuck the RCoA - and you can quote me on that. | -0.316 |
| 31-35 | Female | Non-training Fellow, SAS or other Trust-Grade post | Significant number of ST posts in the region at 0.8 or 0.6 and no increase in recruitment numbers making it challenging to get into training | -0.313 |
| 31-35 | Male | Non-training Fellow, SAS or other Trust-Grade post | Lack of training posts is a disgrace seeing there are significant gaps at consultant level. The whole application process remains fundamentally flawed and does not find the best trainees - still place too much emphasis on poor quality audit and publication instead of ability, experience or potential | -0.313 |
| 25-30 | Female | Intensive Care Medicine Trainee | I am devastated by the change in curriculum and potential to be stuck as single CCT in ICM. I feel abandoned by the RCOA. | -0.306 |
| 31-35 | Male | Non-training Fellow, SAS or other Trust-Grade post | ANRO communication and explanation is rubbish | -0.306 |
| 31-35 | Female | Non-training Fellow, SAS or other Trust-Grade post | - frustrated that goal posts were moved regarding eligibility to apply without full primary FRCA - with retrospect I would certainly have got a job if I had omitted the VIVA/SOE and associated revision and focussed on audit loops instead to gain points - no clear back up plan for this disruptive change in curriculum - why is there such a mismatch and bottle neck regarding CT and ST anaesthetics posts - why is it not a run through training program - unclear why there is no 2022 intake - i will have had 5 years out of training by 2023. Concerned this will count against me. | -0.302 |
| 36-40 | Male | Acute Care Common Stem (ACCS) - Anaesthetics Trainee | Shambolic and unnecessary changes | -0.3 |
| 31-35 | Female | Anaesthetic Core Trainee | I had a lot of anxiety surrounding this application process. As a female trainee trying to plan a family in the next 2 years the chance of not getting a job would potentially mean that much more uncertainty in relation to maternity pay/ leave/ return to work support etc and made me feel very unsure and exposed. Luckily I did get a job, though not until one of the final rounds, however I suspect there will be many in this position. | -0.284 |
| 25-30 | Female | Anaesthetic Core Trainee | Felt like a kick in the teeth after what was the worst year with great personal sacrifices and an interview that asked so heavily questions around stress/burnout. | -0.283 |
| 31-35 | Male | Acute Care Common Stem (ACCS) - Anaesthetics Trainee | Our consultants in the hospital are worked very hard, particularly during the pandemic. Elective lists are being cancelled every day due to lack of anaesthetic cover, at a time where the waiting lists have never been longer. Surely we need more senior trainees, for future consultant jobs in order for us to catch up on the work postponed by COVID? | -0.282 |
| 31-35 | Male | Non-training Fellow, SAS or other Trust-Grade post | The portfolio scoring seems unreasonable and bot being able to justify them seems unfair. | -0.267 |
| 25-30 | Male | Non-training Fellow, SAS or other Trust-Grade post | Lack of posts. Lack of prior information regarding plans for recruitment over next few years | -0.267 |
| 41-45 | Male | MTI Fellow | Very much competitive with limited seats | -0.265 |
| 25-30 | Male | Anaesthetic Core Trainee | It was handled terribly, poor communication, have not even been given CT3 posts, rather have been left to fend for ourselves to find and apply for non training top-up posts. Which are also paid far less than training posts. Terribly disappointed especially as we as a specialty really stepped up during COVID. | -0.265 |
| 31-35 | Male | Anaesthetic Core Trainee | Fighting a pandemic is hard enough without wondering if you’ll have a job to go into afterwards. I saw the crunch coming in November last year which helped my preparation but also added immeasurable stress. I have an ST3 post but getting it left me as burnt out as the pandemic. This was an avoidable and predictable situation which should be remedied with a very significant recruitment round in January 2022 and an apology from the GMC and HEE who both bear more responsibility than the RCOA. | -0.26 |
| 31-35 | Female | Anaesthetic Core Trainee | This year has been made throughly miserable with the curriculum changes imposed. CT2s should be afforded the appropriate top up post by default since this has been imposed upon us with poor notice and during a pandemic. The ST3 recruitment process was ok but has left people unfairly disadvantaged. As someone who gained the primary I was not offered an extension on my training compared to peers that didn’t get their exams. I have taken a specialty post which was not by choice but due to the sheer competition for jobs I was concerned I would lose out if I didn’t take the first one I was successful in. There is a lot of talk about welfare but quite frankly there is very little action or support and I think we have been treated abysmally by the college. | -0.258 |
| 31-35 | Male | Non-training Fellow, SAS or other Trust-Grade post | I don’t have faith in the training system anymore to handle the back log nor have the flexibility to recognise OOPE. I am actively pursuing CESR rather than awaiting a training number in anaesthetics. | -0.257 |
| 31-35 | Male | Non-training Fellow, SAS or other Trust-Grade post | Making the applicants apply without finishing the primary made those who did it compete with those who didnt, also i know some successful candidates who got a job, havent finished their core training certificate yet, which is not fair ! | -0.256 |
| 36-40 | Male | Non-training Fellow, SAS or other Trust-Grade post | Thought I would be more useful as a consultant, I was in my home country. | -0.256 |
| 25-30 | Female | Anaesthetic Core Trainee | I felt generally frustrated that the post numbers have been cut. This is especially disheartening following the COVID surge, the burden of which was hugely carried by anaesthetic trainees. It was also timed badly with a change in curriculum meaning that the seems to be a bottle neck of people with real important skills who are being denied training oppourtinities. | -0.254 |
| 31-35 | Female | Non-training Fellow, SAS or other Trust-Grade post | It is now unfair that people who have not passed the primary FRCA are allowed to apply for ST3. | -0.252 |
| 31-35 | Male | Acute Care Common Stem (ACCS) - Anaesthetics Trainee | The communication from the college and ANRO was very poor. The lack of training posts for doctors that have already finished core training and have slogged themselves the past 18 months is a disgrace. A few claps on a doorstep mean nothing when HEE can’t sort jobs for anaesthetists already in training. | -0.244 |
| 25-30 | Male | Anaesthetic Core Trainee | General feeling of being unappreciated, let down and abandoned. The college should have prioritised current trainees and smoothed the process into ST3 if they are adamant with pushing forward with curriculum changes. Instead, the circumstances were set up to favour those out of training who have been able to CV build while us trainees have been redeployed to ICU and spent 18 months dealing with Covid. How does someone complete core training, plus additional ACCS experience, take part in audit, research, exams and generally complete all requirements without issue and excellent feedback, only to be left jobless and in a position that massively impacts maternity/ paternity leave, pay, job security and career progression? Beyond insulting it’s extremely stressful and disheartening, particularly when this looks unlikely to improve for potentially at least two years to come. Am I expected to continue putting in this enormous level of effort and time, step up for an inevitable further wave of Covid, only to be unsuccessful again in February? In that situation I would be seriously reconsidering my career choices. | -0.243 |
| 36-40 | Male | Non-training Fellow, SAS or other Trust-Grade post | Massively dissapointed regarding the training posts available even though UK need more and more anaesthetist in future. | -0.243 |
| 31-35 | Female | Non-training Fellow, SAS or other Trust-Grade post | I understand there are frustrations around this years recruitment, however many of us were treated badly at least year’s August 2020 recruitment (being unsuccessful in getting posts which were given out based on unverified self assessment scores). Last year’s August recruitment did not gain the same attention that this year’s did, arguably under much wore circumstances as we could not be interviewed. I am glad the recruitment issues are coming to light but as someone who has done an extra year already based on last year’s terrible circumstances I do feel let down by AAGBI and RCOA that this was not dealt with last August. | -0.243 |
| 36-40 | Female | Non-training Fellow, SAS or other Trust-Grade post | UK has problem retaining current and past trainees in UK | -0.237 |
| 25-30 | Female | Anaesthetic Core Trainee | 1) Huge discrepancy between number of CT1 training posts vs ST3 training posts. 2) Interview system is inadequate. 30minutes on zoom does not give an accurate representation of candidates. With covid testing no widely available, I believe the college should look to return to face-to-face interviews ASAP. 3) Points awarded on the portfolio score for online conferences - I have not had the opportunity to present face-to-face since starting anaesthetics due to the pandemic despite having had work accepted into two separate conferences. Online posters did not score as highly as face-to-face poster presentations. This disadvantaged those who started training in the year of the pandemic. | -0.236 |
| 25-30 | Male | Anaesthetic Core Trainee | My mental health has been severely affected. After covid and the stress of exams, it has been such a gut wrench to be cut adrift by the lack of jobs and the new curriculum changes | -0.236 |
| 25-30 | Female | Non-training Fellow, SAS or other Trust-Grade post | Lots of problems and I’m exhausted by having to explain them on multiple surveys and emails to various people. Unclear transition plan has added stress. Don’t see why we need a 18 month break in recruitment. Don’t understand why OSCE/SOE not counted in ST3 application any more. Feel very demoralised by multiple attempts and getting nowhere. I work with people that have training posts obtained in previous years who have less than me in their CV. Online portfolio scoring was unclear - didn’t match guidance and videos. Got points for organisations but no guidance on what optimal organistion was. I lost 1 point for not combining my PDFs - nowhere told me to do this. | -0.234 |
| 31-35 | Female | Non-training Fellow, SAS or other Trust-Grade post | Ongoing vast number of rota gaps for registrar grades yet no more availability of training jobs. No clear reassurance of resolution for this issue in a cohort of burntout anaesthetists. Difficult to see how this would inprove in the future. | -0.231 |
| 31-35 | Male | Anaesthetic Core Trainee | Concerns of facing unemployment. There has been no support or consideration from RCoA or HEE. Vacuous sentiments from RCoA about how great our contribution was during the pandemic feels like a pathetic attempt at placating us. RCoA have published statements regarding the shortage of anaesthetists, their vital position within hospitals and the enormous backlog from COVID-19 which adds to the pre-existing backlog of surgical cases waiting, whilst simultaneously leaving this number of trainees to fall off the training scheme. I can’t believe the RCoA has had any serious negotiation with HEE to have allowed this situation to happen. | -0.23 |
| 25-30 | Female | Anaesthetic Core Trainee | I feel extremely let down that despite the year of disruption and the demands place on me as an anaesthetics trainee that I have not been able to get the job I want and deserve. It is ridiculous and insulting that there are not more places available, given that there is a deficit in consultants and an enormous backlog of surgery to catch up on. I have considered leaving the speciality, the region and the country because of this mess. I am devastated and exhausted and angry that I have been let down in this way. | -0.228 |
| 31-35 | Female | Non-training Fellow, SAS or other Trust-Grade post | The lack of training places and therefore competition for places was a huge concern this year and will continue into Feb recruitment. I think there has been a bit of discrepancy with the marking for organisation for self score between candidates; which for some people has been the difference between getting a job and not. The appeals process was also not acceptable. Causing lots of stress for people already struggling with a stressful application process. (although these did not affect me personally) | -0.227 |
| 25-30 | Male | Non-training Fellow, SAS or other Trust-Grade post | The lack of depth in interview feedback analysis. What feels like a lack of transparency in the whole process. The heightened anxiety from using a system that processes the offer in waves. | -0.226 |
| 25-30 | Male | Anaesthetic Core Trainee | The RCOA should be more transparent and active in their communications about why the bottle neck has occurred, how they plan to improve it (or not), and evidence that they are advocating on our behalf to HEE - where they lay blame for recruitment decisions. | -0.226 |
| 31-35 | Female | Non-training Fellow, SAS or other Trust-Grade post | I will have to spend further time in a very demanding rota due to lack of available fellow posts in the area. | -0.224 |
| 25-30 | Male | Non-training Fellow, SAS or other Trust-Grade post | Video in my interview didn’t work so I could not see any of my interview panel. Subsequently I saw my feedback in communication was marked down because â€œdid not respond to signallingâ€. ANRO have stated that interviews should only be scored on content on answers but this can not be appealed in anyway. Very disheartening when you miss a job by 1 or 2 points. | -0.223 |
| 31-35 | Male | Non-training Fellow, SAS or other Trust-Grade post | The general portfolio scoring to short list candidates undermines certain candidates who have other experiences that can be explored at interview. Some of the scoring criteria in the portfolio seem totally unrealistic. Rather than increasing training numbers to be reducing them seems counterintuitive. | -0.217 |
| 25-30 | Male | Anaesthetic Core Trainee | The fact that HEE don’t seem to care and that a repeat of this will happen in 2 years if nothing is done | -0.213 |
| 31-35 | Male | Non-training Fellow, SAS or other Trust-Grade post | The interview process is not fair, there should be standardisation of the process | -0.208 |
| 25-30 | Female | Anaesthetic Core Trainee | I feel the large gap without recruitment is going to lead to even higher competition ratios. The RCOA seem completely clueless regarding future work force planning and the delays in applications to 2023 just been even more competition for junior trainees. I love anaesthetics but am unsure if I will ever complete training the juniors of SHO grade have had the worst training disruptions due to covid and now feel as though we have been punished again. I also think this unfairly implicates female trainees - my life and family plans either have to be put on hold or I face much worse maternity leave prospects than my predecessors due to having to find funded CT3 jobs at trusts. | -0.205 |
| 25-30 | Female | Intensive Care Medicine Trainee | Very unhappy that training numbers were reduced after the year that we’ve had! None of my colleagues got a London job either. I’m considering leaving anaesthetics and pursing single speciality ICM training. Awful timing to change the syllabus. It was very unclear from the interview what domains were being assessed and difficult to prep for. | -0.204 |
| 25-30 | Male | Anaesthetic Core Trainee | RCOA wrote on its website (in FAQs re: curriculum changes) that one would be able to do an OOPE after CT2 (ie return to an HEE CT3 post). My OOPE application was rejected on the basis there would be no guaranteed HEE approved CT3 posts. RCOA later took this statement down from the FAQs and did not replace it with any info to the contrary. | -0.203 |
| 31-35 | Female | Non-training Fellow, SAS or other Trust-Grade post | Lack of clinical experience in candidates given an ST3 post compared to unsuccessful candidates. Lack of time to adequately boost score before the next application period | -0.203 |
| 31-35 | Male | Non-training Fellow, SAS or other Trust-Grade post | The anaesthetists in the UK have been working hard during the time of COVID, and after that what happened that there is a plan of curriculum change that will stop training recruitment for 1.5 years and it will make training application for ST4 more difficult for the overseas doctors and that’s not the thank you that I expected after working hard for the NHS during COVID time | -0.203 |
| 31-35 | Female | Anaesthetic Core Trainee | Over the last two years, core trainees have already been faced with the uncertainty of planned changes to the curriculum being delayed. As these changes are now due to come into effect next year (compulsory CT3 year for all core trainees), those current CT2s who find themselves without an ST3 post this august are left in limbo with no idea where they may re-enter the training programme and what curriculum at that time will be in effect | -0.202 |
| 31-35 | Female | Non-training Fellow, SAS or other Trust-Grade post | Lack of support from the college since leaving training. Paucity of training posts is disappointing considering the work we were required to do over the COVID crisis with redeployment and in light of the ‘catching up’ with deferred elective work. | -0.199 |
| 31-35 | Male | Intensive Care Medicine Trainee | The huge jump in competition and mismatch between applicants and posts available reflects poorly workforce planning. Anaesthetists are locked into anaesthetic training and as such a lack of registrar posts prevents career progression. | -0.197 |
| 31-35 | Female | Non-training Fellow, SAS or other Trust-Grade post | Concerned about finding employment from August onwards - I may have to move house and location completely. Also concerned that the criteria to enter ST4 are unclear, and I may not have certain things signed off, despite 5 years experience, which will delay my training even further | -0.197 |
| 36-40 | Female | Non-training Fellow, SAS or other Trust-Grade post | The interview was difficult. Trying to build rapport when the signal was bad. Waiting for them to unmute before asking the next question. Very difficult format. | -0.197 |
| 31-35 | Male | Anaesthetic Core Trainee | Already a significant bottleneck between core and higher training. Unless training numbers increase in line with predicted demand for anaesthetists over the next 10 years, there will be an ever increasing number of those who have gone through core training, out of a training post. Run through training would make more sense in the future - similar to emergency medicine and paediatrics. After such a busy year of work, with added stress of delayed exams, this year’s ST3 recruitment was just yet more stress when statistically it now seems as though I was unlikely to ever get a job. | -0.191 |
| 31-35 | Female | Anaesthetic Core Trainee | At the moment in the South West, there is a 10% deficit in Consultant staffing. Over the next 10 years this is projected to increase to 25% it hugely concerns me of the shortsightedness of the decision not to create many ST3 posts this year. Why demoralise a workforce who have already had a tough time with reassignment during Covid and the new curriculum, when in a few years time it’s likely these people will be CCTing and needed to fill the gaps in Consultant posts. | -0.191 |
| 31-35 | Female | Non-training Fellow, SAS or other Trust-Grade post | Uncertainty regarding jobs has an effect on family planning | -0.183 |
| 25-30 | Male | Non-training Fellow, SAS or other Trust-Grade post | I am deeply concerned that the current plan to forgo anaesthetic ST recruitment in august 2022 and February 2023 will worsen the bottle neck for recruitment of eligible trainees. Given that I aim to dual CCT I am now time limited to obtain an anaesthetic number and may have to delay training with an OOPE or part time working to weather this gap in recruitment. | -0.179 |
| 31-35 | Male | Anaesthetic Core Trainee | Annoyed that the primary FRCA requirement has resulted in some people being appointed ST3 jobs but not having gained the full Primary. Unfair that being in a core training programme there are not enough posts to follow through the commitment to specialty - either make enough posts for trainees or consider run through training like many other specialties. Frustrated at lack of recognition/support - now feel a bit lost in what to do for the next 2 years with relatively vague top-up competencies/don’t seem to be standardised. | -0.179 |
| 25-30 | Female | Anaesthetic Core Trainee | I was deemed appointable, I scored 86 when I needed 71 - I did everything that was required of me and more even with the pandemic, but not enough. I do not trust the college or ANRO or the interview process. I do not trust that I will not end up in exactly the same position next year. I am outraged that anaesthetists shielding who could stay at home and get through exams and create outstanding portfolios (BUT HAVE NOT SEEN A LARYNGOSCOPE IN 6 MONTHS) will be getting registrar posts over me. I’m furious. The census last year keeps telling us you need more anaesthetists and that the very real workforce gap approaching over the next decade from retirements and yet still they won’t recruit more people who want to be trainees. Why? | -0.177 |
| 31-35 | Female | Non-training Fellow, SAS or other Trust-Grade post | It’s unfair that so many didn’t get jobs. | -0.177 |
| 25-30 | Male | Anaesthetic Core Trainee | Stressful experience. It felt that core anaesthetic trainees with little prior anaesthetic experience were hindered more. | -0.177 |
| 31-35 | Female | Anaesthetic Core Trainee | Competition ratios | -0.177 |
| 25-30 | Female | Anaesthetic Core Trainee | Lack of career progression, lack of recognition for gaining FRCA exam in self score. | -0.174 |
| 31-35 | Male | Non-training Fellow, SAS or other Trust-Grade post | The design of training appears to be absolutely absurd. In Mersey alone >20 trainees enter core training every 6 months. 10 or less ST3 posts every round, and there is “surprise” that there is a shortage of registrar training posts each year. I have universally positive feedback clinically and professionally, have an adequate amount of CPD/Audit etc and have passed all of the exams, yet was deemed no suitable for interview on this round. I have completely lost faith in the training programme and am really disappointed. There is no guidance for any trainees from the deaneries, except a 6 monthly letter absolving themselves of blame and offering no long term solution. I would like to hear what the proposed plan for the 400+ trainees stuck in the middle of the two training programmes would be given the planned break for entry into higher training. | -0.173 |
| 31-35 | Female | Non-training Fellow, SAS or other Trust-Grade post | Such a low number of training places, especially since anaesthetists were in such high demand during the covid-19 surges and considering there will be a year with no st3 or st4 recruitment resulting in significant delays to career progression and advantages of those who only recently started training over those who are now between ct2 and st3 | -0.172 |
| 31-35 | Prefer not to say | Non-training Fellow, SAS or other Trust-Grade post | Very probable that many applicants will be leaving the profession due to highly stressful workloads particularly during the pandemic and poor job security trying to go through 1 year top up posts. Absolutely poor response by the government who has capitalised on the good will and sacrifices of trainees and non-training grade anaesthetists during the covid pandemic. The general consensus talking to other colleagues suggests working in the nhs is not worth the stress and effort. An employer who has no understanding of service requirements and how to retain hard working staff. | -0.172 |
| 25-30 | Female | Acute Care Common Stem (ACCS) - Anaesthetics Trainee | Overall I felt there was a general lack of consideration to the fact that most of had worked tirelessly on non-compliant rotas due to the fact there was a global pandemic. Then to just carry on as if nothing had happened and find out the number of jobs were slashed in a time where nobody was able to be competitive with their CVs due to not physically having any time between covid and doing the primary felt like we were massively unappreciated. And by wanting to take a career break in order to move deanery and pursue other goals I felt there was minimal information about how the changes to the curriculum would affect me including how to access top up posts, and the announcement for the Aug 2023 ST4 application date came after the deadline to apply for ST3. This should have been a time of assurance and appreciation rather than uncertainty and more stress. | -0.172 |
| 25-30 | Male | Acute Care Common Stem (ACCS) - Anaesthetics Trainee | I received my offer a few weeks after close. The competition was fierce and after a year of reduced training and candidates going above and beyond it felt incredibly harsh to have such reduced training numbers for the number of candidates. Particularly given the workforce shortage predicted over the coming years. Not only this but there’s a significant discrepancy in recruitment to the core training and specialty training perpetuating an inevitable bottle neck at the entry to ST3. Consideration should be made to making anaesthesia a run-through programme. | -0.171 |
| 31-35 | Female | Non-training Fellow, SAS or other Trust-Grade post | Lack of training posts despite countless rota gaps. Change in curriculum so close to the covid pandemic which already impacted training. Lack of support or guidance for CT3 top up - including finding these posts No recruitment for a year - further worsening the bottleneck into ST Lead to poor morale amongst trainees who have worked so hard | -0.17 |
| 25-30 | Male | Acute Care Common Stem (ACCS) - Anaesthetics Trainee | It’s utterly infuriating. What is the point in training people to ct2 level if there are not enough st3 jobs? I am absolutely not prepared to move across the country for the next stage of my training (given house ownership, partner’s medical speciality training, childcare etc) and so I would not be surprised if I end up changing careers, despite Anaesthesia being the career I love and have already committed numerous years to my anaesthetic training. The “flip-flop” of “will they/won’t they?” with the introduction of new curriculum was handled appallingly with little to no communication from the top - again affecting life plans, job plans, and long term family plans for anaesthetists in training and their families. It is a matter of huge concern and stress. There is a feeling amongst core trainees that our working life, and its effects on us, is going to get a lot harder now that essentially no-one from our cohort has got an st3 job, and we are all likely to be out of training posts for at least 18 months. It is galling. | -0.168 |
| 31-35 | Female | Non-training Fellow, SAS or other Trust-Grade post | I was released from training in 2019 after ST4 as I didn’t have my final FRCA. With the change in curriculum I am worried that I will struggle to get back into training due to the change of intermediate trading from ST3&4 to ST4&5 | -0.165 |
| 36-40 | Male | Non-training Fellow, SAS or other Trust-Grade post | The ST3 recruitment process this year has been, in my opinion, the biggest failure of workforce planning and educational reform since MMC/MTAS. After diligently propping up surge ICUs, and caring for critically-ill COVID patients, the NHS has treated anaesthetists-in-training as single-use, disposable, items - burning them out on surge rosters when needed, and then casually discarding them, and leaving them without jobs or careers, when not. The failure of RCoA to care for its own trainees is a particular disgrace. The organisation - which is happy to preach accountability, responsibility, and transparency to those working clinically - has fundamentally failed to take any responsibility for this mess, via its botched curriculum introduction. Instead, it has disingenuously pointed the finger at others (specifically, HEE and GMC), rather than reflecting, apologising, and trying to put things right - as it should. The whole mess is so toxic, and casts the organisations involved in such a poor light, that it cemented my decision to permanently leave the NHS and the UK. | -0.163 |
| 31-35 | Male | Acute Care Common Stem (ACCS) - Anaesthetics Trainee | The application process was appalling and the feedback frankly insulting and demeaning. For example I proudly had a poster presented at an international meeting which I had designed and was a major part of the team. The names were placed alphabetically, which meant my name was 5th. My name was on the programme. I got zero points for this and told â€œ 5th author on a poster doesn’t merit any pointsâ€ . I was unable to appeal this as it didn’t meet the threshold. Frankly this is disgraceful and insulting. | -0.163 |
| 31-35 | Male | Non-training Fellow, SAS or other Trust-Grade post | I think the application process actively discriminates applicants with caring or childcare responsibilities who don’t have the time or money to play the points game to gain an interview. This is something I have witnesses first hand, I have seen colleagues with children fail to get a job, and colleagues who are carers leave Anaesthetics. I appreciate that there has to be a method that allows for the limited (which is another problem) number of posts to be given out, but the current method is discriminatory. If ANRO insist on this method then each point needs to be reviewed for its impact. From what I have read SJT type application would be more fair. I also find it’s mind boggling that the UK tax payer has invested so much into my training at this point and the UK is real need for more Anaesthetic/ICU consultants but there is an artificial road block halting my training. The UK will loose their investment if this continues. | -0.162 |
| 25-30 | Female | Acute Care Common Stem (ACCS) - Anaesthetics Trainee | Large bottleneck of applicants and not enough posts available.. Concerned as someone who has passed the exam that this was no longer a requirement. I feel that I’ve given all my time to my career and the covid pandemic in the last 18 months only to be overlooked as a credible ST3 colleague due to shortfalls in the interview process and planning for what a change in the curriculum might look like as we are coming out of a pandemic. | -0.16 |
| 31-35 | Female | Non-training Fellow, SAS or other Trust-Grade post | I personally feel there were more issues with the previous years recruitment. This year was not particularly different to the standard RCOA recruitment process (albeit with an online rather than in person interview), it was simply more competative. I applied for the August 2020 ST3 start and was not successful. I thought it was incredibly unfair that jobs were allocated purely based on an un-verified self assessment score. | -0.158 |
| 31-35 | Female | Non-training Fellow, SAS or other Trust-Grade post | There is a serious need for trainees , yet there is no increase in the training posts. Its very unfair. It should not be so competitive to get a post of your choice and in a place of your choice. I am seriously thinking of other speciality options that too after 9 yrs of experience in Anaesthesia as I feel this competition is affecting my life and my mental health. | -0.156 |
| 25-30 | Male | Acute Care Common Stem (ACCS) - Anaesthetics Trainee | This has been mismanaged from the start - trainees were not told they were going to change curriculums when initially accepting SHO posts (2018 ACCS Start). Then they were told the change in curriculum would be accompanied by guaranteed CT3 posts. Then the curriculum change was delayed and this took away the CT3 posts from the 2019 Novices. If the curriculum start can be moved, why can’t this employment support? Why can’t there be an ST4 entry in August 2022 meaning no time has to be ‘wasted’ for those unable to get into an ST3 post this year? | -0.155 |
| 31-35 | Female | Anaesthetic Core Trainee | People are lost in the system and the deaneries are not interested. Trainees are broken due to covid and as a thank you gift they are kicked out of the training programme. | -0.152 |
| 25-30 | Female | Anaesthetic Core Trainee | lots of uncertainties & anxiety, lost opportunities and life plan changes. All leading to burn out. | -0.152 |
| 31-35 | Female | Non-training Fellow, SAS or other Trust-Grade post | Lack of priority for those who have already completed core/ACCS training or have the primary exam. Lack of priority for those who missed out on training numbers in August 2020 due to unfair non verified self assessment scoring. Prolonged application window increasing period of stress across an already stressful year. Lack of feedback for online portfolio or interviews. Lack of guarantee regarding increased numbers for ST4 application in 2023, at which point there will be competition from those of us forced out of training for two years, in addition to those coming to the end of CT3 core/CT4 ACCS. Almost feels like a conspiracy to increase the number of SAS doctors (which we know are significantly lacking). | -0.15 |
| 31-35 | Male | Non-training Fellow, SAS or other Trust-Grade post | The delay in reapplying and likely very high competition means the route to consultancy is too long. Training in GP will provide quicker access to CCT and I can continue anaesthetising as a locum or SAS doctor without the need for years of hospital rotations and night shifts etc. | -0.15 |
| 25-30 | Female | Non-training Fellow, SAS or other Trust-Grade post | Concerns with the portfolio system. Marks given were inconsistent between application rounds for the same evidence (this was my third application but my first to get an interview and the same evidence got different points on different rounds) and were also inconsistent between candidates for the same evidence. Colleagues who took part in the same project as me and submitted the same evidence were not awarded the points. Given the obvious subjectivity of this I would like to see an adequate standardisation and appeals process, however you are not allowed to appeal if there is less than a 5 point discrepancy. With competition so tight one point can make all the difference between getting a job or not so seems unfair. In addition there was inadequate guidance about the portfolio, for example I lost points for not including a CV as this was apparently required to ‘prove’ the duration of my additional degree and for the quality of my overall portfolio. However in the guidance they did give us they did not tell us to include a CV but did tell us to avoid any unnecessary documentation so many people kept additional documents to a minimum as directed. Also the applicants significantly outnumbering the number of posts has been a problem for several years now, are we still recruiting the same number of CT1s now because that would also seem unfair if we know their won’t be jobs for them. | -0.149 |
| 25-30 | Female | Anaesthetic Core Trainee | Very rigid system which cannot be adapted. I had to apply in December for a post starting in August and our lives look very different now. Not sure why you cannot apply to all the regions in the UK at least at the beginning. Delays in publishing numbers of jobs and rotations. Hold deadlines in the middle of May to start in august give people very little time to sort out things if they are moving regions. Not a standardised way of scoring portfolios and irritating that 5 points was the cut off for appeals- even 1 point makes a huge difference when things are this competitive! | -0.148 |
| 25-30 | Male | Non-training Fellow, SAS or other Trust-Grade post | I found the implementation of the criteria for portfolio scores to be reviewed was not well applied. The original submitted score I submitted was lower because I had yet to gain more points by the time of evidence submission on the online portal. By this time I would have been deserving of more points. Unfortunately evidence from a whole domain on my portfolio was missed by the verifiers and the responses I was receiving from ANRO were unsupportive, showed little nuance and understanding of the situation I found myself. It was in short: a ‘computer says no’ response; “we just apply the process.” I was not allowed to appeal because the difference between my original submitted score and verification score was <5 points. I had lost 4 points through missed evidence and I wished to contest at least 2 other points from other domains. I informed my educational supervisor and college tutor but they were powerless. I took it upon myself to contact the chair of RAG recruitment, who very kindly informed me that I might still be able to retrieve the points lost by human error by submitting a complaint. The ANRO instructions and my correspondence with them did not make it clear that the complaint process was a way of notifying them about appealing for scoring miscalculations due to human error. Given that the process is very human based, this was an an avoidable scenario. I am disappointed that it required contacting the RAG chair directly and seeking advice on anaesthetic Twitter to “try my luck” to retrieve points. | -0.144 |
| 36-40 | Male | LAT ST3 | The oversubscription is a predictable problem and it is only going to get worse. I feel the move to allow applicants to apply without full Primary FRCA was a mistake. Even if this has to be allowed, applicants who have achieved full Primary FRCA should have been given priority or additional points in self assessment. This change has meant an increase in number of applicants who would otherwise not have been eligible, hence inflating the competition ratio. This also means that applicants who have not achieved full Primary FRCA may now move on to an ST training post, with no guarantee that they will be successful in passing the OSCE/SOE in the coming year. By the end of 2021-22 training year there will certainly be a group of ST3s who failed to achieve Primary FRCA but are in a training job, and a group of LAT/LAS/CDF who have already achieved the basic requirements but are out of a training job. This is unacceptable. I have been unsuccessful two years in a row and I have worked hard to improve my CV to improve my chances this year but unfortunately still unsuccessful. Based on all the feedback that I have got, and the fact that I have passed the full Primary FRCA since 2019, I am confident that I am a trainee that my consultants would vouch for, and that I do deserve to train in this specialty. Having been unsuccessful twice now, I see this as a failure of the recruitment system and I am not sure how much more I could do to mould myself to fit into this failed system in order to be successful in my future applications. | -0.143 |
| 31-35 | Male | MTI Fellow | Lot of competition . As an MTI and new to country , found it hard to complete necessary curriculum especially in ICU as I was unaware | -0.143 |
| 25-30 | Female | Anaesthetic Core Trainee | the change in curriculum plans and lack fo clarity for the last 2 years, with the lack of opportunity to apply for st4 in 2022 closing a number of doors | -0.142 |
| 31-35 | Female | Anaesthetic Core Trainee | I was fortunate this year to get a ST3 post, however, many who are as qualified (or more qualified) did not and it seems ridiculous where there are acknowledged shortages and projected increasing shortfalls of anaesthetic staff that there are not training posts for those who want to continue training. This situation is more frustrating in that it was entirely predictable and the response from the college would have been much more useful to trainees several months ago. Throughout the last year, from when the decision to remove the CT3 year was made (in the middle of the pandemic) there has been a lack of communication about recruitment and how those missing out on posts would be able to proceed with training. This has added a huge amount of stress in an already very difficult time. Speaking to colleagues, many individuals feel totally undervalued, demotivated and fed-up with the huge impact on their professional and personal lives. | -0.141 |
| 31-35 | Male | Non-training Fellow, SAS or other Trust-Grade post | The questions at interview seemed very generic, with little way of really showing your individual strengths and weaknesses. My guess is that scores at interview were very similar, with a very narrow bell curve. If this is indeed the case, then it is not acting as a good discriminator for the role. | -0.137 |
| 31-35 | Male | Non-training Fellow, SAS or other Trust-Grade post | Pushing through the curriculum change during the COVID pandemic has been detrimental to applicant’s/trainee’s mental health and career prospects. Due to the pandemic those who would usually have gone abroad can not, those who were abroad have returned, and due to the curriculum change many have seen this as a “last chance” to get an ST3 number all together leading to the large number of applications and subsequent stress and disappointment. For those not getting a number this year, it is a mandatory two years out of training forcing them to undergo further application and interview cycles, both nationally and locally - potentially four over a two and a half year cycle, including the application round just gone. The provision of CT3 “top up” posts has not been centrally co-ordinated or offered equitably, being down to hospital trusts to organise and fund meaning there is a ‘postcode lottery’ of sorts to obtain these. I am recently engaged, and my partner has had an ST post for the last year that she cannot move from. We would not qualify to apply for an inter-deanery transfer unless she was to give birth due to the restrictions on these. There is no provision of top up posts in a commutable distance from where she works, even hours away. In my mid-thirties and starting a family, it is not practical to move across the country for a top up post. Locally trusts are not recruiting many fellow posts, presumably because their existing core trainees are now staying on and filling those rota slots. I finished Core Training during 2020 and the first wave of the pandemic, despite redeployment to ICU and missing some training. I stayed in a local fellow post largely due to uncertainty during the pandemic and a wish to help out. I am now at a loss as to what to do with my career and have limited options, and am subsequently considering leaving the profession altogether. I feel abandoned by the NHS, HEE and the RCoA. | -0.137 |
| 25-30 | Male | Anaesthetic Core Trainee | Allowing trainees without the Primary exam to apply and start ST3 in the same year as a shambolic curriculum change in the middle of a pandemic was always going to result in an enormously oversubscribed programme. The situation was totally foreseeable and nothing was done to mitigate it. Without prior warning, the weighting of interview to portfolio was dramatically changed compared to usual and this gives unfair advantage to candidates who have had multiple career breaks and changes - hardly selecting for candidates who have had a long-standing commitment to anaesthesia. CT2s are now left in limbo, scrambling for the few CT3-compliant jobs available for fear of being stuck indefinitely in this no-man’s-land. Finding short-term accommodation and forcing partners and families into difficult decisions about their own life-plans in the interim is grossly unfair and immensely stressful in the middle of a global pandemic. To make matters worse, the next ST4 intake is not due to commence until the current CT1s are eligible - again creating a horrendously competitive recruitment round where history can repeat itself. It really is beyond belief that trainees who have been integral to the pandemic response and worked tirelessly throughout are being treated in this way. | -0.137 |
| 31-35 | Prefer not to say | Non-training Fellow, SAS or other Trust-Grade post | Extremely drawn-out process starting in November 2020, spanning 2nd wave of COVID. Constant rumours circulating that there would not be enough posts (correct) and that competition would be unreasonably tough (also correct). Addressed with tone-deaf platitudes by RCoA, who should/could have seen this coming a mile off (all the trainees did). Unbelievably disrespectful to the hundreds of deserving trainees who have given their physical, mental, and emotional health over the past 18 months. I was one of the lucky ones to get a post and will be selling my house and moving my family to facilitate it as I don’t want to have to wait two years to go through this again. Already feeling survivor guilt as there were many far more deserving of appointment than me. | -0.136 |
| 31-35 | Female | Anaesthetic Core Trainee | Very demoralising and stressful. Had to compromise staying geographically close to my loved ones purely just to get a training place. Many colleagues didn’t receive spaces. High competition ratio. Everyone already feeling burnt out from covid escalation. We had sat exams and passed in the middle of a surge - despite this they weren’t counted so the completion ratio was even higher. | -0.136 |
| 25-30 | Male | Anaesthetic Core Trainee | Poor, reactive comms from RCoA/HEE about changes to applications, including dates/curriculum changes/job availability etc. Online portfolio system was inadequate and explanation video provided by College contradictory. Seemingly little consideration of effects of all these last-minute changes on trainees. | -0.132 |
| 25-30 | Male | Not currently in contracted employment, or currently a locum | Verified self assessment scores differed in the recruitment rounds completed in Feb ’21 and Aug ’21, despite uploading exactly the same evidence for both recruitment rounds. No opportunity to appeal this as score was <5 required. | -0.132 |
| 25-30 | Female | Acute Care Common Stem (ACCS) - Anaesthetics Trainee | Extremely competitive application process with apparently very little to discriminate between top scoring candidates and those who did not receive a job. Stressful process which exacerbated feelings of burnout leftover from pandemic | -0.131 |
| 25-30 | Female | Anaesthetic Core Trainee | Generally stressful application process due to uncertainty of number of posts, the stress about potential 2 year gap and having to find CT3 equivalent posts. Late guidance on the top up jobs adding to stress | -0.13 |
| 25-30 | Female | Non-training Fellow, SAS or other Trust-Grade post | I am concerned it will leave me out of a training post for a prolonged time. Competition for any posts that offer CT3 competencies (particularly paediatrics) will be high. | -0.127 |
| 31-35 | Male | Acute Care Common Stem (ACCS) - Anaesthetics Trainee | Very poor communication from HEE and RCoA. Feel left out in the cold especially with a new curriculum change and delayed entry and ST training. Has really made me think twice about continuing to pursue anaesthetics training. | -0.127 |
| 36-40 | Male | Non-training Fellow, SAS or other Trust-Grade post | Opening it up to people with the MCQ only is not fair on people who worked really hard to pass the osce/viva and have the full exam. | -0.123 |
| 25-30 | Male | Non-training Fellow, SAS or other Trust-Grade post | It was extremely obvious to everyone in anaesthetics that this would occur except it seems the RCOA and HEE. The timing of the new curriculum has left us stranded. Despite passing the exam and completing all that was asked in a difficult time there have been no rewards for this. This has made me strongly consider not becoming a consultant. South yorkshire has also become pandeanery in the past few years. I refuse to uproot my family to work as a consultant in a profession that does not reward hard work. We all should have been welcomed with open arms into training as there are shortages across the region in the consultant body and at spr level. Local hospitals are plugging the gap with non training roles. Mine now has 25% of the department staff as fellows or speciality doctors. Being good clinically is not rewarded, those who progress are the ones that spend every waking hour on audits, research and teaching which does not translate into being a good consultant. | -0.121 |
| 25-30 | Male | Acute Care Common Stem (ACCS) - Anaesthetics Trainee | Not enough info of support. Feel abandoned and left. Especially after covid the curriculum change should’ve been delayed. And a whole cohort should’nt have been left to find top up years. | -0.121 |
| 25-30 | Female | Non-training Fellow, SAS or other Trust-Grade post | Inconsistent scoring of points by evidence assessors caused much stress, inability to appeal scores was horrible as I know that other specialty recruitment allow appeals of even disputes of 1 point - I felt I might have a job slip through my fingers because of an assessor’s misunderstanding. Last minute changes to recruitment timelines meant long-term goals (such as exams, projects) were missed off of self-assessment scores with new timelines - I felt like the goal posts were being moved without warning. Having my achievements (for example, passing the full Primary FRCA) mean nothing this round of recruitment felt very harsh given how hard I had worked during CT2 to get them completed. All in all, I felt like RCoA/HEE cared nothing for my wellbeing as a trainee trying to get back in to training after missing out last year due to my honesty in self-assessment scores that ended up negating my chances in 2020 recruitment. | -0.116 |
| 31-35 | Female | Non-training Fellow, SAS or other Trust-Grade post | I have great concerns about ST4 recruitment 2023 - I will be one of those applying with 7 years of experience under my belt, alongside current novices, CT2s and CT3s who will by then be eligible. The numbers looks even worse than it was this year. Might a separate recruitment process into ST5 in 2023 be a solution, so long as it had strict eligibility criteria? I have further concerns about the logistics that fellows now face when trying to evidence experience - LLP hasn’t been updated to the new curriculum, so there isn’t currently a way to properly log stage 1 top-up competencies or further competencies for those considering the cesr route. | -0.114 |
| 25-30 | Female | Anaesthetic Core Trainee | A lot of things to try and achieve which was difficult with working the pandemic. The system feels like it favours those with more experience and potentially time out of training to boost their cvs. Difficult as it has become more and more difficult year on year to apply for st3 with no increase in numbers. | -0.114 |
| 31-35 | Female | Non-training Fellow, SAS or other Trust-Grade post | It constantly feels like the goal post is being moved and these changes are very poorly communicated. I came out of training to finish the primary exam, this was delayed because of covid, I have then been out of training too long to be eligible to sit it in the first few rounds so was pleased when this criteria was removed from applications but why is my additional experience not counted? Now I have spent years trying to get into the next stage of training and I cannot keep putting my life on hold as it effects my family too. If you are happy as a specialty to let me run trauma calls and be senior on call why can’t I have some benefit from official progression? | -0.114 |
| 31-35 | Female | Non-training Fellow, SAS or other Trust-Grade post | I’m hugely dissatisfied with the current situation and feel saddened that I’m facing leaving a profession I love and something I’m clinically good at. I think that some of this comes down to bad timing and choosing to have a family whilst in training but for a long time I’ve also had this feeling that being from a working class background where I couldn’t afford additional degrees and time out or foreign electives will continue to disadvantage me for the rest of my career and it’s hard to un-do that. I think the failure of the 2 year core training programme is being carried by myself and those who entered that training at the same time as me and now I’m stuck without job security, in full time short term contracts with no real light at the end of the tunnel and I’m having to reconsider how long I can put my family and myself through this. | -0.113 |
| 31-35 | Female | Intensive Care Medicine Trainee | The self assesment scoring was vague, it was very marker dependent. People had national presentations counted if accepted but hadn’t presented yet, others didn’t. The guidance states will no longer take into account courses not being done but having had over 3 of my ATLS courses cancelled, 1 the night before due in. The face to face courses not at same rate. Interview - limited information prior. | -0.111 |
| 25-30 | Female | Anaesthetic Core Trainee | Overwhelming competition for trust grade jobs and future st4 application | -0.111 |
| 31-35 | Male | Non-training Fellow, SAS or other Trust-Grade post | The self assessment section appears extremely unreflective of the job of an ST3 it is very unfair. It also significantly favours graduates who have had a previous career or research background prior to medicine. | -0.109 |
| 25-30 | Male | Anaesthetic Core Trainee | It genuinely felt like the competition this year was way too high - given the key role that anaesthetics trainees had on ICU in dealing with the COVID pandemic, the significant impact this had on our training/well-being, and the key role future consultants in anaesthetics will have in clearing the vast backlog of surgical cases over the many coming years, it seemed ridiculous to me that places were so limited. It makes sense to expand the number of anaesthetics trainees, or just make it run through (why does there even need to be an ST3 recruitment round?). I feel like I only got a job because I was able to apply to every region. Those with children/families/other commitments who cannot work outside of the region they currently live were significantly disadvantaged. This seemed unfair to me. | -0.104 |
| 25-30 | Male | Anaesthetic Core Trainee | why so few jobs? There is shortage of consultants, and registrars are needed to fill the rotas | -0.104 |
| 25-30 | Female | Anaesthetic Core Trainee | Enough experience given COVID affected training | -0.102 |
| 25-30 | Male | Non-training Fellow, SAS or other Trust-Grade post | My portfolio score was not strong but with the pandemic I have had only limited opportunities to improve upon it, leaving me without an interview and dispirited as a consequence. I am now facing only a single chance, likely highly competitive, to obtain a training post before facing a prolonged period with no further chance to apply for anaesthetic training. I don’t know how these curriculum changes are likely to plan out and if I do obtain an ST3 ICM post in august 2022 (I was also unsuccessful in this application round) I don’t know what to do if I fall out of sync with the anaesthetic programme. I have found the whole application process very stressful and depressing at a time when my clinical workload was both highly demanding and at times very depressing working in a busy tertiary ICU. The rejections from both programmes this year has left me feeling despondent and anxious. I am not sure what I would have done had my current department been so supportive. | -0.102 |
| 31-35 | Male | Non-training Fellow, SAS or other Trust-Grade post | The number of applicants applying for st3 (or st4 soon) will keep increasing above the number of posts, as there seems to be so many core training numbers still available. This is going to make it harder to get any registrar post in the future. This is all on the background of a shortage of Anaesthetic consultants. I worry that the government is trying to cheapen anaesthesia and increase the number of Anaesthetic associates and eventually make anaesthesia not a physician led speciality. | -0.101 |
| 31-35 | Male | Non-training Fellow, SAS or other Trust-Grade post | That the situation will not improve - applicant numbers are only going to increase and if they don’t increase the number of ST3/4 posts, this is going to keep happening. I also have concerns that online interviews with one panel are not the fairest way of selecting candidates. I was forced to take time out of training following CT2, due to personal circumstances, and now I feel further trapped in circumstances beyond my control. However, as long as the new curriculum can be fulfilled without a training number, people will just find their own way to consultancy. Otherwise I fear lots may leave anaesthesia altogether. | -0.099 |
| 31-35 | Male | Non-training Fellow, SAS or other Trust-Grade post | This recruitment process has been an absolute shambles. The people involved in workforce planning clearly have no clue what they are doing. Hundreds of trainees are now without ST3 posts and it seems logical that the next few rounds of applications will also be massively oversubscribed. This puts our lives in a lot of uncertainty and is a slap in the face after the all the help and sacrifice these anaesthetic trainees made in order to help their hospitals cope during the pandemic. We are treated as a number by the NHS and we are not valued as skilled employees. The selection process itself is a joke as the portfolio scoring system is just a tick box exercise with no real value. The rigid rules governing selection and even around those who wish to return to training after previously leaving the training programme are completely arbitrary and lack any common sense. The new curriculum changes by the college just add to the uncertainty that we are facing. The fact that we can be treated this way and just have to accept it, combined with the utter lack of planning and common sense shown in the recruitment process, makes me seriously consider whether this is a specialty I want to be a part of and makes me consider leaving medicine altogether. | -0.097 |
| 36-40 | Male | Non-training Fellow, SAS or other Trust-Grade post | Its very dissapointing after working so hard and unable to get a training post specially in a feild you need more and more specialist in the future. | -0.096 |
| 25-30 | Female | Anaesthetic Core Trainee | As a cohort I feel that the CT2 group have been really messed around by RCOA and this st3 recruitment round demonstrated this perfectly. Upon starting our CT1 year we were told we would have a 3 year training programme due to the curriculum change, this was changed at last minute in the middle of the pandemic. We were then told we would have to pass our exams within 2 years of ct not three and after passing the mcq not allowed to sit our viva in the most convenient setting due to backlog. We then had Reg applications moved forward significantly with no notice. I think that the relatively low numbers of ct2s going directly into training from st3 demonstrates how disregarded by the college we have been. I also feel the recruitment process for st3 was unnecessarily vague and there was no reason to so heavily weight portfolio when a clinical station would have been easily feasible as demonstrated in the online viva. Those who have been working covid rotas will inevitably have fewer audits/research but will probably be clinically more experienced. The message coming from the college is that “all will be fine” but there has been very little in the way of organised LAT/CT3 competency jobs. At the end of the day we have mortgages to pay and we can’t rely on promises which, going by their track record, are likely to be empty. This whole episode seems like a slap in the face after we stepped up and cross covered in horrendous itu conditions. Having jumped through all of the changing hoops mid pandemic to then be told we are unemployed is disgusting | -0.092 |
| 25-30 | Male | Acute Care Common Stem (ACCS) - Anaesthetics Trainee | Why were we allowed in to core training if this was going to happen at specialty training. It should come as no surprise that anaesthetic SHOs will attempt to become anaesthetic registrars after investing 2 to 3 years of their life in the training programme. So how did 2/3rds (the majority of applicants) not make it into anaesthetic ST3 posts?! It’s appaling how we have been treated this year and the change in curriculum after a generation defining pandemic is surely Ill conceived! I would also like to know is this representative of the amount of jobs available at the end of training? In which case should I leave anaesthetics now as there will be no job for me when I finish? Furthermore what is being done about applying again in 2 years. Surely the same problem will arise unless training numbers are increased?! What is HEE or the college going to do about that? | -0.088 |
| 31-35 | Male | Non-training Fellow, SAS or other Trust-Grade post | I’ve been really disappointed by the application and appointment process since covid has struck. I’ve not been able to get an interview for 18 months despite having the primary exam before finishing my ct2. I’ve worked hard and had good feedback from my colleagues. The measures of being a good candidate have no relation to whether a candidate is able to do their job or not. The college say they care about us and are trying to keep the process fair. However, I have been disadvantaged by being a February starter and missing out on the application window by 2 weeks. Since then I have not been deemed a better candidate than colleagues despite them not having the primary frca. I’m fed up and tired. | -0.087 |
| 25-30 | Female | Anaesthetic Core Trainee | I found this round of recruitment to be incredibly unfair. I passed the primary in January of this year (all components first time), and yet because my self assessment score was not deemed good enough I have not been offered an ST3 post when some who have not passed the primary have. I do not see how the self assessment score at all reflects my abilities as an anaesthetist and why it should have denied me the opportunity of an interview. I also find it appalling that trainees in my year will not be offered a CT3 top up year like those in the year below us but will have to sort something out for ourselves. Not only this but if we are unsuccessful at securing ST3 posts again in February 2022 we will be forced to take another year out. I think the curriculum change has been poorly thought through and has left many like myself in limbo. It makes us feel undervalued, especially after a year stepping into ITU surge rotas, and has diminished enthusiasm for the specialty significantly. | -0.086 |
| 25-30 | Female | Anaesthetic Core Trainee | Due to threat of being with no training job for 2 years, I felt I had to apply and accept post. It’s particularly discriminatory to women wanting to start a family, who then face uncertainty for 2 years. | -0.084 |
| 25-30 | Male | Anaesthetic Core Trainee | The communication from the RCOA has been awful throughout this entire process. They helped create an atmosphere of panic by giving no information about transition plans or what would happen to those who didn’t get an ST3 number. Their subsequent refusal to accept any responsibility afterwards is also pathetic. The decision to force through changes in the curriculum during a pandemic is also extremely questionable. I understand it is GMC mandated but did they really need to change the length of core training? I also refuse to accept that they didn’t see this mismatch in job applications to places coming as a result of the change since every single trainee knew it was coming. The entire situation is farcical and has damaged the reputation of Anaesthesia in the UK for an entire generation of doctors. | -0.084 |
| 25-30 | Male | Non-training Fellow, SAS or other Trust-Grade post | Inconsistencies in self assessment verification marking | -0.082 |
| 31-35 | Male | Non-training Fellow, SAS or other Trust-Grade post | The key concern for me is the lack of training post availability more than the curriculum change. It is nonsensical to take trainees on through core training and not have enough speciality training posts for them all, especially given the shortages seen in most hospitals on both the registrar and consultant rotas. This problem is longer standing than simply the impact of the new curriculum, and has to change. More speciality training posts must be made available through HEE in order to ensure that the efforts to train CT’s are not wasted and the anaesthetic workforce continues to be adequately staffed into the future. | -0.081 |
| 25-30 | Male | Anaesthetic Core Trainee | More effort should have been made to anticipate the huge increase in applicants because of the College’s decision to press on with the updated curriculum and plan accordingly e.g. secure additional posts for ST3 Applicants. Communication regarding the use of the online portfolio evidence system was poor. The application portal itself could have been better designed - e.g. A CV was supposed to be uploaded in the Undergrads Degrees section, despite this not actually being evidence of any undergraduate degrees, and candidates were marked down for not uploading a CV here. Given the impact of covid on existing rotas, which in my personal experience are often understaffed, plus the anticipated large backlog of elective surgery that must be addressed, I feel the college and Health Education authorities should have done more to secure additional places for ST3 applicants. Overall the experience has contributed to my disillusionment with the NHS and the medical training careers system as a whole, made my and my colleagues skills and experience feel unappreciated, and further validated my personal decision to seek further work abroad. | -0.081 |
| 31-35 | Male | Acute Care Common Stem (ACCS) - Anaesthetics Trainee | I found the changing numbers of posts across the application process particularly stressful. I felt these should have been published early and been consistent. | -0.079 |
| 31-35 | Female | Non-training Fellow, SAS or other Trust-Grade post | There is a national shortage of doctors. Anaesthetists have sacrificed training opportunities during this pandemic to assist in ITU. There is now a large number of competent anaesthetists who will not be employed causing a huge bottleneck in the system. Doctors are constantly being forced to move around the country. It’s a slap in the face to trainees. | -0.079 |
| 25-30 | Female | Intensive Care Medicine Trainee | I am completely devastated disheartened by anaesthesia and the RCOAS response to recruitment this year. Recruitment to dual training is so challenging it’s like they are forcing us to choose one or the other when both compliment each other perfectly | -0.077 |
| 31-35 | Male | Intensive Care Medicine Trainee | It was a positive that the self assessment was verified this year. However I feel that the interview questions poorly discern between candidates ability - anyone can talk about a critical incident or why they want to do anaesthetics but it would be more testing to discuss a clinical scenario and/or prepare a short presentation whilst in the “Waiting Room” (both of these still possible via teleconference). | -0.076 |
| 25-30 | Male | Anaesthetic Core Trainee | Feb 22 intake will be even more competitive. There were ~550 CT1 posts this recruitment round leaving a 200 job deficit at ST3 level for 3 years’ time, representing a waste of training resources and a waste of trainees’ time. There is a shortage of registars to staff middle grade rotas which post-CT2 anaesthetists will largely fill in fellow posts, so the NHS pays at least as much and probably more when the likley increase in locum shifts is factored in. In addition there is a shortage of consultant anaesthetists. Why then is NHS England not funding more anaesthetics registrars and instead choosing to exacerbate the problem? | -0.076 |
| 25-30 | Female | Acute Care Common Stem (ACCS) - Anaesthetics Trainee | The whole thing was poor from the start. The change in recruitment timeline was informed to us by Oriel, not the college. Lack of communication and transparency, when we have all been taught since medical school that patient complaints are as a result of poor communication. The application process itself was ok. We were informed about the process through RCoA videos. I think if these were more advertised, this may have helped some people. The document upload was fair for me. The interview process is beyond poor. The interviewers have no access to our portfolios, they do not know anything about us and they have fixed questions with a fixed time limit to find out about us. Some of my colleagues were unable to finish their answer to a question as interviewers interrupted and moved them on. This is not an interview, not for a training job. It is disrespectful. | -0.075 |
| 31-35 | Female | Acute Care Common Stem (ACCS) - Anaesthetics Trainee | I had a 1 yo during the time of the pandemic, nursery closed even to NHS workers, mine and my husband’s family live in Rep of Ireland so not able to help. Somehow (and I honestly have no idea how we did it) we managed to juggle my full time awful surge rota with the demands of a small child. My husband taught full time with our 1yo on his lap for the first 3 months of Covid. In the midst of all of this we had a leak in our house and had to move out. We made it through and I feel so fortunate that we didn’t lose any loved ones etc but we are both so burnt out. I was on a surge rota for a year until March 2021. I was very conscious that my portfolio could have been a lot better for the ST3 interviews but I could not have physically done it. I am so exhausted. I don’t know where I would have had the time to do extra projects, run teaching sessions etc. During the interview when they asked me about the most difficult period of my training I just wanted to say ‘you honestly have no idea.’ I am not a dramatic type at all and tend to just get on with stuff but honestly after the amount of sacrifice not to be given a training job this year feels beyond insulting. I will do my non-training post and then see where I get to but I think my husband and I will probably take our now 2yo to Rep of Ireland and find work there. The Irish health system has its faults of course but I don’t think I could find it in me to go back into a training scheme run by the faceless HEE. They absolutely don’t care for trainees and I honestly want nothing more to do with the training system in the UK, at least this is how I feel at the moment. | -0.075 |
| 25-30 | Female | Non-training Fellow, SAS or other Trust-Grade post | I was offered a job out of region and so am relocating with two small children. Whilst this is a big undertaking and a massive upheaval to our lives I feel the current situation warrants it. | -0.073 |
| 31-35 | Male | Non-training Fellow, SAS or other Trust-Grade post | I felt let down by the entire process. Despite putting in massive efforts, working through the pandemic and building a strong portfolio at the same time, clearing the full Primary FRCA, and the interviewers commenting â€œsolid candidateâ€, I felt a 30 minute interview was not good enough to gauge the full merit of a candidate, especially when there were so many candidates presumably with very similar scores and are more or less at a same level. I felt I truly deserved a post and I am disappointed that I have been denied one. | -0.072 |
| 25-30 | Female | Non-training Fellow, SAS or other Trust-Grade post | I have put aside big life decisions waiting on the geographical stability of an ST3 post. The pandemic, the change in the curriculum and the years long terrible workforce planning meant that I, and hundreds of others, are left hanging. The changes have been rushed through with very little thoughts spared for so many of us. I feel angry and let down by the College who has failed to advocate for us. After a very challenging 12 months we deserve better. | -0.07 |
| 36-40 | Male | Non-training Fellow, SAS or other Trust-Grade post | Selection heavily favours those who have taken additional years out of training to gain points. Couple this with me being unwilling to move house as I am 37, have a family and a home I’m planning to stay in for decades, I feel I was incredibly lucky to gain a suitable training post. Others just as capable were not so lucky. | -0.069 |
| 31-35 | Female | Non-training Fellow, SAS or other Trust-Grade post | I dont know what more I can do to obtain a post: I have received good feedback and maximised my portfolio. All round frustrated. | -0.069 |
| 25-30 | Male | Acute Care Common Stem (ACCS) - Anaesthetics Trainee | My wife and I are both in anaesthetic trainees in the same situation. We had planned on moving region for ST to be closer to family and plan our future, as we only married in May. A training post would have given us some certainty. I’m concerned we may not be able to get a training post in the same region due to the competition. | -0.068 |
| 25-30 | Male | Acute Care Common Stem (ACCS) - Anaesthetics Trainee | I didn’t apply but had lots of colleagues who did. There seemed to be a lack of coherence between different college tutors who I assume were as blind as us to the application process. Different colleagues were told to do different things. | -0.068 |
| 36-40 | Male | Non-training Fellow, SAS or other Trust-Grade post | Completely let down by the RCoA and HEE. I have completed all parts of the primary (where others who were assigned posts had not, which is a real insult to us who have worked hard to pass the VIVA and OSCE which was previously a pre-requisite) I have a further 4 years of anaesthetic experience which seems to be a disadvantage compared to those who have completed just CT1 and 2. One of the extra years experience is due to having to take extra time to pass the exams, which is an issue that the RCoA are aware of quoting it as the main reason for the attrition rate 0f 48% from CT2 to ST3. Additionally there has been a paucity of posts for the last 2 rounds not just the latest (although it is the latest which has made the newspapers.) As such there are an increasing amount of non-training grade doctors who have the requirements to start an ST3 place but also a decreasing amount of posts. Essentially there has been created an enormous bottle neck from the last 2 years of hundreds of non-training grade doctors who can’t get a post, which will continue to grow. The RCoA sounds increasingly like a government organisation rolling out the same vitriol again and again but with no action, my colleagues have lost faith, they do not represent us. We are doctors who have dedicated our professionals lives to be anaesthetists and have made huge sacrifices in terms of family, housing etc and have been abandoned with no plan in place to support us. Instead the RCoA is championing Anaesthetic PA’s rather than the anaesthetists. I have no confidence in the RCoA, a feeling which is echoed by many of my colleagues up and down the country. Many of my colleagues have given up on their dream of becoming anaesthetists despite passing all the exams and have changed medical careers, I fear the problem will continue and we will loose hundreds of good anaesthetists in favour of the Anaesthetic PA’s. I and my fellow colleagues am utterly, utterly dejected. | -0.067 |
| 31-35 | Female | Anaesthetic Core Trainee | Feel really demoralised by the situation. Stressed and concerned about the future. When you start core training there is an expectation that it you meet the criteria there will be a job for you, I feel really let down by system. | -0.066 |
| 31-35 | Female | Non-training Fellow, SAS or other Trust-Grade post | In my area there are a lot of rota gaps but the ST3 jobs were cut proportionally more so than any other region. I personally scored high enough to traditionally get a job in any area but due the cuts did not get a job in my chosen region, which is typically not a highly competitive area. I strongly feel that the royal college has let this cohort of trainees down. Having worked hard taking the majority of the extra work load caused by covid in our hospital, was this the most appropriate time to cut job and impose an 18month recruitment freeze? I was at several online discussion forums held by the royal college where this concern fell on deaf ears and the difficulties the college are putting a large group of trainees was not even acknowledged. My area has lost numerous trainees, as a direct result of the current recruitment issues, to other specialties and overseas training. In a time when the royal college has issued statements stating they are committed to increasing the number of anaesthetists this whole situation seems ludicrous. There has also been a lot of work produced regarding trainee well fare but the colleges actions are having a large negative impact. | -0.066 |
| 25-30 | Female | Anaesthetic Core Trainee | Too few seats, not very helpful feedback from the intervierwers. I felt like the selection criteria that the interviewers were holding candidates to was unattainable for somebody at core level. My feedback was along the lines of ‘as expected’ and ‘typical for level’ but it seems like being at a core trainee level is not enough to qualify for an ST3 post. | -0.064 |
| 25-30 | Female | Acute Care Common Stem (ACCS) - Anaesthetics Trainee | Extra jobs available after upgrade deadline passed and therefore offered to people who had ranked low but not already accepted a job. All jobs should be rescinded and offers reallocated based on scores. | -0.064 |
| 36-40 | Male | Acute Care Common Stem (ACCS) - Anaesthetics Trainee | The whole process has been incredibly frustrating. I decided to sit my primary early, to then concentrate on audits etc… later in my training. During the first wave, there was no audit activity or regional teaching opportunities at all in my place of work. By the time I started my CT3, there wasn’t a huge amount of time to complete things ready for application. I know why it was decided that you could apply for ST3 with only the MCQ, but I didn’t feel like the same allowances were made in other areas. So, by working hard to pass the full primary, I feel I was actually disadvantaged having done so. I should have spent that time working on audits etc.. instead it seems. The changes to the curriculum were also discussed, then delayed because of the pandemic and then introduced anyway. It’s been incredibly stressful trying to work out where this leaves us all. I cannot see getting a job in the final ST3 round in Feb is going to be any easier and can’t help but think there will be far more applying for ST4 when the opportunity arises. All of us at work are now desperately trying to work out how to build up portfolio points. The process of which I cannot see will make us any better at what we do. | -0.062 |
| 25-30 | Male | Anaesthetic Core Trainee | Clear lack of future planning regarding the Anaesthetic workforce. Feel there has been a real lack of support for the current trainees. | -0.062 |
| 25-30 | Male | Non-training Fellow, SAS or other Trust-Grade post | 1. Pre interview Points scoring system punished you for doing extra ICU which makes no sense 2. The interview was too short and the subsequent scoring seems to be based on three clinical anecdotes that could be made up. 3. The communication from the college and ANRO is a disgrace. The RCOA using Twitter as its primary means of communication is ridiculous. 4. The interview process seems to be far too subjective - my interviewers gave very different t scores | -0.06 |
| 31-35 | Female | Anaesthetic Core Trainee | Was stressful as I only received offer in clearing when 5 extra posts were suddenly added. Many applicants didn’t receive a post and are disheartened with the specialty as a result | -0.059 |
| 25-30 | Male | Non-training Fellow, SAS or other Trust-Grade post | Simply put- there are not enough training jobs for the amount of anaesthetists applying for the specialty There are rota gaps across the board and I’ve been told that some regions can’t increase their training numbers as it would detract from other areas. We need more anaesthetists nationally- therefore we need more training numbers across the board We also need guarantees that previous experience will count for a CCT | -0.057 |
| 25-30 | Male | Anaesthetic Core Trainee | I feel very anxious that I will find it very difficult to ever achieve a speciality training number in my preferred region due to the very high competition | -0.057 |
| 25-30 | Female | Acute Care Common Stem (ACCS) - Anaesthetics Trainee | My main concern is that we have worked so hard in the past year with COVID, out of my 2 years of core training which was supposed to be anaesthetics with 4 months ICU, I’ve now done nearly 12 months of ICU therefore it was a decision not to apply for ST3 which I took because I didn’t feel competent and of the concerning affects this would have, as well as the fact my CV is empty because all my time has been spent on COVID ICU. But now I can’t apply to get a training post for another 2 years which is huge! This is a significant time to be â€œforcedâ€ out of training because it would not be my decision to be out of training for that long, I was only originally planning 6 months. And I dread to think of the competition ratio in 2023, now no-one got a job this year, plus some from last year plus the next 2 years! | -0.054 |
| 25-30 | Male | Anaesthetic Core Trainee | This situation has led to me feeling completely undervalued. This specific recruitment round has been extremely difficult, with a lack of communication around a likely predictable situation, and extremely reactive rather than pro-active reactions from all bodies involved. More worryingly, the wider application process is no longer fit for purpose. The inflation of scores is leading to unobtainable points needed on self-scoring, unless you are either able to take time out of training in other clinical areas or are able to pay thousands of pounds for post graduate courses and qualifications. The huge amount of applicants within a few points of one another shows the incredibly high standard of candidates across the board and an inability of this system to differentiate them. The amount of pressure this system is putting on trainees to not only be diligent, responsible clinicians but also flawless auditors, researchers, educations, leaders, managers etc all while passing relevant exams and working in one of the most high acuity clinical areas is detrimental and is causing a huge amount of extra stress and exhaustion in trainees. | -0.053 |
| 31-35 | Male | Anaesthetic Core Trainee | Many of my excellent colleagues missed out on posts. This really isn’t fair. Core anaesthesia has very few ST3 destinations so creating a massive bottleneck halfway through the anaesthetics program is profoundly flawed and deeply unethical. Talent will leave and never come back. Who can blame them after the way they’ve been treated? | -0.052 |
| 31-35 | Male | Non-training Fellow, SAS or other Trust-Grade post | My concerns are number of ST4 post available in August 2023 with huge number of current ct and accs post given out this recruitment. I think it will be very competitive and I felt the recruitment should go back to the old system where we use regional recruitment rather than national recruitment as it will disrupt our career and resettlement issue with family. | -0.051 |
| 25-30 | Female | Acute Care Common Stem (ACCS) - Anaesthetics Trainee | There are so few ST3 post therefore no movement in fellow/staff grade jobs in the region and therefore high possibility of not being able to get a permanent job on completion of training. In addition, many of the fellow/staff grade jobs are being paid at the CT1 nodal pay point mean I will reduce my pay by £10,000 - £15,000 despite working at a high level of responsibility than I currently do in the training programme. I also feel the college have completely dump us as trainees that have work extremely hard during the pandemic, been redeployed, sat exams and are now faced with no jobs!!! | -0.051 |
| 31-35 | Female | Non-training Fellow, SAS or other Trust-Grade post | 1. Felt like there was a change in goal posts with the full primary FRCA no longer a pre requisite. With retrospect, if I had abandoned my successful attempt at OSCE/VIVA and presented a poster instead, I would likely have a job. 2. Seemingly no back up plan. Many trainees feel left behind. Presumably competition ratios will be the same if not higher for st4 in 2023. 3. Why must we wait until 2023 for next entry point? 4. I will have accrued 6 years out of training by 2023, as per the current person specification this would make me a much less attractive candidate. Seems unfair 5. I imagine, like previous years, there will be many rota gaps in departments across the country. This will be extremely frustrating for trainees denied a number, who will likely pick up the slack of rota gaps 6. When will there be information about Feb intake for st3? Would be useful to know ahead of time so that necessary preparations can be made. | -0.051 |
| 31-35 | Female | Anaesthetic Core Trainee | I am concerned that once again this will have adversely affected female and LTFT trainees. The male bias in the previous round of recruiting (self-scoring without checks) still needs to be addressed. I am not confident in any way that I will be able to achieve what is required to get a post in training and am considering leaving the profession. The uncertainty around training and the lack of initial support from the college has been sickening. Too little too late. The future of the profession will not forget this betrayal. | -0.051 |
| 25-30 | Male | Acute Care Common Stem (ACCS) - Anaesthetics Trainee | Highlights a consistent lack of empathy and forward planning in organisational bodies that is ironic given the increasingly high standards that are imposed on trainees to not only display but have tangible evidence of. | -0.05 |
| 25-30 | Male | Not currently in contracted employment, or currently a locum | The interview was only based on commitment to specialty/communication skills/reflective practice and NOTHING clinical. The whole process was not really explained properly and feel people may have lost out due to this | -0.049 |
| 25-30 | Male | Non-training Fellow, SAS or other Trust-Grade post | Broadly speaking I think a lot of trainees feel let down by the college. Training has been disrupted for a number of us and we have had to adapt and slot in to different rotas to help cope with the pressures of the last 12 months. To be faced with the uncertainty of a curriculum change, reduced opportunities to work abroad, increased competition numbers has been difficult. There is no clear strategy to reduce the backlog of trainees who are appointable but are now stuck in limbo. I am concerned that the longer this continues the greater the risk of good trainees dropping out of training - a huge waste of resources. | -0.049 |
| 25-30 | Male | Anaesthetic Core Trainee | 2 main issues: 1) Current CT2s were poorly treated from the beginning - we applied for 2 year CT jobs, were told just before starting that it was actually a 3 year job, and now back to 2 years due to COVID. The latter was understandably unforseen, however the curriculum change and implementation should have been explicitly communicated to us well before we applied. 2) The ST3 evidence verification process was very poorly conducted, with assessors clearly not aware of/not following explicit guidelines on the ANRO website on what evidence was acceptable. I was marked down by 8 points - most of which were reinstated on appeal. Why should we have to go through such a stressful process, meticulously following the guidelines, only to have to appeal in order for them to be (partially) correctly followed? Despite the appeals process, my evidence was still underscored even though there is an explicit line detailing its acceptability in the ANRO guidance. Very demoralising. | -0.048 |
| 36-40 | Female | Non-training Fellow, SAS or other Trust-Grade post | Although I did get a post this year I had to go above and beyond to score highly on both portfolio and interview, at a time when everything has been made more difficult (FRCA Primary, online portfolios etc) as well as a massive toll taken on myself and my family from working ICU through the Covid surges and lack of decompression options (family, holidays, friends, eating out etc). | -0.044 |
| 25-30 | Male | Non-training Fellow, SAS or other Trust-Grade post | Appeal process was a joke, additional evidence wasn’t considered and deemed ‘too late’ - it was lip service rather than a genuine attempt to gather evidence. That people could apply without the full Primary FRCA means next year there will be people unable to progress as they can’t pass the OSCE/VIVA, they will lose the best anaesthetic trainees who are most driven, as they will not content themselves with 2 to 3 years of middle grade work. | -0.043 |
| 25-30 | Female | Non-training Fellow, SAS or other Trust-Grade post | It seems the bar is getting higher and higher. I’ve been working at an ST3 level for the past 9 months and I’m not deemed good enough to get a formal ST3 post. It’s a slap in the face for me when I’ve worked so hard to get my exams first time and worked all through the pandemic at registrar level. | -0.043 |
| 25-30 | Male | Non-training Fellow, SAS or other Trust-Grade post | I am concerned that those who have completed the full Primary FRCA, particularly within their two core training years +/- passing at first sitting are not able to use this to gain points for their application, when other specialties (eg medicine) are doing this. Surely passing this examination in full demonstrates significant commitment to specialty and it seems unjust that those who have completed all parts are being overlooked for ST3 jobs compared with those who have only completed the written. I appreciate that the pandemic has in some ways affected examinations but it has also affected other means of having self assessment points that are counted for in the self assessment. | -0.043 |
| 25-30 | Male | Acute Care Common Stem (ACCS) - Anaesthetics Trainee | I do not understand why core + ACCS anaesthetic recruitment numbers are significantly greater than for registrar recruitment. Because of this imbalance, many trainees like myself have been trained for 2-3 years, worked hard and passed their exams but are mathematically unable to continue to progress. We are not trained to work in other specialties, we have different portfolios and curricula, our exams are specific to this specialty. If this situation continues it is a waste of training resources for the country and a waste of our time as well trained doctors. I feel betrayed. The deal should be that if you work hard, you can have a job. This is no longer the case, and the specialty (and the country) will lose doctors if there is no correction to this mess. | -0.043 |
| 31-35 | Female | Non-training Fellow, SAS or other Trust-Grade post | I have been very disappointed with the recruitment process. This is a hugely important process which will determine where we are to live for the next 5 years and affects not only us but our families. I understand it being a competitive process but this recruitment process does not seem to reward the best candidates. The goal posts have changed from what we had expected from before covid 19 hit and now there is far more weight on the self assessment score which previously was only used to get an interview. I lost points for not uploading my CV as proof that I did a 6 year course, for my teaching program being for medical students and my national presentation being a poster. These details were not stated on the self assessment matrix and I feel if I had had am in person portfolio station i would not have lost points. I would have had my CV and all my presentations and teaching experience. I feel I was disadvantaged massively by having a remote portfolio - is this really how we should be assessing our anaesthetic registrars? This recruitment process is not fit for purpose and is not a way of picking the best candidates. More emphasis on the interview, as has always previously been the case, is key to getting to know a candidate and assessing who is the best for the job. | -0.04 |
| 25-30 | Male | LAT ST3 | I was not initially offered a job in the first wave of offers, but was successful on the final day of offer recycling (therefore had 2 weeks to contemplate my position). Having done a LAT3 year and met the competencies to achieve stage 1 equivalent certification, I did not require a top-up year, and so with no recruitment to ST4 in 2022, would potentially be out of training for at least 2 years. There did not seem to be any consideration put forward to trainees in a similar position. The fact that higher specialty training numbers have been going down on average over recent years is baffling when simultaneously reading news articles about the shortage of consultant anaesthetists and how vital anaesthetists have been during the pandemic. It almost seems as though some of the mistakes from the MMC era are being repeated again. On the whole this stirs feelings of anger/frustration and when I thought that I had been unsuccessful in this round, I can vouch for feeling hopeless and considering whether I would return to training in the future or not. | -0.04 |
| 31-35 | Female | Non-training Fellow, SAS or other Trust-Grade post | Given the number of applications to post ratio for Aug 2021 session, increasing the number of training posts would have helped us who made it all the way through interview , however not offered a training number by a very close margin. | -0.04 |
| 25-30 | Female | Non-training Fellow, SAS or other Trust-Grade post | there were clearly a lot more applicants than jobs. There was a lack of clear communication from the RCOA regarding the plans for those who did not get a training post and how their training would be impacted. And the response from the RCOA to the overwhelming amount of applicants who did not get a training post felt completely out of touch with the gravity of the situation. | -0.037 |
| 25-30 | Female | Non-training Fellow, SAS or other Trust-Grade post | The whole process lacked transparency. As a non-training fellow, I was left out of Deanery communications and HEE communications, and only heard important pieces of information via colleagues and social media. The portfolio assessment process lacked transparency and a lot was left open to interpretation- for example, I was docked a point for portfolio organisation because I did not upload commentary in each domain and the assessor had to cross-reference to my CV- yet the Oriel portfolio did not have a space to write comments! My other main grievance is with the College pressing on with a curriculum change that made the ST3 application process so competitive this year, with seemingly no proper plan in place for the 700 trainees left behind. Either more training places needed to be created, given the anaesthetic workforce vacancies, or there needed to be proper plans made for the ‘top up jobs’- at the moment a list of curriculum requirements, not even connected to the LLP, is all that exists, for these posts that departments are supposed to organise despite not being funded to do so. It is most disappointing that these 700 trainees, many of whom were instrumental to their departments’ COVID responses, have now been cast aside and left to sort themselves out. I have been a fellow for a year, and have to say, expecting anaesthetic departments to treat fellows out of training equally to trainees and facilitate the top-up competencies, despite no funding, is idealistic nonsense- I have struggled to receive any of the privileges trainees do during my fellowship, let alone basics such as parity of pay (for reference, I am paid £560 less per PA of work per year than I was as an ACCS trainee). | -0.035 |
| 36-40 | Male | Non-training Fellow, SAS or other Trust-Grade post | Starting a ST3 ICM post may complicate the ability to complete a standalone ct3 year. Which would prevent me from dual training. | -0.034 |
| 25-30 | Female | Non-training Fellow, SAS or other Trust-Grade post | I feel upset with the whole process. Although I did get a job, I think that so many have been left behind and it is extremely unfair. The cohort of doctors applying for ST3 have had so many changes affect them, and I feel so disappointed in the College that is supposed to look after us. What has happened this year shouldn’t have come as a surprise - we all knew the curriculum change would create a bottleneck so I am saddened that this has been allowed to happen, with no provision for those who missed out. I understand the huge challenges that the RCOA has been under the last year, but surely safeguarding and looking after the workforce should be prioritised, especially when we have been so encouraged to look after ourselves. All the well-being messages feel very empty when really what people want is job security. when you are made to reapply through a highly competitive process (plus the primary, all the modules etc) you’ve already been through two years earlier, and at the end don’t get a job no amount of well-being initiatives from the RCOA is going to help. I didn’t get an ST3 job last year and felt ignored and forgotten about by the college. There is was no information or support for people in my position (this is something the AAGBI could make and would have been very welcome!) | -0.032 |
| 31-35 | Male | Non-training Fellow, SAS or other Trust-Grade post | Having spent 2 years doing what the application requires to a high level, it remains this difficult to progress. Having passed the full primary, commitment to the specialty is still a part of the specification, the exam is no easy feat. The largest specialty in the hospital at a time when needed the most is now this difficult to enter? I am not sure the true long term plan for anaesthesia is here. | -0.032 |
| 31-35 | Male | Non-training Fellow, SAS or other Trust-Grade post | I feel that the selection process favours people who “play the game” or are able to “talk a good game” but not people like me who are able to do the job really well but are more reserved in nature and find it more difficult to sell themselves in an interview or lack the ruthlessness to grab the opportunities to score points on the self-assessment. This does not make us bad doctors or unworthy of progressing. A large part of the blow is that there is no planned recruitment round for August 2022 and I can’t really understand why. The vast majority of those who did not get a job this year probably get their CT3-equivalent competencies and would therefore be in a position to a start ST4, and with the numbers who were not successful, an August 2022 recruitment round would still be reasonably competitive. Delaying to 2023 when there will be a fresh batch completing their core training to add to the numbers will only exacerbate the problem. Holding an August 2022 ST4 recruitment round would be an excellent chance to get more people back into training and moving their careers forward. Those starting ST3 this August will be transitioning over to the new curriculum from August 2022 anyway, so I really don’t see the need to not recruit next year, especially when the college seems to recognise the workforce shortages at all levels and increasing future demands in anaesthesia. I recognise that the college is not responsible for setting training numbers, but I can well imagine that they were involved with the decision not to have an August 2022 recruitment round, and simply don’t understand that decision. A change of heart in favour of holding an August 2022 recruitment round would surely recognise the hard work and unprecedented sacrifice that this group of doctors in particular have made during the pandemic of the last 12-18 months and would certainly be a major boost for their morale, well-being and mental health. | -0.031 |
| 31-35 | Male | Anaesthetic Core Trainee | Lack of ability to appeal discrepancies in self assessment, when 2-3 marks was the difference of 60-80 rankings . The annual number of Core / ACCS anaesthesia exit posts should be at least matched to ST3 posts available OR training should be run through. Otherwise this is unfair on trainees who can get stuck. There are also international graduates entering at ST3 which further increases competition at this level | -0.031 |
| 25-30 | Male | Non-training Fellow, SAS or other Trust-Grade post | In my opinion, the interview did not give an opportunity to demonstrate our strengths for the role. There were 13+ questions over a 30 minute interview, none of which were on clinical management and there was no way for the interview panel to verify the information. After being disappointed with my interview score, I requested further feedback. One examiner stated that I had unstructured answers, whereas the other interviewer stated I had well structured answers. However, the second interviewer marked me down on the basis that it was not possible to see my entire face on the video screen. However, this had been checked by the administrator before starting the interview, was not mentioned at any time during the interview, and appeared completely normally on my screen. Given the impact to my family as a result of being marked down, I was extremely disappointed by this feedback. | -0.03 |
| 31-35 | Male | Non-training Fellow, SAS or other Trust-Grade post | It’s a massive kick in the teeth. I had my exam cancelled, I’ve been delayed in moving on twice and I’m now being asked to jump through extra hoops again. To bolster my portfolio points now I need to do a 2 year MD or lead a research program or do time in another speciality. What more do you want from me? I’m very close to losing trust and faith in the training system. | -0.03 |
| 31-35 | Male | Anaesthetic Core Trainee | Insufficient number of training places Having to move to continue with training | -0.029 |
| 25-30 | Male | Acute Care Common Stem (ACCS) - Anaesthetics Trainee | Far too few training posts in Severn. Fellow posts have fewer protections than training posts meaning I will have a significant pay cut | -0.029 |
| 31-35 | Male | Non-training Fellow, SAS or other Trust-Grade post | Lack of available posts, unclear instruction from the college re ‘top up posts,’ ad-hoc and piecemeal placements advertised from regions/trusts. Potentially having to move home again to pursue career, for 3rd time in as many years with no certainty or security going beyond a year at a time. Stress on personal relationships. Feels like being given a choice between personal life and goals or continued profession/specialty. | -0.029 |
| 31-35 | Male | Non-training Fellow, SAS or other Trust-Grade post | Predictable increase in number of applicants for the same or reduced numbers of places through combination of curriculum changes, COVID, criteria changes (including not requiring full primary FRCA and potentially removal of RLMT) as well as a persistent discrepancy between the number of core and ST training numbers, despite the ongoing shortage of anaesthetic and ICU consultants and a clear need to expand the workforce. The potential impact of the curriculum change and the change to recruitment at ST4 on trainees who have already completed core training has been apparent for years and overall there has been little in the way of concrete assurance that there would be a straightforward way to demonstrate equivalence and become eligible for ST4 recruitment if unsuccessful in this round - although some CT3 style posts are now being advertised this was after recruitment was all but completed so at the time before receiving an offer I felt no guarantee that I would be able to access such posts; there should be a formal mechanism to ensure all trainees who have completed core training and wish to continue with anaesthetics training are offered a CT3 post, and not a reliance on individual trusts filling gaps in the system. | -0.026 |
| 31-35 | Female | Non-training Fellow, SAS or other Trust-Grade post | I’ve been putting starting a family on hold until I get a registrar training job. I’m not prepared to wait another 2 years now. Bad timing that if I had started my family a few years ago as I wanted, I probably would have missed the bottleneck. It does make me wonder why bother with the effort of trying to plan life around my career now that I’m in this position - you make so much personal sacrifice and get nothing in return. I really regret not spending more time with my friends and family over the past few years. If I don’t get a CT3 job I’m planning to leave medicine, I’ve really had enough. In my hospital 6 of us applied for ST3 jobs, 3 of us (Female) can only stay local due to having caring responsibilities (Children or unwell parent), none of us got jobs. The 2 men with no caring commitments were able to accept training jobs across the country/Wales. I’m sure the current bottleneck particularly affects those with caring responsibilities. | -0.026 |
| 25-30 | Male | Non-training Fellow, SAS or other Trust-Grade post | Lack of transparency and consistency with conduct of interviews. High weighting of interviews, which provide poor differentiation between candidates based off subjective criteria. Lack of training posts available in a specialty with proven workforce deficits at Junior and Consultant levels. Poor planning with respect to transition period to new curriculum - lack of recruitment in August 2022 will inevitably lead to massive competition for August 2023. | -0.024 |
| 25-30 | Male | Anaesthetic Core Trainee | Passed exams early yet not taken into account for points when applying. I thought one of the main reasons for the curriculum change was to allow more time for trainees to pass exams yet disadvantaged having passed all the exams, with time spent revising rather than enhancing application strength. Also the change of working contract from CT2 to CT3 then back to CT2 was not helpful. Still not released the cc three top up portfolio section on lifelong learning platform either. | -0.024 |
| 31-35 | Male | Not currently in contracted employment, or currently a locum | The hiatus in further recruitment for ST jobs until August 2023 was not made clear from the RCOA website and required specific searches after being told about it by a fellow trainee. Consultants in positions of trainee responsibility that I spoke to did not seem to be aware of it either. For a curriculum change that is occurring due to a failure to progress core trainees to higher training, this seems to have backfired somewhat. | -0.023 |
| 31-35 | Male | Non-training Fellow, SAS or other Trust-Grade post | I was eligible to be interviewed both times ( feb and aug 2021) and have a well rounded portfolio I think . But despite that wasn’t successful in getting a ST3 post, and the feedback from both times were â€œneeds more interview practice â€œ. I feel let down and unsure about the future in Anesthetics . If there is a face to face interview with clinical element to it or if the interviewers have our portfolio on the day of interview I feel I might stand a chance else the future in Anesthetics looks bleak . | -0.022 |
| 25-30 | Male | Anaesthetic Core Trainee | This system currently has no recognition of clinical ability. There are no points for hard work, honesty and charity. The self assessment only rewards academic work. Academic work doesn’t necessarily mean a good anaesthetist. International presentations don’t make you a good anaesthetist etc. I got very lucky. The issue is across the board | -0.021 |
| 36-40 | Male | Non-training Fellow, SAS or other Trust-Grade post | I just want to add that the recruitment was a whole non sense process particularly the interview. How come in 15-minute remote meeting with typical cleche questions you would evaluate someone out of 800+ on scale of 5s !!! It was a process for interviewers prospective where whoever gives 4/5 for what he/she sees is the best v.s. who give 5/5 for same performance!! which ended up in a situation where 2 marks would make tens of different in ranks for no obvious reason or points where a development plan can be done!!! It was not a fair competitive it was a jackpot where lucky was the key of the game. Your process literally ruined people’s lives. | -0.02 |
| 36-40 | Female | Non-training Fellow, SAS or other Trust-Grade post | After working incredibly hard throughout a global pandemic, I feel very let down that there are not enough training posts allowing progression. | -0.019 |
| 31-35 | Female | Non-training Fellow, SAS or other Trust-Grade post | This recruitment process has been extremely difficult and exhausting. Firstly, the way that we are scored does not reflect the atttributes of what makes a good anaesthetist. Candidates that are good at tick boxing exercises, participating in research and doing MRCP are getting the jobs/jobs in highly competitive regions, rather than it being about the non-technical skills that are important. Most people in the interview perform ‘avergagely,’ so the portfolio then massively has a swing on who gets jobs or not. After the interviews, I realised that some people were interviewed by Consultants that knew them, which brings out a massive element of bias into the process. Finally, in Scotland, there were five new jobs recycled from ICU numbers at last minute, but for some reason they weren’t recycled till after the upgrade deadline. This meant that people with significantly lower rankings got their first choice job (if they only put one region) in highly competitive regions. This completely undermines the whole interview process and it is extremely disheartening to hear of this, especially when these jobs were requested to be recycled on the 5th May (and weren’t till 17th May). This is trainee’s lives we are talking about. Their wellbeing. The places they want to live for a significant period of their life. The recruitment process does not sympathise with this at all. | -0.018 |
| 25-30 | Male | Acute Care Common Stem (ACCS) - Anaesthetics Trainee | I believe that the ANRO have created a bottle neck in recruitment by having CT1 posts far in excess of ST3 posts. I acknowledge that there will always be a drop out rate and/or people who take longer to complete core training, however, the discrepancy in posts between CT1 and ST3 appears to be notably in excess of this . I therefore feel strongly that special consideration needs to be made by the powers that be to avoid this unfair situation where large numbers of trainees that have dedicated 2+ years to anaesthetic training, completed their core training, and undergone all the required stressful examinations are being “squeezed out” of the opportunity of completing higher training. It is these people who should be prioritised for the future of the speciality and it may be necessary to restrict CT1 entry until the bottle neck can be cleared. | -0.018 |
| 25-30 | Male | Non-training Fellow, SAS or other Trust-Grade post | Removal of resident labour market test disadvantages trainees who have completed core level training and face a now even harder bottleneck at st3/st4 What proportion of those successfully getting st3 numbers this round without the exam are actually on say attempt 5 or 6, and will continue to fail and subsequently relinquish the number? In the deanery I applied to, many trainees are on mat leave/OOP/delayed CCT, leaving numbers occupied and without freeing up spaces for new trainees - only 3 numbers available in that region. Is there a way to work around this to allow trainees in? Entire application process is biased towards trainees with ‘tick box’ achievements. Does having a PhD (wildly unachievable for most post core trainees) really make you a better anaesthetist? There needs to be more assessment of clinical ability, use of references, letters of recommendation alongside how many publications you’ve got. The process as it stands currently massively disadvantages those without the means to do all the life support courses, conferences etc. | -0.017 |
| 25-30 | Male | Acute Care Common Stem (ACCS) - Anaesthetics Trainee | Trainees forecast the problem but no steps were taken to address this; no coherent national plan for CT3 posts; pig-headed approach to pushing though the new curriculum against an overly tight timescale (contributing to previous points); defence of the new curriculum being it will make trainees lives easier, reduce stress and improve progression with no hint of irony that it is causing the opposite for those of us stuck in the gap. Personally, I won’t buy a house until I have a reg job. My current landlord wants to move back in to my rented property so I am currently unemployed from August, and will be homeless from September, and I can’t fix the home problem til I fix the job problem. This is a really small point, but we keep being referred to as ‘unsuccessful’ or ‘not in training’ with there being no explanation that that is not our fault. I have been successful - Primary FRCA MCQ passed, sitting OSCE/SOE for first time this week, all of core curriculum covered, audit completed etc. I was appointable. They didn’t have a job for me. There’s a difference. | -0.017 |
| 25-30 | Male | Non-training Fellow, SAS or other Trust-Grade post | It was a very stressful process. Having not succeeded in August 2020 application, I had to secure a non training grade post last year after completing CT2. This was a difficult process with very little guidance at the time. I was again unsuccessful for February 2021. The stress of working on covid rotas, as well as having no study budget etc on my current role while trying to boost my cv to make me more “employable” has pushed me to the brink of burnout. I was one of the lucky few to get a job this time round, but I wonder how much toll the process has taken on my mental health with the amount of stress I and my colleagues were under. I feel so sad and angry on behalf of my colleagues who were unsuccessful in this application round. Some of the applicants are extremely talented anaesthetic CT2s who undoubtedly would have got jobs a few years ago. Furthermore, I very much doubt many of the CV boosting activities I did have made me a better anaesthetist - in fact I am due to start ST3 in August having not even given an anaesthetic for the last 12 months because the application process gave more weight to me working in Emergency Medicine as an SHO rather than getting more experience in anaesthetics. A last note re fairness of the application process. Many CT2s have lost out on training posts to experienced anaesthetists trained abroad who have applied for ST3 jobs as a 5 year fast track to gaining a CCT in anaesthetics. These anaesthetists are very talented and often consultants back home with many years of experience in anaesthesia. They are often working in jobs (e.g. senior clin fellow/staff grade type positions) that UK trainees post core-training are not even able to apply for due to lack of experience. Allowing them to subsequently apply for a 5 year TRAINING programme seems very flawed and an insult to them and the UK core trainees who are unable to get these jobs. A training job should serve the purpose its name suggests - to TRAIN anaesthetists. I fully agree that experienced anaesthetists from abroad have lots of experience to offer our NHS and they should have the opportunity to become UK consultants, but if they have already completed training elsewhere, surely this should be via the CESR pathway that is not appropriate for post-CT2 trainees. It seems absurd that they are having to opt for ST3 training because of how difficult CESR training can be. The RCOA website says that the MTI scheme is not a route into ST3 training, but I have seen anaesthetists completing the MTI scheme do just that successfully gain ST3 jobs in this round. So to protect our national training scheme whilst also increasing the number of trained anaesthetists to bolster the workforce, should we not be aiming to simplify the CESR process for these very talented and brilliant anaesthetists who are providing their expertise to help our (failing) NHS? | -0.016 |
| 25-30 | Male | Intensive Care Medicine Trainee | Although I am fortunate enough to have an ICM ST post, not having an anaesthetic post has prolonged my Stage 1 training. I will be unable to prolong it further to make it to the August 2023 intake for ST4. I am now faced with having to single-speciality in ICM, or progress to stage 2, having to repeat speciality placements when I get an anaesthetic job. This will further prolong my time in training. Another of my concerns has been the situation regarding the Primary FRCA and applications. I realise that it is unfair to candidates who had disruptions due to the pandemic, but it feels odd to be competing with people who have not passed the primary for a post where it is a requirement. Given how competitive the self assessment scores have been, it definitely feels that my time prepping for the exam could have been better spent in research/publication/presentation/ teaching. Guidance at the from my supervisors and college tutors was to focus on the exam, even to the point that study budget and study leave was restricted for non-primary topics. I feel like now this advice has not benefited my career progression. | -0.015 |
| 25-30 | Female | Acute Care Common Stem (ACCS) - Anaesthetics Trainee | I feel totally let down by the college. Not getting a job isn’t the biggest issue, it’s the lack of foresight about the situation we are now in. I am going through multiple applications and interviews with no certainty of where or even if I will be working come August. The college seems to be surprised by this situation, whereas it was entirely predictable given the change of curriculum compounded by Covid resulting in a large number of applicants. It goes without saying that this is an incredibly stressful situation on top of an already horrendous year involving applications, interviews, exams and a global pandemic. I have jumped through every hoop put in front of me and done everything asked of me and I feel like I have been treated like a number on a spreadsheet rather than a person. | -0.014 |
| 25-30 | Male | Acute Care Common Stem (ACCS) - Anaesthetics Trainee | The pandemic is an unprecedented event that required incredible sacrifice from all anaesthetic trainees. Despite all of this the RCOA has shown little flexibility in their plans. There were more trainees applying for st3 jobs with less jobs available than most previous years partly due to delays to career progression caused by covid. The implementation of the new curriculum should have been delayed further to allow the bottleneck to be reduced a bit more. | -0.013 |
| 25-30 | Female | Non-training Fellow, SAS or other Trust-Grade post | They keep recruiting more CT1s but fewer ST3 posts each successive round. Massive bottleneck, in my region, I know lots of trainees who are 1-2 yrs out of core training still applying for ST3 posts and without jobs. Rota gaps everywhere and propping up the itu rotas during covid working as ST3 equivalents, but won’t give us more training numbers. Especially frustrating while trying to pass the Primary FRCA. Also for last 1 year, college hadn’t released competencies for CT3 equivalent, leaving those who were already post CT2 flying blind. RCOA have messed up big time and it’s not enough just to blame HEE, they’ve known for years they were moving to ST4 recruitment and have failed to keep trainees updated on any changes. | -0.011 |
| 31-35 | Male | Non-training Fellow, SAS or other Trust-Grade post | Lack of employment security, difference between SAS and AS jobs life versus a non training junior doctor. We work long hours, antisocial, for ST3 pay without the promised land of consultancy. The pace of full time training is OK for short periods but for me is imcompatible with starting new aspects of my life: having children or buying a house. This has cemented my plan to undergo the NZ Anaeshesia training programme which is just five years long (with upto two years recognised prior learning) | -0.007 |
| 25-30 | Male | Acute Care Common Stem (ACCS) - Anaesthetics Trainee | Why did the primary FRCA not give candidates any points? Whilst exams were cancelled in spring 2020, there were many opportunities since then and this is underlined by the numbers of trainees passing the primary. This is on the background of awarding points for MRCS, MRCP and EM exams. This doesn’t make sense | -0.007 |
| 31-35 | Male | Acute Care Common Stem (ACCS) - Anaesthetics Trainee | Concern that ST4 applications in 2023 will be even more competitive, potentially having to decide whether to relocate with young family or leave the specialty | -0.006 |
| 36-40 | Female | Acute Care Common Stem (ACCS) - Anaesthetics Trainee | I feel there has been no provision from HEE for those that are left in the Â«Â gapÂ Â» who have no ST3 number to move into. The top up postes available are not HEE funded and it makes it very difficult to know what level of training we will actually achieve, and what recourse we have if we don’t actually get Â«Â trainedÂ Â» as advertised. Will we have anyone to appeal to if our training needs are not being met? I worry after investing years of my time into a competitive specialty, and sacrificing energy and money into expensive and stressful exams i will now be disadvantaged due to the ST gap, my training will be prolonged or perhaps even lost to me. | -0.006 |
| 31-35 | Male | Non-training Fellow, SAS or other Trust-Grade post | I initially fell off the ‘bandwagon’ after I became ill with a critical illness, delaying me taking my OSCE/VIVA. Not long after this the pandemic happened. Had I wanted to get through training ASAP there is no way I could have. Very skilled colleagues have been denied jobs on the most pinickity analysis of their portfolios. I doubt I will ever be able to assemble a portfolio water-tight enough to get through such extreme pedantry. Now that I work (and enjoy working) as an ITU Fellow, I will never get paid to be trained in enough Obstetrics for ST4, so if I ever decide to try to progress I will have to switch to core medical training and then try that route into just ITU. | -0.003 |
| 25-30 | Female | Anaesthetic Core Trainee | - shortage of jobs given the results of 2020 anaesthesia census. How will we increase the number of anaesthetic consultants if we don’t have posts to allow anaesthetists to compete their training? - Interview was very generic and scoring very subjective. - Unusual to be asked very general questions by a panel who don’t know your CV. It was difficult balance to answering the question directly and informing the interviewer of all your experience to date - Marginal differences in an application overall score (from subjective scoring in matrices which don’t necessarily reflect candidate ability to do the job) had dramatic effects on ranking and whether people got jobs - This has had a significant effect on people’s lives, particularly as not getting a job means 2 years out of training | -0.001 |
| 31-35 | Male | Acute Care Common Stem (ACCS) - Anaesthetics Trainee | I feel used and let down, and it’s knocked me down to the extent where I’m considering leaving medicine | 0 |
| 31-35 | Female | Anaesthetic Core Trainee | I feel let down by the college as it has constantly changed the goal posts for my core training. When I started it was 3 years and then it went back to 2 and now I don’t even have a training post. | 0 |
| 25-30 | Male | Acute Care Common Stem (ACCS) - Anaesthetics Trainee | None | 0 |
| 25-30 | Female | Anaesthetic Core Trainee | N/A | 0 |
| 25-30 | Male | Acute Care Common Stem (ACCS) - Anaesthetics Trainee | There are too few training posts in Severn | 0 |
| 25-30 | Male | Anaesthetic Core Trainee | My interviewers looked bored heading out my teamwork / leadership / audit experience relating to covid and extra itu. I feel undervalued. The North West Mersey Deanery hasn’t centrally created CT3 posts and there is no forum for Q&A s for how to approach the next steps. | 0 |
| 31-35 | Male | Non-training Fellow, SAS or other Trust-Grade post | Curriculum changes have lead to a clear bottle neck in the recruitment process and I feel the college should have provided reassurance regarding the CT3 LAT posts to bridge the gap in training for current CT2s who have not been successful. Having spoken to some of my colleagues I think a lot of very capable and enthusiastic anaesthetists will be lost from the specialty as a direct result of this and given the shortages of anaesthetists across Scotland this seems pretty laugable. | 0.001 |
| 36-40 | Male | Non-training Fellow, SAS or other Trust-Grade post | I feel there is always a bad bottleneck going into ST3 and COVID and curriculum change has hugely exacerbated the problem. I also feel there were fewer jobs available than normal, despite HEE saying otherwise. It seems crazy to have a points based discriminatory system whereby “not enough points = no interview. Just because you have a PhD or multiple publications, for example, does that mean you are a better anaesthetist than someone that doesn’t? I consistently get excellent feedback from colleagues and patients and yet this counts for nothing when surely it should count for everything in such a patient-facing and team working job. It makes me feel very demoralised about the whole thing. Another option is CESR, but I have heard (and read in the latest RCOA Bulletin) that it is a clunky and tiresome process. The whole process seems crazy considering how many unfilled consultant Anaesthetist posts there are. The maths is simple: more needed out the top means more needed in the bottom… | 0.001 |
| 31-35 | Male | Anaesthetic Core Trainee | There is now no ST4 applications until 2023. This to me is compounding an already terrible situation. Having just completed a CT2+ due to covid interruptions in my training, I am now looking at two years out before I can apply again. That is three years after successfully completing my core training and passing my primary before I can even apply for higher training again. My major concern (and that of many of my colleagues) is that by August 2023 there will be even more candidates (as there is no 2022 recruitment rounds) and we face the same if not worse competition ratios. This is unacceptable to those who have completed core training and the primary FRCA. I have been unable to buy as a house as a result. It has implications on my family life and it has contributed to a low mood which is very unusual for me. I feel completely let down by the profession and the college. | 0.002 |
| 31-35 | Male | Non-training Fellow, SAS or other Trust-Grade post | Not enough jobs. Not fair process, self assessment points too big a proportion of overall score. Experience outside of Anaesthesia in adult ICM should be valid. | 0.002 |
| 25-30 | Female | Non-training Fellow, SAS or other Trust-Grade post | Very few numbers available when this was going to be a predictably high applicant year. So many reasons for more people applying than usual and just the same number of jobs. Interviews were a joke with 20 mins of essentially osce style questions with no depth, just loads of superficial answers. Self assessment wasn’t reproducible with the same piece of evidence getting different scores depending on who was verifying. The difference of one point is many rankings therefore really doesn’t distinguish between people. None of the ways we are marked actually has any bearing on whether you are a good anaesthetist or whether you are a valued member of a team. Every consultant is saying that not getting a job isnt a reflection on the applicant and doesn’t mean you don’t deserve it therefore the process isn’t fit for purpose. No way of taking into account that my husband was also applying and we were given jobs at complete opposite ends of the country. | 0.003 |
| 25-30 | Male | Anaesthetic Core Trainee | I was misled into believing ST4 applications would be available from Aug 2022 (ie, I could extend my SHO training into a fellowship role from Aug 2021 to July 2022, but if I met the training requirements for CT3 could be signed off by a regional advisor and apply for ST4). This information was from a college tutor from one training deanery (where I applied for my fellowship) and from my regional training programme director (an adjacent deanery). I was then informed I could apply for a CT3 equivalency year, ie attend my planned fellowship, return to a CT3 training post and then apply for ST4. I was then informed you cannot apply for stand alone CT3 position and I would effectively have to collect my own evidence of completion of competencies while sourcing my own employment until August 2023 when I can apply for ST4. If I am then unsuccessful I would be heading into my third year of ‘fellow’ work or staff grade work and have been working in anaesthetics without a number longer than in a training post. I have no guarantees these roles will allow study leave, study budgets or allowing study leave for exam prep or courses (for example for the final FRCA). Despite assurances to the contrary I am sure my application would be looked on less favourably than a trainee having completed their three years routinely, and this would only escalate with any subsequent failed applications (I am tied to the area via family commitments and work in a competitive deanery). I now have one window to apply next February (2022) but would mean my planned fellowship is cut short (I had planned this as I went straight into training from F2) and I would have one shot at this application (where if I had been informed you cannot apply for CT3 equivalent positions an that there will be no ST4 application in August 2022, I would have also applied in this most recent batch). It is disappointing having been redeployed to ICU for extended periods due to Covid, working to pass primary FRCA during a pandemic (having exams cancelled) and now struggling to work towards ARCP competencies we could not have been better informed. I have made significant life choices based on erroneous / incomplete information from the people who are supposed to know and help guide us (I’m not blaming them, they are simply relaying what they’ve been told). I’m only a single voice but overall I just feel let down during what has already been a demanding and demoralising year. | 0.005 |
| 25-30 | Male | Anaesthetic Core Trainee | I feel the entire process has been poorly communicated and the implementation of this transition during the pandemic when training has already been affected will massively effect trainee well-being and drive people out of the specialty. | 0.008 |
| 25-30 | Female | Non-training Fellow, SAS or other Trust-Grade post | The self assessment scoring system is contradictory. You are meant to show commitment to anaesthesia but a lot of the avenues were you score top marks are unrelated to anaesthesia : additional degrees , pHds, time spent in other specialties. Clinical scenario is a good way to gauge a candidates ability to work under pressure, prioritise and show their clinical competencies it should be brought back into the interviews The GMC and RCOA need to streamline the cesr route and remove the stigma attached to the cesr route. | 0.009 |
| 31-35 | Female | Non-training Fellow, SAS or other Trust-Grade post | I know many colleagues who are excellent anaesthetists who were unsuccessful in gaining an ST3 post. Everyone including our consultants are disappointed and angry at the lack of jobs available. | 0.009 |
| 25-30 | Male | Non-training Fellow, SAS or other Trust-Grade post | The whole process felt dehumanised. Especially my interactions with ANRO who were less than helpful. Despite having evidence accepted in the Feb 2021 round it was rejected in the latest round. For some reason I was denied 5 marks in the self assessment score that I had been given previously but awarded 1 extra (incorrectly).This meant my overall change was 4 and I couldn’t appeal. The comments on my self assessment completely contradicted guidance provided. I felt completely let down by the system especially after the year of service I had given mainly re-deployed in ICU. | 0.009 |
| 25-30 | Male | Anaesthetic Core Trainee | I feel the portfolio scoring as a whole was disadvantaging trainees that have opted for Anaesthesia as their primary speciality, and have not transitioned into it from other specialties. Moreover, trainees on an ACCS pathway receive an extra 2 points for their year in medicine, which I feel disadvantages those that are on a pure core anaesthetic training pathway. Those 2 points can be a large discriminatory factor between getting a job and not getting a job | 0.01 |
| 31-35 | Female | Non-training Fellow, SAS or other Trust-Grade post | Frustrating application that does not rely on clinical practice. Need more information about CESR. | 0.011 |
| 25-30 | Male | Non-training Fellow, SAS or other Trust-Grade post | The creation of a sudden bottleneck in training has been extremely frustrating, not to mention wasteful of the resources that have been put into my, and others training so far. The application process, especially portfolio scoring, was opaque and arbitrary. The application process is unfairly weighted towards ‘extracurricular activities’ and does not reflect trainees ability at doing the actual job of anaesthesia, merely how well they can tick application boxes | 0.011 |
| 31-35 | Male | Non-training Fellow, SAS or other Trust-Grade post | I think it’s been a total shambles and a total kick in the teeth after we worked so hard stepping out of training supporting the Covid response. The lack of a solid plan for what happens if we don’t get the last st3 jobs is really concerning, and there is also the fact that this is going to have a knock on effect with competition ratios for a long while. I think it’s bad enough in core anaesthetic training that you have to competitively re-apply to continue your training, at a late stage in life when we have established homes, families etc. We’ve already proved that anaesthetics is the career we want to pursue by taking exams, completing core requirements. Why do we have to prove it again and risk having to move to another part of the country? I feel fortunate in a way that my plans always involved time out of training, I only hope there is a consistent plan to get CT3 sign off completed so I can be eligible for st4 in August 2023 | 0.014 |
| 31-35 | Female | Anaesthetic Core Trainee | I think the fact that the timeline was brought forward was really problematic - we had to apply in December, and this meant that many trainees had not had time to complete projects/do presentations/ extracurricular activities which may have increased the portfolio score. We spent the majority of CT1 focussing on exams and of course the pandemic took over everything, and I know a lot of people (myself included) had been planning to use CT2 to buff our CVs and get extra portfolio points, but we found ourselves in a position where we didn’t have time to do this. | 0.014 |
| 31-35 | Male | Anaesthetic Core Trainee | If there are plans to ‘maximise intake’ for the next round of st3 applications, why was this not done for this round when the bottleneck was anticipated since conception of the new curriculum? Is there hope that trainees drop out so that the competition for places isn’t as fierce (as otherwise there may always be 700 or so not gaining employment with each intake) The intake process occurred at the same time as revision for the frca primary viva reached its peak. This was after the dates were brought forward. Why was this? | 0.015 |
| 36-40 | Female | Non-training Fellow, SAS or other Trust-Grade post | I am someone who gained a good portfolio score (38) however when it came to the interview I was marked down on my global rating score (12/20) even when I scored 8/10 on 4 out of 6 of the sections, I was not given feedback as to why this was. I also was not able to interview in my first choice deanary as the slots were filled up when I tried to book. I ended up being offered a place which I accepted with upgrades in the North East. I was then told that places were available in my first choice deanery (East Midlands ) and contacted ANRO however the only response I got was that the Upgrade deadline has passed and I would not be offered those places. I had no choice but to accept this North East offer as I’ve been out of training for 2 years. However this has made life extremely difficult for me as I regularly need to go back to London to help care for my father who has cancer. I now have a 4hr journey instead of an hour and half to go see my family. This application process has been completely unfair and disadvantaged people like myself who are not in a position to apply for special circumstances as I did not want a job in London where my family is, but wanted to remain in the deanery I completed my core training as my partner lives in the Midlands. | 0.017 |
| 25-30 | Male | Anaesthetic Core Trainee | Overall very poor organisation from HEE and RCOA in implementing curriculum changes before the provision of an adequate number of CT3 jobs. Seems like the onus fell on the Local trusts/schools to create these jobs at last minute. The bottle neck will continue to increase this year with significantly more CT1 than ST3 posts, even accounting for attrition over core training/ACCS. Equally frustrating that the full primary exam scored no points when, over the past 2 years, the exam and the pandemic will have been the main focuses for a lot of trainees. | 0.017 |
| 25-30 | Female | Intensive Care Medicine Trainee | I am an intensive care trainee that has been working full time throughout the pandemic. It has been made exceptionally hard to dual train. I have limited time to develop my cv with certain aspects that are specifically suggest on the self-assessment as I am working towards experience and exams in intensive care. I was heavily marked down in the self assessment process in areas were I had gained letters of evidence by experienced consultants. I feel particularly disappointed and let down by this process. | 0.018 |
| 25-30 | Female | Non-training Fellow, SAS or other Trust-Grade post | Not enough posts for the number of applicants. Reduced number of posts in Yorkshire compared to previous years, despite there being gaps in the rotas across a number of different hospitals. | 0.018 |
| 36-40 | Male | Non-training Fellow, SAS or other Trust-Grade post | Many very good capable trainees have been left high and dry searching for employment with trusts as a result of poor planning from HEE and insufficient training posts. It is wholly concerning that an entire generation of very competent and capable anaesthetic trainees will be left looking at alternative employment and likely leave the profession to other areas and specialties due to this lack of foresight at a high level. Especially when middle grade anaesthetic Rotas are understaffed and haemorrhaging at the seems. A profession that has propped up the nation in its time of need has sadly been hung out to dry by HEE and the government. | 0.018 |
| 25-30 | Female | Acute Care Common Stem (ACCS) - Anaesthetics Trainee | Not enough information provided and certainly not done in a timely manner at all. Just adds to the overall anxiety which is not needed at this time. Expected better from RCoA | 0.019 |
| 25-30 | Male | Non-training Fellow, SAS or other Trust-Grade post | Only 2 interviewers rather than the previous 6 (2 on each station) meaning reduced number of opinion on a candidates employability Very drawn out process- initial applications around October/ November- only found out result in May. Ineffective scoring matrix which doesn’t focus on high quality anaesthetic and medical care delivery- should be more focus on MSF and colleague feedback | 0.019 |
| 25-30 | Male | Anaesthetic Core Trainee | This year’s recruitment allowed applicants who had not passed the FRCA to apply for the first time - this is really unfair on those of us who (successfully) worked hard to attain the exams prior to ST3 applications. The weighting of portfolio to interview was disproportionately skewed compared to normal - moving goalposts in favour of an arbitrary tick-box points system is totally unfair on those of us driven, focussed and motivated trainees who have come straight through training without career breaks and changes. | 0.019 |
| 31-35 | Male | Acute Care Common Stem (ACCS) - Anaesthetics Trainee | It’s been somewhat of a slap in the face that this has happened after COVID. Surely the curriculum could have been modernised without the need to move the break between core and registrar training. The system has worked well so far in producing well trained anaesthetic consultants. | 0.02 |
| 25-30 | Male | Acute Care Common Stem (ACCS) - Anaesthetics Trainee | There should have been an obligation/ incentive for deaneries to offer their Core CT2s/ ACCS CT3s a top-up year, with safeguards in place that these will meet the required curriculum objectives and provide trainees access to teaching, study leave etc. While some deaneries have taken this initiative, the demand exceeds the supply of these posts and I’m not sure what assurances are in place to prevent these jobs from becoming pure service provision at the expense of training. | 0.021 |
| 25-30 | Female | Non-training Fellow, SAS or other Trust-Grade post | I have always truly enjoyed and loved anaesthetics as a specialty, and would always highly recommend it to anyone thinking of applying at CT1 level. Now I’m not so sure- I feel really disillusioned by the way the application process has panned out, with the potential for 2 years out of a training programme and no guarantee of an ST4 job in 2023 as there will likely be another huge bottleneck at this point. Coupled with an exhausting and difficult 12+ months of covid, I feel undervalued and unappreciated. It’s making me seriously consider dropping out of a specialty that I otherwise love. | 0.024 |
| 31-35 | Male | Non-training Fellow, SAS or other Trust-Grade post | I find it disgraceful that there was no explanation as to why there were so few ST3 jobs advertised. The decision to do so clearly had some rationale, but the fact this was not communicated to anaesthetic trainees whose lives that decision affected is inexplicable. The person or people who thought this was a good idea should have fronted up and explained to the hundreds of anaesthetic trainees who have passed every assessment asked of them, and gone the extra mile during the pandemic why so few jobs were advertised. The selection process into specialty training is, in my view, one of the most critical functions of the RCOA. The resource allocated to this function should be commensurate with the importance of the decisions being made. How can it be therefore that just 30 minutes of time is considered adequate to accurately identify the very best candidates? More time is allocated to every doctors annual ARCP for goodness sake! I realise there are pressures on everyone’s time during the pandemic, but it just seems typical that the compromise should be made on the selection process rather than the enumerate other onerous demands on our time. Although I have been offered a job I recognise how flukey this year’s selection process was and feel very strongly on behalf of my friends and colleagues that have done everything right and yet have not been given adequate opportunity to demonstrate their skill as anaesthetists. As you can tell I am seriously disillusioned with the various governing bodies that have overseen this process. I don’t expect that feeling to fade in the future. I do expect a generation of embittered anaesthetic trainees to be spawned by this process. | 0.025 |
| 31-35 | Female | Acute Care Common Stem (ACCS) - Anaesthetics Trainee | I am relocating for my ST3 post and received the news that I had a job through clearing so quite late on. This gives me 2 months to relocate and yet the application process itself has been drawn out over 7 months. This is a 7 month period when you can’t plan any big things in your life as you have no idea what will happen moving forward. This needs to change, we have no autonomy. | 0.025 |
| 31-35 | Male | Non-training Fellow, SAS or other Trust-Grade post | It is frustrating that there is an 18 month gap in recruitment from February 2022 until august 2023. I will have achieved the CT3 top up competencies by august 2022 as will many of us. An explanation as to why we have been abandoned for another year would be nice. I am a more experienced trainee and have come to anaesthetics from an EM background so delays of 18 months are more frustrating and have major impacts on my life away from medicine, possibly more so than for those who have come straight through from foundation. | 0.027 |
| 25-30 | Female | Non-training Fellow, SAS or other Trust-Grade post | I generally feel there has been a complete lack of planning for this situation. Applications for ST3 recruitment closed in December 2020 so it must have been known for almost 5 months exactly how many trainees would be affected by these changes and the systems have not been put in place to support the significant numbers now stuck in between curriculums. This is only now gaining traction because the numbers involved have been released publicly and anger from those involved is growing. It is not clear if there will be enough CT3 equivalent jobs to go around and it is not looking as though the competition ratios will be any better when it comes to applying for ST4. It may well be more than another 2 years to wait to get a training number. It has also been said that CT3 equivalent posts cannot be guaranteed in the area in which you currently work. Particularly after the pandemic, I am not willing to uproot my life to another part of the country to take up a CT3 equivalent clinical fellow post which has absolutely no guarantee of leading to a training number at the end of it. On a more general recruitment note, this is now the 3rd round of national recruitment I have been involved in since I graduated. Every few years we are expected to be open to moving house, county, or even country to take up coveted training numbers. I don’t know of any other professional career that recruits in this way, which essentially amounts to a geographical lottery, and frequently separates families and support networks. Until this point of my career I had never considered moving abroad or leaving medicine. I love my job, but the lack of control we have over our own careers and personal lives when working within medicine in this country has lead me to seriously consider other options now. | 0.03 |
| 36-40 | Male | Anaesthetic Core Trainee | Not enough jobs followed by “extra” jobs appearing after the fact. | 0.03 |
| 31-35 | Female | Acute Care Common Stem (ACCS) - Anaesthetics Trainee | Incredibly disheartening to work very hard for 3 years to achieve a very high score in the self-assessment criteria and to pass the primary first time and receive nothing in return and to have to accept that this is because of bad luck and HEE’s poor planning. Difficult to accept that the curriculum change is due to people not passing the exam in time but receive no reward for passing the exam in time. Overall makes you feel so unimportant and unvalued. | 0.036 |
| 25-30 | Female | Non-training Fellow, SAS or other Trust-Grade post | I appreciate the difficulties in the interview process during this year. I also appreciate the difficulties in the ranking system. However, many have been penalised during the online marking system of the portfolio (for example, being unable to appeal for 1-2 points, which could have great bearing on the final marks). The interview itself was not broad enough to explore different aspects and attributes of a candidate. Further to this, the 30minute interview holds the same weight as a portfolio which has taken years to cultivate. | 0.036 |
| 25-30 | Female | Acute Care Common Stem (ACCS) - Anaesthetics Trainee | I feel I have to delay buying a house and starting a family due to the uncertainty about if there will be any jobs available or where these will - I don’t want to have to uproot my life with a young family. | 0.039 |
| 31-35 | Female | Non-training Fellow, SAS or other Trust-Grade post | A lot of excellent candidates didn’t get jobs. I feel the interview process was quite subjective with a large emphasis on global score, and only one panel with only one style of assessment rather than similar to previous format with multiple assessments (presentation, clinical etc). Every point makes a big difference in terms of ranking yet you can only dispute your self assessment score if the discrepancy was more than 5 points which doesn’t make sense. The goalposts keep changing and I don’t understand why people were allowed to apply without having completed the FRCA primary considering only one exam sitting was cancelled in the last year and there has still been ample opportunity to pass. Due to the self assessment criteria points decreasing after 24 months in complementary specialties (which I already have), I am now in the position of having to decide between doing something that I would enjoy and would be good for my learning and experience and make me a better anaesthetist (eg PICU/retrieval/prehospital) but decrease my self assessment criteria points and doing another job that I don’t want to do such as ICU fellow posts with exhausting on calls which won’t develop me as a clinician/anaesthetist but won’t drop my points. More clarity is required about the new curriculum, and about what the method of assessment will be for entry into ST4 for 2023. There should be some extra ST3/st4 posts for august 2022 given the large number of trainees affected who are caught up in the middle. The discrepancy between CT1 post numbers and ST3 post numbers is absurd, further amplifying the bottleneck. This issue is only going to get worse year on year, especially if the new curriculum improves the attrition rate between core and registrar training as anticipated due to more trainees passing the exam in time to progress. It doesn’t make sense that anaesthetics training doesn’t have a run through option like emergency medicine training. | 0.04 |
| 25-30 | Male | Acute Care Common Stem (ACCS) - Anaesthetics Trainee | The communication regarding interviews and CT3 top up years has been incredibly poor. I’ve had training programme directors apologise to me as their advice has been changing on a monthly basis for the last year. The situation has result in an incredible amount of stress for key workers during a pandemic. Whilst I understand the need for the change, it has been mismanaged and become a shambles. A generation of anaesthetists have lost faith in the system. | 0.041 |
| 31-35 | Female | Anaesthetic Core Trainee | So many amazingly competent friends who are being delayed due to recruitment rather than their worth. So disheartening, even for those of us with jobs | 0.042 |
| 36-40 | Male | Non-training Fellow, SAS or other Trust-Grade post | Although I have significant reservations about the disparity between the number of ST3 posts and those at CT1, my main grievance has been with the new online self assessment score process. It was clear from the self assessment feedback I received that they had incorrectly marked down my score in one of the domains, and had used this as a reason to reduce my global score. I initially submitted an appeal to ANRO, but was told that I did not meet the appeals criteria (as per applicant guidance section 6.3 here <https://anro.wm.hee.nhs.uk/downloads>). In order to qualify for an appeal, your self assessment score (submitted at the time of application) must have reduced by 5 marks or more by the consultant assessor. My score had actually increased between time of application and deadline for document upload as I had done some relevant things in the interim which scored on the self assessment criteria. As such, ANRO used this as a reason not to grant me an appeal. Judging from social media this seemed to be a common issue, and the chair of the RCOA Anaesthetists in Training(AiT) Committee tweeted that those who did not reach the criteria for an appeal should lodge a formal complaint with ANRO. This was also echoed in an email sent out by the RCOA. I did just this, but received a similar reply; that I didn’t qualify for an appeal and had no grounds for a complaint (the complaints procedure is here - <https://specialtytraining.hee.nhs.uk/portals/1/Content/Resource%20Bank/Recruitment%20Documents/MDRS%20Complaints%20Policy%202021.docx>). I found this extremely frustrating and disheartening; it wasn’t as if someone had re-looked at my evidence and decided they disagreed, they had just refused to look at it. The initial appeals criteria cut-off seemed particularly arbitrary; especially in the context of a highly competitive application process where a single point could make the difference between getting a job and not. I emailed the RCOA AiT (and the Association of Anaesthetists Trainee committee) and received assurances that the RCOA training committee were liaising with ANRO and the MDRS to highlight trainee grievances with the system. Shortly afterwards ANRO announced they were reopening another appeal window, with limited criteria (basically for clerical or admin errors); for which I didn’t qualify. I then didn’t hear anymore about it. I discovered on social media (an hour before the deadline) that ANRO had emailed candidates who had been marked down between 1 and 4 marks details of the new appeals process; an email that I had not received. I am extremely fortunate that I happened to see this on social media, as I was able to submit a last minute appeal. On review my appeal was upheld and I was given 5 (!) extra points. I have no doubt that these extra points helped me secure the job I wanted. I feel very strongly that at present the online self assessment system is not consistently marked and does not have a robust, transparent or fair appeals process. I had to spend considerable time and effort to get my appeal reviewed, which is not fair or right, and added significant stress to the process. I am extremely fortunate to have a job at the end of this, but I suspect many of my colleagues have been disadvantaged by the current system. The appeals process should be open to everyone if they feel that a mistake has been made, and there should be no arbitrary points cut-off. | 0.043 |
| 25-30 | Female | Anaesthetic Core Trainee | I am concerned that there will now always be a backlog of applicants, as those unsuccessful this time round then saturate the next round of recruitment. Are there plans to increase training numbers? I am also concerned about maternity leave when not in a training post. I hope to start a family soon but don’t know where I’d stand if I don’t manage to secure a CT3 top up and lose maternity leave entitlement? If I am successful, what then happens when my contract expires the following year or 6 months - I imagine pretty difficult to convince someone to employ you if pregnant and likely to need may leave. 2 years feels a long time to wait to start a family just so I can have some job security. It really doesn’t feel like there’s enough advice around this for doctors who are not in training posts. | 0.046 |
| 25-30 | Male | Intensive Care Medicine Trainee | Unclear about ability to take time out of ICM programme for ct3 top up year. Time pressure on achieving anaesthetics post for dual training | 0.046 |
| 25-30 | Male | Anaesthetic Core Trainee | - Generic, non-personal, interview question - Standardised questioning over a 2 week period disadvantage for those who went earlier in the process - Lack of requirement of full FRCA primary favours those who have left the exam later in their training and would normally expect to take clinical fellow posts to complete exams - Inability to plan starting a family, buying a new home, and settling down due to the complete lack of control I have over my career. - Feeling that career should be valued above all else to obtain an ST3 post is completely demoralising | 0.047 |
| 25-30 | Female | Non-training Fellow, SAS or other Trust-Grade post | High pressured stakes in terms of obtaining points and high pressured interview in terms of the short time of the interview to present myself as a good candidate | 0.047 |
| 31-35 | Male | Acute Care Common Stem (ACCS) - Anaesthetics Trainee | Health Education North East have failed to facilitate the progression of their own trainees. Instead, favouring to saturate the intermediate and higher training numbers with LTFT. HENE ACCS Anaes continues to recruit numbers of which on a fraction will progress to ST3. | 0.049 |
| 31-35 | Male | MTI Fellow | The support offered to MTI colleagues in general is definitely far less than what the core and specialty trainees with training numbers get. Most of us don’t have prior knowledge about starting audit or QI works. Also MTI fellows start their training at intermediate level. Still there is a lot of confusion about achieving core level equivalence to apply for the trainee posts. Also it’s been difficult to get a block in a sub-specialty of interest most of the times. We sometimes feel unheard. I appreciate the College’s recent publication on guidance for trusts offering MTI post. Despite this, I believe still a lot of clarity is needed. | 0.049 |
| 31-35 | Female | Anaesthetic Core Trainee | Severn, yet again, released 10 jobs then added additional jobs later (now up to 16)… what game are they playing?? Having a year without recruitment puts so much stress on the 2023 ST4 round. I don’t mind not having a job this time round but it’ll be a total disaster not getting one in 2023. | 0.05 |
| 25-30 | Male | Anaesthetic Core Trainee | The guidance around CT3 top ups are vague, with no clear idea of how we are supposed to evidence the competencies prior to ST4 training. | 0.05 |
| 25-30 | Male | Acute Care Common Stem (ACCS) - Anaesthetics Trainee | This year’s application has knocked my confidence and has caused me to question my desire to remain in the profession. I feel deeply uncertain about the future and it seems that my hard work over the last 3 years of core training has, in the event, amounted to very little. | 0.054 |
| 25-30 | Female | Non-training Fellow, SAS or other Trust-Grade post | Subjective self assessment process, lack of oppportunity to appeal unless score varied widely to one awarded, interview marred with technical difficulties, jobs awarded to those with no exam and less experience- makes those with exam and more experience feel extremely undervalued. | 0.055 |
| 36-40 | Male | Intensive Care Medicine Trainee | This was my sixth application and fifth interview to ST3 anaesthetics. This was in part due to the fact that was only able to rank the Severn deanery due to family reasons, and then because I accepted an ICM number here. Due to very high competition levels, several attempts before gaining a number are common in Severn, usually people try for 2, 3 or 4 times before becoming successful or moving region, country or leaving the specialty. I believe this is caused in part by HEEs decision to recruit more SHOs than Registrars, which appears to serve the purpose of providing a driving pressure to move trainees to less popular areas at ST3. This is at the expense of trainees in popular areas. Many of my friends have not returned to the UK or entered GP training to avoid the Severn ST3 anaesthesia bottleneck. This is a great loss to the specialty as these are high performing trainees who performed well at interview, but did not clear the high bar required for this region. I would favour reducing the number of CT1 posts in the popular regions and increasing the number of ST3 posts there . People are more likely to move at CT1, and so increasing CT1 numbers in less popular areas should result in higher fill rates later at ST3. I also feel that the interview is very poor at differentiating between good candidates. To receives an offer in Severn one usually needs to score within the top ten to fifteen percent of interviewed candidates. A few marks make a huge difference, and much of the CV score relates to things that have no bearing on your ability to make the a good consultant anaesthetist. Prizes depend on the medical school more than the individual, intercalations are ridiculously over rewarded and working as a medical registrar is objectively better preparation for anaesthesia than working as an SHO. Not only are the marks in these areas poorly thought out, but they are historical and cannot be changed or improved on. The most impressive lines in my CV did not score me any points, and there was no opportunity to discuss them as they did not fit into the rigid framework of the interview either. I have eventually been successful by working hard on the aspects of my CV that I could change, but it took one interviewer to give me the full forty marks before I was eventually offered a job. By contrast in the ICM interview I interviewed twice for Severn, receiving offers both times. Overall, I believe the interview process has been designed with national goals in mind and discriminates poorly between candidates. It has been very stressful stage of my career, and I am extremely grateful to the regional CDs, college tutors and subspecialty leads who allowed me to carry on training outside of the training programme during this process. | 0.056 |
| 25-30 | Male | Non-training Fellow, SAS or other Trust-Grade post | Additional training numbers for my first choice region were only released AFTER the upgrade window had closed. This meant 4 candidates ranked below me in the system got offered the job I originally wanted leaving me completely deflated and feeling undervalued. It brings in to question the whole point in having a ranking system or an upgrade window. I have been penalised for offering to be an Anaesthetist in any region of Scotland where my colleagues who would only do the job in a certain region have been rewarded. This has had a huge impact on my life and the life of my partner who is having to relocate with me. I have had to sell my current residence and will suffer significant financial loss paying to move region and cover the associated legal fees all because I was committed enough to the specialty to do the job anywhere. | 0.059 |
| 31-35 | Female | Intensive Care Medicine Trainee | It’s frustrating and upsetting to have worked hard through core anaesthetic training dealing the significant stress and disruption to work and life caused by covid and now not be offered the chance to continue that training. I really enjoyed anaesthetics, passed the exams and portfolio and had good feedback from all my placements. Also had good feedback from my portfolio and interview at application but have failed to get a job at 3 rounds. It makes me sad because I think I’d have enjoyed anaesthetics as a career but after covid I’m not sure I’ve got the energy or motivation to continue with the uncertainty of multiple applications and repeated rejections especially with the complications of CT3/ST4. I’ve got an intensive care number and planned to duel CCT but am now considering single specialty. I’m not sure about the sustainability of this as a career long term so I am also considering time out of training to work/travel abroad and maybe look at career options outside of medicine. | 0.059 |
| 31-35 | Female | Non-training Fellow, SAS or other Trust-Grade post | The whole process of preparing the application was stressful and following the guidance on the E portfolio carefully and not overloading the sections, was probably against me and not uploading a full CV. It has left with me with a lot of uncertainties, and it’s frustrating that I’m being forced to take more time out than planned. I will try to make the most of it but I wonder if I’ll still be considered eligible and not overtrained by the time I apply for ST4. | 0.059 |
| 31-35 | Male | Non-training Fellow, SAS or other Trust-Grade post | Number of jobs in some regions as well as lack of intake for a whole year left me and otherw in a really challenging place. There were only 9 jobs across pennisula/wessex and severn. There have been 30odd CT jobs in pennisula each year for several years. The backlog in the West Country is nothing new and has already caused several fantastic anaesthetists to move to other careers. Fewer and fewer are able or willing to substantially change region at ST3/4 application as many more are taking time out to do other things and are older and more settled. Consideration needs to be given to the regional balance at CT and ST intake which in some areas seems very imbalanced. | 0.059 |
| 25-30 | Male | Acute Care Common Stem (ACCS) - Anaesthetics Trainee | I felt I had no choice but to apply for an ST3 post, despite having had very limited clinical anaesthetic experience due to repeated redeployments. I had been planning to take a year out of training to consolidate my experience, but was extremely concerned by the plans for CT3 top-up posts. It is incredibly risky to rely on trusts to provide posts that enable CT3 competences to be met. I was not prepared to gamble with my anaesthetic career in this way. | 0.062 |
| 31-35 | Female | Non-training Fellow, SAS or other Trust-Grade post | Despite providing evidence that I did a fulltime research post for 1 year I got marked 0 for research with the comment that without a copy of the protocol they couldn’t assess the domain. I also provided PowerPoint slides, confirmation email and programme of events for a conference I spoke at. Again I was deducted these marks because I had not provided a letter of thanks from the organisers so they felt they had no evidence that I had actually turned up and delivered the talk. I asked for a review and no changes were made. I scored very low on my application. The level of suspicion my application was treated with made me doubt whether a birth certificate would even have been accepted as proof of my date of birth. | 0.065 |
| 31-35 | Male | Non-training Fellow, SAS or other Trust-Grade post | I have lost faith in health education England and feel demotivated about the future. | 0.067 |
| 25-30 | Male | Acute Care Common Stem (ACCS) - Anaesthetics Trainee | The relative lack of number of ST3 posts nationally, despite surveys such as the RCOA census demonstrating a growing gap in the anaesthetic workforce and the need for more anaesthetists to overcome the backlog of operations not undertaken during the pandemic. | 0.067 |
| 25-30 | Male | Anaesthetic Core Trainee | Many concerns. First and foremost why is training not run-through? ST3 applications generate a lot of stress and soak up a lot of time (for both trainees and consultants), but to what benefit? Second, the metrics measured in the applications mostly have little to do with clinical competence as an anaesthetist - there is no attempt to measure if you’re a good colleague, have great technical skills not shrewd clinical acumen; ICM applications at least attempt this by including a section on consultant feedback. Yet you do get points for having a prize in medical school irrespective of its relevance to anaesthesia… Next, why are there consistently fewer ST numbers than CT numbers? The anaesthetic training attrition rate is relatively low and is not entirely concentrated at the Core/Specialty checkpoint. Furthermore the appointed ST posts do not account for Whole Time Equivalents, with the increase in LTFT, parental leave and academic leave/OOPEs ST rotas are consistently under-filled affecting training opportunities due to the relative increase in service provision. Finally in Scotland when additional posts were funded due to existing anaesthetic ST trainees gaining ICM numbers (and thus having their funding taken over by ICM) these were distributed not to the trainees who otherwise would have been eligible for upgrades earlier in the process, but to lower ranked trainees who had yet to be offered a job due to inflexibility about where they were prepared to work. While this may be the letter of the existing rules, it seems to ‘punish’ flexibility setting little value on the financial, social and emotional costs that that flexibility incurs. Ultimately it undermines the sense that the application process is a true meritocracy (at least within the limits of the metrics assessed). | 0.068 |
| 31-35 | Male | Non-training Fellow, SAS or other Trust-Grade post | I have many concerns around ST3 recruitment over the past 2 years. Previous recruitment involved solely a self assessment which was not verified in any way. It is impossible to view this as a fair and transparent way to run a recruitment process. There was also the removal of the Primary FRCA OSCE/SOE as essential criteria for starting st3. This has directly led to the increase in applicants for st3 posts when there was already processes in place for those who were not able to complete the primary FRCA in core training. Allowing applicants without the exam to progress poses a potential risk to patient safety and should never have been considered as an appropriate step to take | 0.07 |
| 31-35 | Female | Non-training Fellow, SAS or other Trust-Grade post | With regards to the online portfolio verification process, I didn’t feel that I was treated as a person, but as a number. I am concerned about the subjectivity of the process, and subsequent discrepancies in scores that I and many colleagues experienced. I am also concerned that I raised an appeal and didn’t feel that it was given due attention, it felt as if there had been an automatic response of just agreeing with the original assessor. I was happy with how the interview was conducted, but felt that I never really stood a chance due to my portfolio being under scored. The whole process has left me feeling frustrated and under appreciated. | 0.072 |
| 31-35 | Male | Non-training Fellow, SAS or other Trust-Grade post | Continued excess of core trainee to registrar places. Portfolio points are difficult to improve over the short term - extra degrees, teaching jobs. Surely portfolio could include review of feedback at work etc.? Why are there no st4 jobs in august 2022? There are many of us who will have the competencies by then. Likely to end up having been out of training for 30 months if I get an ST4 job at the first time of asking. Massively extending my training. | 0.072 |
| 25-30 | Female | Not currently in contracted employment, or currently a locum | I was withdrawn from the process by accident as the new clinical portal was hard to find and I had all my documents uploaded on oriel and not that portal and they refused to look or let me move it over. I found this unfair as I was working flat out in covid icu and it was an honest mistake I was penalised…not 6 months as would be normal….but to 2023! | 0.073 |
| 25-30 | Female | Anaesthetic Core Trainee | The applications felt very rushed and the bottleneck has increased the reliance on portfolio scoring which felt very tick-box .There is only limited things you can do to improve the score given the limited courses/ conferences running due to covid. The fact that they let people with only MCQ’s to apply have also further increased the competittion. | 0.074 |
| 25-30 | Male | Anaesthetic Core Trainee | Just feel completely let down by the college. After spending so much time and effort going above and beyond for them to feel that this is the right time to implement a huge change that leaves lots of trainees without jobs just feels awful. | 0.075 |
| 31-35 | Female | Non-training Fellow, SAS or other Trust-Grade post | I found this years ST3 applications to be difficult and inequitable. I was one of the trainees who was fortunate to be offered an upgrade through round 2 of ANRO recruitment to ST3 anaesthetics to SE Scotland. Unfortunately several of my colleagues who ranked higher than me in the interview/ portfolio scoring system, but who had also applied for other regions, and accepted with upgrades, did not get offered an upgrade.Â  This is because the recycled upgrades were only released after the deadline for ‘accepting with upgrades’ closed. So deserving colleagues who desperately wanted Edinburgh ST3 jobs, and had ranked sufficiently high to get these jobs, have missed out. This seems exceptionally unfair. Had these jobs been recycled a week earlier (it was known they were available at least 2 weeks prior to the deadline for accepting with upgrades), then these people would have got the upgrades they deserve. It is unreasonable of the college to ask trainees to jump through numerous hoops to distinguish ourselves from our colleagues in order to get a ranking, and then completely ignore that ranking when distributing jobs. | 0.075 |
| 25-30 | Female | Anaesthetic Core Trainee | I am worried that despite having a good score that I will have further difficulty when I reach application for st4. This makes me want to leave anaesthesia completely despite loving the specialty. | 0.075 |
| 31-35 | Male | Intensive Care Medicine Trainee | I intend to dual CCT, and was offered an Anaesthetics ST3 job in 2020, only to decline this in preference for ICM. I have since been unable to obtain another Anaesthetics ST3 offer, and I find the whole sequence of events completely ridiculous. Dual appointment should be possible, and encouraged, at the same time. | 0.076 |
| 31-35 | Female | Acute Care Common Stem (ACCS) - Anaesthetics Trainee | I worked exceptionally hard during the pandemic. I sacrificed my education, my revision, my mental health, and my chance of career progression, to serve the NHS throughout both waves. I didn’t apply for an ST3 post as I recognised that I needed time away from training to allow my education and my mind to recover. I have been unable to revise adequately to pass my exams because of surge rotas. I have missed out on many educational opportunities. I felt that, my recognising all of this, I was doing the right thing. But now I feel like I’m being penalised for not being able to keep up with everyone and find the time (how? When would I have been able to find the time?!) to study, pass my exams and prepare for interview. Had I known that I would lose two years - first to a CT3 top up year, and then another to a recruitment freeze (which I cannot see the reason behind and I feel that the RCOA or GMC have been very underhand in doing) - I would have applied this year. Now I have to waste two years of my life, and then fight with exceptionally high numbers of applicants - from several years of training! - to get a registrar job. I wanted to start a family over the next year. Now it’s going to be at least another two years before I can. I expected there to be a price to pay from COVID, but I didn’t think I’d be paying it to my own college. I didn’t sacrifice everything during the pandemic for the promise of some reward, but this is no way to repay us for all our hard work. I’m just being punished. Why? | 0.077 |
| 31-35 | Female | Anaesthetic Core Trainee | I found it difficult to feel I had given my best in the online interview format, it is hard to engage people from behind a screen and the questions asked were strange. | 0.08 |
| 36-40 | Male | Non-training Fellow, SAS or other Trust-Grade post | Insufficient jobs available creating an even bigger bottleneck in career progression at a time when there are in reality 50% underfill of middle grade anaesthetic rotas. | 0.08 |
| 31-35 | Male | Non-training Fellow, SAS or other Trust-Grade post | I think it was a tough year to apply but the application process was fair. My biggest criticism would be the portfolio marking. I got my score increased by 15 points on appeal. Whilst in my case I was delighted it shows you how much variation there can be when people mark portfolios. I also got negative feedback the first time in capital letters - I found that a bit hurtful and unnecessary. I got an ST3 job but I really hope the AAGBI and RCOA support all of these talented clinicians who didn’t get a job and help them stay in anaesthetics. | 0.081 |
| 31-35 | Male | Anaesthetic Core Trainee | In general - given everything that has happened with the pandemic over the last year I feel incredibly let down by the whole situation. If unsuccessful in applications for CT3 top up jobs which have now also become extremely competitive I may consider my future in medicine and look at alternate careers where I feel more valued. | 0.081 |
| 31-35 | Male | Anaesthetic Core Trainee | Extremely concerning the situation. Lack of job security and assurances for the future considering anaesthetics is a focused training programme. | 0.081 |
| 31-35 | Female | Non-training Fellow, SAS or other Trust-Grade post | I had to extend my core training to pass the Primary Viva and then COVID hit and I was not granted a job in the pure self assessment round because I had spent 3 years focussed on passing the Primary FRCA instead of boosting my CV. Then this year the fact that I have passed the full Primary FRCA is irrelevant. At this rate I will have completed 7 years of Anaesthetics training and be 37 years old by the time I get a training number. If I have children in the next few years then that will delay things further and I might not be a consultant until I am at least 45 years old. I am absolutely committed to Anaesthetics and am a very good doctor clinically but am being punished by a system that rewards points for things that I am not interested in like research and audit. I do not want to be a speciality doctor. I want to be a consultant so that I can implement changes and support trainees in the way that I have been supported. | 0.084 |
| 36-40 | Male | Non-training Fellow, SAS or other Trust-Grade post | Obviously there is an issue with training numbers at ST3. This is acting as a severe bottleneck and I would like to see the AoA and RCOA being aggressive in their lobbying of government. In the meantime, please put pressure on trusts to turn LAS posts into LATs so people can at least advance their careers. I’d like to see more information regarding the points required to secure posts in certain regions - ideally sharing information such as score at interview and portfolio. This would allow informed decisions about where to apply to and set a threshold score to aim at for prospective trainees. I would also like to see competition ratios for each region which I know HEE has sporadically published previously. | 0.086 |
| 31-35 | Male | Non-training Fellow, SAS or other Trust-Grade post | I think there has been very little understanding or conversation with anaesthetic trainees who have completed core training, and are now put in a position like this due to curriculum changes. The timing of these changes could not be at a worse time, and has been a real disappointment for candidates like myself. The application process is also flawed with regards to identifying those who would make good trainees, and I believe there should be an overhaul of the application system. With regards to my interview, I was unable to see the interviewers throughout the interview, and when I informed recruitment of this, I was told it made no difference to my interview because the interviewers were not aware (zero consideration for how it may have affected my performance)! Finally, the communication with trainees has been abysmal, with most communication coming via Twitter - how is this appropriate!? | 0.087 |
| 25-30 | Male | Anaesthetic Core Trainee | CESR has a place in medical specialties for individual trainees who have gone off the ‘standard’ route through training/taken time out etc. If CESR starts being seriously suggested to CT2 anaesthetists who have been keen on the specialty since medical school and done all sorts of extra things around trying to get through training as efficiently as possible, there is something wrong with the system! | 0.088 |
| 25-30 | Female | Non-training Fellow, SAS or other Trust-Grade post | Concern in my region of several posts being made available from relinquished ICM numbers after the upgrade deadline. Those who scored highly but accepted their second choice jobs were not able to be upgraded and lower ranking candidates who had only ranked their first choice received the posts. When the system is designed to favour higher ranking candidates this was unfair. It is in the small print of the recruitment documents but surely relinquished Anaesthetic numbers from ICM should have been known about before the end of the upgrade deadline | 0.091 |
| 31-35 | Male | Acute Care Common Stem (ACCS) - Anaesthetics Trainee | How is the royal college going to ensure the same doesn’t happen in 2 years time? There has to be more funding for more jobs otherwise the bottleneck will only persist and may even worsen | 0.092 |
| 25-30 | Male | Anaesthetic Core Trainee | I’m still not sure how they decided to penalise my portfolio. Valuable and useful pieces of work become worth nothing, quick and facile work to earn CV points somehow still stands. Either I choose to play the points game or I choose to do something meaningful. I feel forced to do something meaningful, so although I hate it, I’ll probably leave anaesthetics. | 0.093 |
| 31-35 | Female | Non-training Fellow, SAS or other Trust-Grade post | Not enough posts for number of applicants. | 0.094 |
| 36-40 | Female | Acute Care Common Stem (ACCS) - Anaesthetics Trainee | I am concerned that the CT3 ‘Top Up’ posts available are in fact not funded posts by HEE and as such are not true training posts. The payscale and contract are different and it will mean a paycut or loss of annual leave for some trainees. In addition there is no guaranteed training which is at the goodwill of the hospital and department that you are working in - how can we be assured these unfunded posts will fulfil the training gap we have ben left in? Furthermore, as a trainee in my late 30’s I had been looking forward to buying a house and ‘settling down’ after nearly 10 years of rotational posts between foundation training and core training. I now have another 2 years before I can even think of doing this if I want to continue to pursue the career i have heavily invested in so far. This is if I am lucky to get a post in 2023, where the competition for ST4 places will likely be even higher unless there is a significant increase in the number of training posts available to account for the bottleneck. To remain ‘competitive’ as an applicant in 2023 I have to continue to maintain my (very expensive) ‘courses’ for points on application (eg ATLS,APLS etc at hundreds of pounds a go) without access to the training budget trainees have to do this. My training will be prolonged by at least a further year if not more. I feel very letdown, unsure about my future and whether I should continue to pursue anaesthetic training or if I am better off looking at other avenues. | 0.097 |
| 25-30 | Female | Anaesthetic Core Trainee | Very very few CT3 posts available, despite obviously large numbers of people needing them to progress. Feel very hurt and let down by lack of care from RCOA for us after this terrible year. Unfair to give so many of us core training posts without any intention of giving us opportunities to progress. | 0.099 |
| 31-35 | Male | Acute Care Common Stem (ACCS) - Anaesthetics Trainee | Low numbers of posts, leading to an expected shortfall in the number of anaesthetists. Feels exceptionally short-sighted with no real long-term planning. This is on the back of working exceptionally hard on the frontline during the covid pandemic. Leads to feelings of not being valued by the profession for what we have done. | 0.101 |
| 25-30 | Male | Acute Care Common Stem (ACCS) - Anaesthetics Trainee | The biggest issue for me was the initial announcement that an extra year was being added to the training post I was currently in (ACCS Anaesthetics) for the new curriculum, then later being told that this was no longer the case. I was fortunate enough to get an August 2021 ST3 job in the same area. I had made family plans for an extra year in the area so am not sure what I would have done if I had to move a year “early” | 0.102 |
| 36-40 | Male | Anaesthetic Core Trainee | The remote portfolio combined with the oriel and rcoa self assessment criteria do not correlate. Not all assessors seem to fully understand what they are assessing and therefore do not award scores appropriately. I had to appeal my initial score and was successful in my appeal which was re-marked from a 1 to a 5 in one particular domain! I was offered an interview but due to exams was not able to proceed. Further to that, there are these and other issues with how the current ST3 recruitment is being done in its watered down state, I’m not convinced you are recruiting the best candidates. Meanwhile the ST4 curriculum and subsequent recruitment process changes are being railroaded through, with a hiatus on higher recruitment to boot, seemingly without any real acknowledgment or proper insight of how it is going to affect a significant number of good potential anaesthetic candidates. People who have already demonstrated commitment to speciality and acquired the necessary skills and training. For these reasons I will be seriously considering all options in the coming 6 months including speciality change or even roles outside medicine. | 0.109 |
| 25-30 | Female | Acute Care Common Stem (ACCS) - Anaesthetics Trainee | After the year we have had, and the impact on our training, this has been devastating. I don’t understand why there are not enough jobs, especially when there are so many rota gaps. It is a disgrace that I have had to competitively apply for a post that will be a top up year, while also trying to sit my FRCA. I was accepted into a core training programme. I should be trained until I am eligible to apply for the next stage of training. | 0.112 |
| 31-35 | Male | Non-training Fellow, SAS or other Trust-Grade post | It seems rather cruel that the changes weren’t brought in a graduated/cohort fashion with a new core trainee cohort, and instead wholesale so that hundreds of careers have been needlessly disrupted and thrown into doubt. | 0.117 |
| 31-35 | Female | Non-training Fellow, SAS or other Trust-Grade post | I was disappointed by my interview…I could only see one interviewer as the other had connection problems and I couldn’t see myself so it was very hard to know if I was maintaining eye contact. The interviewer I could see yawned several times during my answers and was drinking a cup of tea/coffee. Still….I can’t complain too much as I got a job offer and so was much luckier than some of my excellent colleagues. | 0.123 |
| 31-35 | Female | Non-training Fellow, SAS or other Trust-Grade post | I am frustrated and feel very let down by the application system. I worked hard during the pandemic and prior to this ensured I passed the primary with plenty of time to apply for ST3 post. I have failed 3 times to gain an ST3 post despite being consistently told I am good trainee and having very good consultant feedback at appraisals. I have spent time in other areas of medicine prior to starting anaesthetic training and am keen not to delay my progression any further. On top of this, I am considering when to start a family, but at present with clinical fellow posts only being given 6 months at a time, and the constant pressure of trying to improve my self assessment score (despite it already being high) and continuous interview practice, I am having to delay those plans as well. I have considered leaving the profession, however really enjoy my day to day job in anaesthetics. I feel the application is flawed and I know many very good trainees in a similar position to myself. After all the hard work during the pandemic, it feels like a ‘slap in the face’ to be honest. And to be 32 and still not able to have a family when I would like to due to job issues despite training hard throughout my career feels very infuriating. | 0.124 |
| 25-30 | Male | Acute Care Common Stem (ACCS) - Anaesthetics Trainee | My feedback stated that I lacked experience and should do a clinical fellow year, having progressed straight from foundation to ACCS. What is the point of running a core training programme if this is apparently insufficient experience in itself to progress to ST3? | 0.125 |
| 25-30 | Male | Anaesthetic Core Trainee | Primary FRCA is not worth any points in the overall process, passing the primary FRCA despite COVID-19 during CT1-2 should be recognised and/or made mandatory again for future applications. It would be interesting to know what proportion of successful ST3 applicants: a) had achieved the full primary FRCA b) whether those who did not have the primary FRCA were still in core training or post CCT As not spending time passing your exam during CT1-2 which is the conventional and expected route up until COVID is actually advantageous compared to spending months revising for an exam that does not contribute any weighting to the overall ST3 process! In addition to this, the rules for current ST3s from Feb 2021 intake for passing the primary FRCA by end of ST3 seem to have been relaxed, another goalpost move and arguably unfair advantage. The interview process seemed targeted at trainees with additional experience in anaesthesia (disadvantaging those who have gone straight through core training without any need for gaps etc.), especially when the first interview question is “what is your clinical experience outside of core anaesthetics!” There is significant weighting given to the interviewers being a single station with fairly generic questions that do not allow any demonstration of clinical acumen. It is an interesting observation to note that for CT1 my rank was in top 50 and for ST3 it was ~600th! The individual interview domains were ranked as “very good” or “typical” whereas global rating was significantly lower with unhelpful feedback comments given. | 0.126 |
| 25-30 | Male | Anaesthetic Core Trainee | Frustrating to have had 16 months of anaesthetics before application deadline, achieved the full primary exam and contributed to several waves of the pandemic through which there were genuinely fewer CV opportunities. Would be interested in how many core trainees on the same timeline got jobs vs those with extra years (not pandemic effected) experience that could count towards points including post core fellows and ACCS. | 0.137 |
| 31-35 | Male | Acute Care Common Stem (ACCS) - Anaesthetics Trainee | It is infuriating that workforce planning has been so poorly handled as to leave so many skilled professionals without a continued path to completion of training. What makes this even worse is the arms race that one must engage in to reach the top. This includes pointless dead end projects of no benefit to patients, expensive courses and qualifications, finished off with a game of fabricating the most beautifully contrived interview answers. The whole process is of little value and is primarily a distraction from being a good clinician. | 0.139 |
| 31-35 | Male | Non-training Fellow, SAS or other Trust-Grade post | It feels like a kick in the teeth after the past year of disruption, redeployment and reduced training opportunities. The route to a new ST4 post seems way off in the distance and I have no idea whether I’ll make it there. | 0.141 |
| 25-30 | Male | Acute Care Common Stem (ACCS) - Anaesthetics Trainee | The guidance for self scored point was not as clear as it could have been. The information about uploading a Cv was under the heading of extra degrees (which you may not have read if you has no extra degrees to declare). I nearly didn’t up loading a CV as I didn’t have any extra degrees to score points for (thank fully I did up load one so was not marked down for this but know a lot of colleagues that didnt) | 0.142 |
| 25-30 | Male | Non-training Fellow, SAS or other Trust-Grade post | The RCOA state gaining a clinical fellow post in anaesthesia will provide the requisite experience to achieve st3 equivalence. This is problematic as ultimately these posts are governed by the employing trusts and there are no safeguards to ensure we will have access to requisite training opportunities. Most departments encompass both ICU and Anaesthetics, therefore it is plausible clinical fellows are often redeployed to ICU to manage staff shortages and end up in limbo unable to achieve st3 equivalence. As someone who completed core anaesthetic training in February 2021 I am currently an “anaesthetic clinical fellow” that mostly provides service provision on ICU and am therefore not much closer to St3 equivalence despite 6 months of additional experience. In addition to this I believe extra marks should have been awarded to individuals that have fully completed the primary FRCA (this is what RCP does). Virtually every individual who applied to August 2021 St3 would have had the opportunity to sit both the written and VIVA/Osce once. I worked hard to pass both first time whilst working on ICU during covid surges in 2020. Individuals who prioritised audit/teaching/research during this time period likely benefitted from this change in rules. Lastly I believe a clinical component to the interview would have enabled better differentiation between candidates applying for ST3. Having only 2 interviewers asking generic questions - I believe it is hard to objectively differentiate the quality of candidate. | 0.143 |
| 25-30 | Female | Anaesthetic Core Trainee | I feel that there was very little planning for what would happen to those who didn’t apply for/get an ST3 post this year. A lot of the information was published only in the last few months with little information on what constitutes an appropriate ‘CT3 top up’ post/how these posts will be provided. The provision of CT3 posts varies greatly across regions and trainees are not guaranteed to be able to access them. The nature of top up posts also vary greatly between deanery supported training posts to trust grade posts with more service provision expectation. The limited ST3 places in February and lack of ST4 recruitment in 2022 leaves trainees who have not been recruited in ST3 this August at a disadvantage and makes life planning (where to settle/buying a house/family planning etc) difficult when individuals don’t know where they will be spending the rest of their training/whether they will get a training in a region of their choice. I have found the lack of foresight and communication frustrating and has made me consider going through a non training route as an alternative. | 0.15 |
| 31-35 | Female | Non-training Fellow, SAS or other Trust-Grade post | Overall, I welcome the interview process as a healthy balance with which to offset the numerical self assessment: it enables candidates who are more clinical than academic to level the playing field, or candidates who are not driven by “box-ticking,” but who might be very wholesome and balanced anaesthetists, to showcase themselves. However I worry that one interview with one panel leads to an increase in the subjectivity and “luck” of the process. I had an interview for the February 2021 intake, and my panel at the time scored me as being an “average to poor” candidate. This came as a horrible shock at the time - I was very well regarded at work by my seniors, peers and colleagues, but I acknowledge my interview technique lead to me not coming across as well as I could have. I recall “not clicking” with my interviewers in those first minutes (I had that feeling within a few minutes that my way of answering had irritated one of them). As my awareness of this increased, so I was losing confidence as the interview progressed (and exacerbating the problem). In short, it was a bad interview. 6 months down the line, with practice and training, I was re-interviewed for the August entry, and was marked out as an “outstanding” candidate. I clearly benefited from the hard work put into practicing - and am sure that was the key difference. I realise that the two interviewers on the panel mark separately, and would like to commend the efforts already made to make the process fair. However, this experience has led me to wonder if one interview leaves too much to subjectivity and chance for such a significant appointment. 6 months separated my two interviews, and although I worked hard on improving my technique, I had not fundamentally changed the content of my answers. I wonder whether there should be more than one interview such as in the face-to-face process: eg. one general interview, one clinical interview with two different consultants in each, in order to dilute individual interviewer subjectivity, and insodoing perhaps improve the accuracy with which they might measure a candidate’s quality. | 0.151 |
| 31-35 | Male | Non-training Fellow, SAS or other Trust-Grade post | Interview process was treated as a VIVA station rather than a conversation/discussion between adult healthcare professionals. Also allowing applicants without the full primary FRCA reduced the chances of many in getting jobs. | 0.155 |
| 31-35 | Female | Non-training Fellow, SAS or other Trust-Grade post | I prepared and worked very hard on my application throughout the peak of the second wave of the pandemic. I spent time putting together my self assessment score and then ensuring I had the evidence, involving chasing consultants for signatures and hours of extra activities on my days off. I am published, have been involved in many projects, had posters presented at international conferences, am undertaking a masters related to perioperative medicine and have good teaching experience. I did well in my interview (hours and hours of practice on zoom with helpful senior colleagues and consultants), with really positive feedback. Last summer I couldn’t apply for ST3 application because I had not passed my OSCE. I passed this at the first sitting possible last November. It really grates that so many people have been allowed to apply who don’t even have the full Primary FRCA! Others of us have the full Primary FRCA, a good CV and are generally good applicants, yet can’t achieve a training post in the part of the country they live in! As for your question regarding will I have achieved the necessary competencies to be an ST4 applicant in August 2023… By that time, I will have got the Final FRCA, completed my CT3 top up and hence stage 1 training, hopefully finished my masters and have a considerable amount of anaesthetic experience! Maybe all of those things at that point will mean I am eligible for a training post at that point! | 0.156 |
| 25-30 | Male | Anaesthetic Core Trainee | Unable to plan family, accommodation, nursery and school waiting lists | 0.158 |
| 25-30 | Female | Anaesthetic Core Trainee | High competition for trust grade /ct3 top up posts. Concerned I will not be able to get a job in anaesthetics from august. | 0.161 |
| 31-35 | Female | Acute Care Common Stem (ACCS) - Anaesthetics Trainee | The relaxation of criteria regarding exams has negated the hard work that some of us have put into studying to pass the exams during COVID. Jobs have been given to trainees who have yet to complete the exam, a previous limiting factor to continuing training. Previously, these trainees would have had to take a year out to complete the exam. The bottle neck of applicants vs job availability is surely going to increase in size. There will be no jobs for 2 years, but extra applicants due to the addition of the current core trainees completing their core training. | 0.163 |
| 25-30 | Female | Anaesthetic Core Trainee | I took a job in a deanery I would not ordinarily have accepted a number in because of the change in curriculum and competition ratio. On balance, being in a training programme is a better guarantee towards my completion of training, and offers better pay/ study leave/ budgets than holding trust grade jobs. I also work on a tier 2 visa, which meant that going into training was a more secure route to having it renewed (the visa expires immediately after the end of my CT2 year and if I had not secured a CT3 job well before this with sufficient time to renew the visa I would be forced to leave the country). However, I am still deeply unsatisfied with the process because it now means that I work in a completely different deanery to my husband who is also medical, will not be eligible for IDT, and will be away from the home that we have just bought. I do not feel that our personal circumstances are considered with much weight by HEE, and some additional flexibility to the IDT process would promote wellbeing and make training more sustainable. Separately, I also think that the recruitment process is not fit for purpose and fails to discriminate between so many excellent candidates. The interview process is also not objective, open to great variability between interviewers and susceptible to a wide range of bias. | 0.164 |
| 25-30 | Male | Non-training Fellow, SAS or other Trust-Grade post | It’s incredibly disappointing. I missed out on a job despite being deemed appointable at interview. I’ve passed my primary, had an incredibly taxing year working in ICU during the covid pandemic, and to now be left in limbo like this is incredibly frustrating. I love anaesthetics, and I really invisage a happy and fulfilling career as an anaesthetist, but to be honest this has left me, and lots of my colleagues in similar positions feeling deflated, disrespected and disenchanted with the idea. | 0.167 |
| 25-30 | Male | Non-training Fellow, SAS or other Trust-Grade post | The backlog of applications will still be there in 2 years time, or potentially even worse. It’s all very well describing a work force gap, but short of producing more trainees, theres not a huge number of solutions. | 0.174 |
| 31-35 | Male | Non-training Fellow, SAS or other Trust-Grade post | Increasingly high standards will require more time and effort to build a competitive CV/portfolio | 0.176 |
| 31-35 | Male | Intensive Care Medicine Trainee | Having such a bottleneck at ST3 level is ridiculous. We are specialists on a pathway that we can’t turn away from without starting again. I accept that HEE make political decisions regarding numbers of jobs they’ll fund, but the college should have not pushed ahead with the curriculum change during a global pandemic. | 0.176 |
| 36-40 | Female | Anaesthetic Core Trainee | There is no part of the application process which considers how an applicant actually performs in their role. I wonder if referees should be invited to submit a grading of the applicant that would count towards points in the application. Most of the portfolio scores reward things undertaken ‘since starting anaesthetics training’ hence rewarding candidates who chose to gain extra experience (eg abroad/in a different specialty) after completing core training rather than after foundation training before starting core. I struggled to see the logic for this. | 0.177 |
| 25-30 | Male | Non-training Fellow, SAS or other Trust-Grade post | Tied to region due to wife’s career but lack a reason accepted for pre-allocation. Region is ultra competitive and even though I score in the top 8% of candidates, this is not enough to secure role. This has been my 3rd unsuccessful round now and thus I’m considering other career pathways as a result. | 0.178 |
| 25-30 | Male | Anaesthetic Core Trainee | Considerable number of people missing out on training posts will roll over to the next application, combined with increased number at core training level makes for further competition. Concerned I won’t be able to progress despite passing the primary and completing competencies | 0.191 |
| 25-30 | Female | Anaesthetic Core Trainee | I felt that there was no consideration re the impact that covid had on our ability to audit/ present and therefore compete with our unsuccessful clinical fellow colleagues in portfolio. | 0.192 |
| 25-30 | Male | Anaesthetic Core Trainee | The application process doesn’t seem to be based on anaesthetics skill, aptitude or competence but rather on ability to complete paperwork and push portfolio tasks. Given there are only so many hours in a day, this will inevitably lead to less skilled practitioners. | 0.206 |
| 31-35 | Male | Non-training Fellow, SAS or other Trust-Grade post | Whilst I was happy to find out that I was successful in the most recent recruitment round, this has been a long and arduous process for me having been unfortunate in the previous two rounds of application, which were also massively impacted by the pandemic and changes to the anaesthetics curriculum. I have been considering my career plans throughout the last 18 months with moving abroad and switching specialties as definite options. I feel sorry for those guys who didn’t get a job this time, but I am glad to see there are plans in place to help them get the competencies they need via CT3 top up posts. | 0.206 |
| 36-40 | Female | Non-training Fellow, SAS or other Trust-Grade post | Unable to fully explain my CV; penalized for experience; system not designed to reflect you as a doctor, your work ethic and ability, your working relationships with medical and nursing colleagues | 0.207 |
| 25-30 | Female | Anaesthetic Core Trainee | I felt the interview was very rushed. Surely better to have fewer questions to allow candidate time to answer than being bombarded with little time to speak. | 0.217 |
| 36-40 | Female | Non-training Fellow, SAS or other Trust-Grade post | impact on major life decisions/family planning/dependencies | 0.23 |
| 25-30 | Male | Acute Care Common Stem (ACCS) - Anaesthetics Trainee | I prepared very well and worked really hard for my interview and although I eventually got a job, I did not get an offer initially and this caused considerable anxiety. I scored pretty well in both portfolio and interview and yet due to the high ratio of applicants to jobs I still had to wait nervously for a job offer. Having done everything I am required to (completed core training modules, passed Primary exam etc) it is disappointing that it is STILL so difficult to get a job. CAN THE RCOA PLEASE CONSIDER RUN THROUGH TRAINING LIKE ED TRAINING CURRENTLY IS? | 0.234 |
| 31-35 | Female | LAT ST3 | Was not particularly easy to find information about how trainees already in LAT ST3 job would gain CT3 equivalent competency. My college tutor has been very helpful fortunately but without that support I don’t know how I would have found out what I needed to do. | 0.242 |
| 25-30 | Male | Anaesthetic Core Trainee | I’m aware that there are many factors that contributed to the increased competition ratios this year including relaxation of the primary FRCA requirement due to Covid exam disruption, reduced international travel/working abroad opportunities and the curriculum change. Although these variables could not all be controlled, it seems they would all lead to a fairly predictable rise in competition ratios. Although I have seen several statements from the RCOA since application results were released, I do not recall seeing any communications acknowledging this likely issue in the year leading up to this. Given that the increase in competition was fairly predictable, it was disappointing not see any acknowledgement of this prior to applications, with details of how the college would advocate for trainees at this point, or highlighting discussions with HEE raising these concerns etc. Everything feels like it’s been extremely reactive rather than pro-active. 2. I note that in a recent article published in the Independent, Professor Wendy Reid, medical director of HEE stated ‘We recognise this year is unique, which is why we are supporting trainees as much as possible … including additional recruitment rounds and more flexibility for applicants.’ Whilst I am aware this is a statement from HEE rather than the RCOA etc, this seems more than a little ambiguous i.e. when will these ‘additional recruitment rounds’ be, and what format will the increased ‘flexibility’ take? When statements such as this pertain directly to anaesthetic trainees, it would be reassuring to see a direct response from the college/AoA, acknowledging that HEE will be asked to further clarify statements such as these. 3. Finally with regard to the actual application process, I feel that some criteria are slightly unfair when judging trainees. For instance, the section on ‘Undergraduate prizes and awards’ seems particularly unfair given the wide variation in prizes awarded by universities to undergraduates. In addition, this does not seem in any way to recognise the current ability and commitment of anaesthetic trainees, who may have won several awards as post-graduates, but will receive no recognition for this. As a core trainee, I have also found myself at a disadvantage in ‘Postgraduate experience in other specialties’ when compared to my ACCS colleagues. It is hard not to feel that by committing to anaesthetics and working incredibly hard to secure a core training post following my foundation years, I am now penalised for this in my ST3 application having not completed a mandated year in medicine/EM. | 0.243 |
| 31-35 | Male | Acute Care Common Stem (ACCS) - Anaesthetics Trainee | The application for ST4 is going to be even more competitive on return | 0.25 |
| 25-30 | Male | Acute Care Common Stem (ACCS) - Anaesthetics Trainee | This is affecting those at a fairly challenging time - not just COVID but many trainees will be entering a stage of their life where they are desperate for stability. Settling down with a partner, buying a house, having children - these things are all up in the air until there is some stability. | 0.252 |
| 31-35 | Male | Non-training Fellow, SAS or other Trust-Grade post | Felt obliged to accept a lower ranked preference due to scarcity of posts meaning I have to compromise on quality of life to finish training. This has left a feeling of dissatisfaction due to having to accept a suboptimal job and overall means I am less likely to stay in the UK after CCT. | 0.26 |
| 25-30 | Female | Non-training Fellow, SAS or other Trust-Grade post | How big will the bottle neck of trainees applying for ST4 posts in 2023 be? Will I be supported if I qualify through the cesr programme? | 0.267 |
| 36-40 | Female | Non-training Fellow, SAS or other Trust-Grade post | It is difficult to accept that you gain nothing additional on the application for having the full primary as there have now been opportunities for those affected by cancellations to pass it. Having accepted an ICM post, nobody still seems to have any idea how we might go about getting the additional ST3 competencies in anaesthetics in order to be able to apply in 2023, so I am still completely unclear as to whether anaesthetic CCT is now an option without giving up the ICM training number. I can’t understand how it has got to this point and there is no plan for this, or if there is a plan why nobody in the region knows what that plan is. | 0.291 |
| 36-40 | Male | Non-training Fellow, SAS or other Trust-Grade post | The selection process remains astonishingly demoralising, arbitrary, and unimaginative, with little to distinguish between candidates actual merit or commitment to the specialty. How can someone respect an application system where a decision, taken at medical school, to intercalate, is awarded significant credit, but completion of the Final FRCA is awarded none? | 0.295 |
| 25-30 | Male | Anaesthetic Core Trainee | It does not feel as though there is any centralised help in arranging the required CT3 competencies. We have been told to achieve certain things (paediatric, Obs, regional, ITU experience) but knowing that there are almost 700 trainees in the same boat I am concerned that the training opportunities to achieve these competencies will not be easy to come by. There has been no information about how many CT3 equivalent jobs will be available or when these will be available. | 0.296 |
| 25-30 | Male | Non-training Fellow, SAS or other Trust-Grade post | The current system for recruitment does not seem to assess the attributes required for an ST3 post. Academic achievements seem to be prioritised above all else and the options for becoming involved in academia or research vary greatly by region. Specifically this seems to advantage trainees in London where such opportunities are more plentiful. While there has been some disruption to the FRCA primary exam, it is now running in a format that many are familiar with. It does not seem fair to ignore achievement of the full primary exam during application. Finally, with an end to ST3 recruitment coming in the near future, support and specific guidance on how to achieve ‘CT3’ equivalent competencies has been very poor. The opportunities to achieve these as part of trust grade posts are also few and far between. In my view, HEE need to provide funding for posts to allow those stuck after core training to achieve the gap curriculum in order to avoid a lost generation of anaesthetic trainees. | 0.299 |
| 31-35 | Female | Non-training Fellow, SAS or other Trust-Grade post | I am currently very worried, not only about not being able to get an anaesthetic training number in the future because of the imbalance between the numbers of applicants and posts, but at this point I am also worried about how I am going to complete my CT3 top up requirements while going through my ICM training. I know that the college holds the opinion that ICM trainees wishing to do dual training should have priority in completing their CT3 top up, but I don’t have a concrete plan on hand and I worry about becoming ineligible to apply for anaesthetics in the future. | 0.348 |
| 31-35 | Female | Anaesthetic Core Trainee | more reassurances and guidance on how to obain a CT3 top-up post transparency on what the recruitment criteria for ST4 posts will be so that we can work towards making our applicaions as competitive as possible | 0.362 |
| 36-40 | Female | Anaesthetic Core Trainee | reduced number of ST3 jobs available, feedback from verified self assessment scoring | 0.375 |
| 31-35 | Male | Non-training Fellow, SAS or other Trust-Grade post | Difficult to achieve points and more difficult to achieve competencies because of covid | 0.388 |
| 25-30 | Female | Anaesthetic Core Trainee | I was fortunate to be appointed to my first choice ST3 anaesthetics but feel for friends and colleagues who have not been successful and where there is no guaranteed plan in place for top-up posts to allow them to be eligible to apply for ST4 in 2023. | 0.394 |
| 25-30 | Male | Non-training Fellow, SAS or other Trust-Grade post | Fairness and objectivity of online self assessment verification | 0.424 |
| 31-35 | Female | Non-training Fellow, SAS or other Trust-Grade post | Non training post puts me in low priority and provide me lowest opportunity to progress due to where I will be in terms or priority( non trainee) and experience (non SAS). | 0.449 |
| 25-30 | Male | Anaesthetic Core Trainee | I would like there to be clear guidance as to what we need to do for a CT3 equivalent year, along with access to the lifelong learning platform (or something like it) to evidence our training. | 0.467 |
| 25-30 | Male | Anaesthetic Core Trainee | I think it’s extraordinary that under the current system various parties involved in training will with one hand promote wellbeing initiatives, and with the other be complicit in trainees abandoning their friends/family/social support structures when national recruitment schemes require them to move hundreds of miles for jobs to continue training. It’s not complicated - recognising the importance of being able to put down roots, build healthy relationships/friendships outside work, and develop a healthy work/life balance by having more control over where you live is key to promoting happiness. Mindfulness initiatives, whilst still valuable, are clearly not going to make up for abandoning these things to take a training post in a location that you have no links to and are moving to solely because of work. | 0.539 |
| 25-30 | Female | Acute Care Common Stem (ACCS) - Anaesthetics Trainee | Denary has no formal CT3 equivalency plans, left to us to arrange multiple fellow jobs to achieve right competence. | 0.688 |
| 36-40 | Male | Non-training Fellow, SAS or other Trust-Grade post | Admission heavily favour those who have taken extra fellow years or pre anaesthetic medicine etc, putting the dedicated hard working core trainee at a disadvantage | 0.72 |
| 31-35 | Male | Non-training Fellow, SAS or other Trust-Grade post | Provide more training slot. | 0.72 |
