## Supplementary material for "Anaesthetic Higher Specialty Training Recruitment in the United Kingdom During the COVID-19 Pandemic: A National Survey": Full survey questions

### ST3 Recruitment Survey

During the most recent specialty training application round for ST3 Anaesthetics, the number of posts available was far exceeded by the number of applicants. According to figures released by the Royal College of Anaesthetists, 1056 applicants competed for 359 posts at ST3 level this year, while 865 were granted an interview.

This is a difficult time for many trainees who have worked hard over the past 12 months of the pandemic supporting their colleagues on ICUs up and down the country. We sympathise with everyone who missed out on an ST3 post this time around, and would like to help to facilitate everyone in achieving their plans in the next few months.

We would therefore like to find out what trainees who were eligible for this recent round of ST3 applications are planning to do for the next 12 to 24 months. We intend to collect this data to then forward to the relevant stakeholders at the Royal College, Association of Anaesthetists and any organisations planning to employ trainees in the next year.

* Required

1. Were you eligible to apply for an August 2021 ST3 post? *

*Mark only one oval.*

- ( ) Yes *Skip to question 2*
- ( ) No *Skip to question 20*

*Skip to question 2*

#### Demographics

Please tell us a bit about yourself so that we know the profile of those who may have been affected by issues brought about by this recruitment round.

2. What is your age? *

*Mark only one oval.*

- <25
- 25-30
- 31-35
- 36-40
- 41-45
- >45

3. What is your sex? *

*Mark only one oval.*

- Female
- Male
- Prefer not to say
- Other: ____________________

4. Which region are you currently working in? *

*Mark only one oval.*

- ◯ Health Education East Midlands
- ◯ Health Education East of England
- ◯ Health Education Kent, Surrey & Sussex
- ◯ Health Education London
- ◯ Health Education North East
- ◯ Health Education North West (Mersey & North Western)
- ◯ Health Education South West (Peninsula & Severn)
- ◯ Health Education Thames Valley
- ◯ Health Education Wessex
- ◯ Health Education West Midlands
- ◯ Health Education Yorkshire and the Humber
- ◯ NHS Education for Scotland
- ◯ Northern Ireland Medical and Dental Training Agency
- ◯ Health Education and Improvement Wales
- ◯ Outside UK

5. What is your current role? *

Non-training Fellow can include research fellows, SIM fellows, teaching fellows, ICU fellows, etc.

*Check all that apply.*

- ☐ Anaesthetic Core Trainee
- ☐ Acute Care Common Stem (ACCS) - Anaesthetics Trainee
- ☐ Intensive Care Medicine Trainee
- ☐ Non-training Fellow or other Trust-Grade post
- ☐ Not currently in contracted employment, or currently a locum

Other: ☐ ____________________

6. How many years of clinical experience in anaesthesia would you have completed by August 2021? *

Please round to the nearest whole number in years, including any experience abroad.

______________________________

7. What is your current contracted working pattern? *

*Mark only one oval.*

- ( ) Full-time
- ( ) LTFT
- ( ) Not currently contracted, or working as a locum

8. Did you apply for an ST3 Anaesthetics post to commence in August 2021? *

*Mark only one oval.*

- ( ) Yes *Skip to question 9*
- ( ) No *Skip to question 12*

#### ST3 Application Outcome

9. Were you successful with your ST3 application in this application round? *

*Mark only one oval.*

- ( ) Yes *Skip to question 10*
- ( ) No *Skip to question 16*

#### Successful Application

10. Have you accepted an ST3 Anaesthetics post to commence in August 2021? *

*Mark only one oval.*

- ⬭ Yes *Skip to question 20*
- ⬭ No *Skip to question 16*

11. If not - please tell us why?

_______________________________________________

_______________________________________________

_______________________________________________

_______________________________________________

_______________________________________________

*Skip to question 16*

#### Those that didn't apply

We would like to find out a bit more about why you didn't apply for ST3 this year even though you might have been eligible.

12. What was your reason for not applying? *

Please select all that apply

*Check all that apply.*

- [ ] Lack of core experience due to the COVID-19 pandemic
- [ ] Awareness of increased competition ratios and/or reduced CV opportunities during COVID
- [ ] Wanted to take a break from training
- [ ] Wanted to explore another medical specialty other than anaesthesia
- [ ] Wanted to pursue other interests or opportunities, e.g. research or management, etc.

Other: [ ] ____________________

13. At the time of ST3 application were you aware of the planned break in recruitment from August 2022? *

*Mark only one oval.*

- ⬭ Yes
- ⬭ No

14. Did you feel the effects of curriculum changes on recruitment were well-explained to you prior to the ST3 application deadline? *

*Mark only one oval.*

- ⬭ Yes
- ⬭ No
- ⬭ Not sure

15. Would more information regarding changes to the curriculum and recruitment have changed your decision not to apply? *

*Mark only one oval.*

- ⬭ Yes
- ⬭ No
- ⬭ Not sure

*Skip to question 16*

#### August 2021 plans...

We would like to find out what your plans are for the next 12 to 24 months starting in August 2021 if you were not successful with your application for ST3.

16. What are your potential plans for the upcoming 1-2 years? (Select all that apply) *

Non-training fellow includes research fellow, SIM fellow, teaching fellow, ICU fellow etc

*Check all that apply.*

- [ ] CT3 top-up post
- [ ] Non-training fellow post
- [ ] ICM ST3 post
- [ ] Pursue training in a different specialty - specify below
- [ ] Further education, e.g. pursue a higher degree
- [ ] Work abroad
- [ ] Career break
- [ ] Permanently leaving the medical profession

Other: [ ] ______________________________

17. Do you feel confident or assured that you will be able to achieve the necessary competencies to apply for ST4 in August 2023? *

*Mark only one oval.*

( ) Yes

( ) No

( ) Don't know

18. Has the time out of a training programme affected any of your wider life plans, e.g. moving location/family planning/buying a house? *

*Mark only one oval.*

( ) Yes

( ) No

( ) Don't know

19. How confident are you that you will be able to achieve GMC registration on the specialty register in Anaesthesia without a training number? *

*Mark only one oval.*

- Confident
- Not confident
- Not sure

#### End of survey

Thank you for your participation, your opinions on this issue are important to us.

20. What general concerns would you like to share with us regarding this year's ST3 applications? If there are specific personal circumstances you are happy to share with the relevant stakeholders, please feel free to contribute them here. We may publish some of these as anonymised quotes to convey the depth of feeling around this issue.

21. Do you consent to be contacted in the future via email? If so, please provide your email address.

Your email address will not be shared with any third parties. None of your responses will published in such a way that you will be personally identified. We may contact you in the future to follow up on your responses to this survey and/or invite you to participate in a further survey on this topic.

This content is neither created nor endorsed by Google.

Google Forms
